# COVID-19 vaccination and the risk of cardiovascular and thromboembolic events after SARS-CoV-2 infection: a systematic review and meta-analysis

**DOI:** 10.64898/2026.05.21.26353568

**Authors:** Stephane Heymans, Bettina Heidecker, Zoe Marjenberg, Rhiannon Green, Triantafyllos Pliakasf, Gregory Y. H. Lip, Thomas F. Lüscher, Sultan Abduljawad

## Abstract

**Background and Aims:** SARS-CoV-2 infection is associated with an increased risk of cardiovascular, cerebrovascular and venous thromboembolism events. We aimed to assess the impact of COVID-19 vaccination prior to SARS-CoV-2 infection on the risk of these events post-infection.

**Methods:** Embase and MEDLINE were searched from January 2021 to 11 September 2025, supplemented by citation searching. Observational studies were included if they reported risks of cardiovascular, cerebrovascular, or venous thromboembolic events after SARS-CoV-2 infection between different vaccination groups (e.g. unvaccinated, vaccinated, or booster vaccinated), or reported risk of events after SARS-CoV-2 infection compared with no infection, stratified by vaccination status. Random-effects meta-analyses were conducted to estimate pooled hazard ratios (HRs) comparing vaccinated and unvaccinated individuals across prespecified outcomes.

**Results:** Twenty-three studies were included in the systematic review; most reported an association between vaccination and a reduced risk of post-infection vascular events. Ten studies were included across meta-analyses comparing vaccinated and unvaccinated individuals. Pre-infection vaccination was associated with significantly reduced risks of composite cardiovascular/cerebrovascular events (HR 0.60, 95% confidence intervals [CI] 0.51–0.69), stroke (HR 0.75, 95% CI 0.64–0.88), acute coronary syndrome (HR 0.70, 95% CI 0.52–0.95), arrhythmias (HR 0.82, 95% CI 0.69–0.98), and venous thromboembolism (HR 0.51, 95% CI 0.36–0.73). No statistically significant reduction was observed for heart failure (HR 0.72 [95% CI 0.47–1.10]).

**Conclusions:** Pre-infection COVID-19 vaccination is associated with lower risks of cardiovascular, cerebrovascular and venous thromboembolism events following SARS-CoV-2 infection in the pre- and post-Omicron eras, supporting its role within broader prevention strategies.

## Introduction

Severe acute respiratory syndrome coronavirus 2 (SARS-CoV-2), the causative agent of coronavirus disease 2019 (COVID-19), has caused substantial global morbidity and mortality with ongoing seasonal waves of infection since 2019. Although initially characterised as an acute respiratory illness, COVID-19 is now recognised as a multisystem disease with considerable bidirectional cardiovascular involvement during acute infection and throughout post-acute recovery.^1,2,3(p20)^

SARS-CoV-2 infection is associated with increased risks of myocardial injury, heart failure, cardiac arrhythmias, and both arterial and venous thromboembolic complications.^3,4^ Meta-analyses have reported markedly elevated risks of pulmonary embolism, acute cardiovascular and cerebrovascular events, including myocardial infarction, and stroke, with the highest risk observed in the first month immediately following infection and a gradual decline thereafter.^5^ However, large real-world cohort studies have demonstrated that excess cardiovascular risk persists beyond the acute phase, with risks of several clinical outcomes remaining significantly elevated for months and, even in some cases, years after infection.^6–8^ In addition, SARS-CoV-2 infection is associated with an increased incidence of specific cardiovascular risk factors in the months after infection, such as hypertension, dyslipidaemia, diabetes, and chronic kidney disease, underscoring the bidirectional relationships between COVID-19 and cardiometabolic and renal conditions.^9^ Although cardiovascular risk is greatest following severe COVID-19, it remains significantly elevated even after mild, non-hospitalised disease, with risks particularly pronounced among older individuals,^7^ males,^7^ and those with pre-existing cardiovascular or pulmonary disease.^10^

Influenza infections are known to increase the risk of cardiovascular outcomes,^11^ and vaccination has been shown to mitigate this risk. A recent meta-analysis of six randomised controlled trials found that adults receiving the influenza vaccine had a 34% lower risk of major adverse cardiovascular events (MACE) in the year after vaccination versus unvaccinated adults, compared with a 45% lower risk in adults with recent cardiac issues.^12^ Current guidelines accordingly recommend influenza vaccination as a strategy to reduce infection-associated cardiovascular risk.^13,14^ While COVID-19 vaccination has also been proposed as a method of prevention of adverse cardiovascular events after SARS-CoV-2 infection,^14^ the precise extent of the association between COVID-19 vaccination and reduced cardiovascular risk is insufficiently characterised. The objective of this systematic review and meta-analysis was therefore to identify, critically appraise, and quantify the available observational evidence on the impact of COVID-19 vaccination on cardiovascular disease (CVD), cerebrovascular disease (CeVD) and venous thromboembolism (VTE) following SARS-CoV-2 infection.

## Methods

The study protocol was registered with PROSPERO (CRD420251148251), conducted according to Cochrane and Centre for Reviews and Dissemination (CRD) guidelines,^15,16^ and reported according to PRISMA^17^ guidelines (Supplementary material online, Table 1)

**Table 1.**
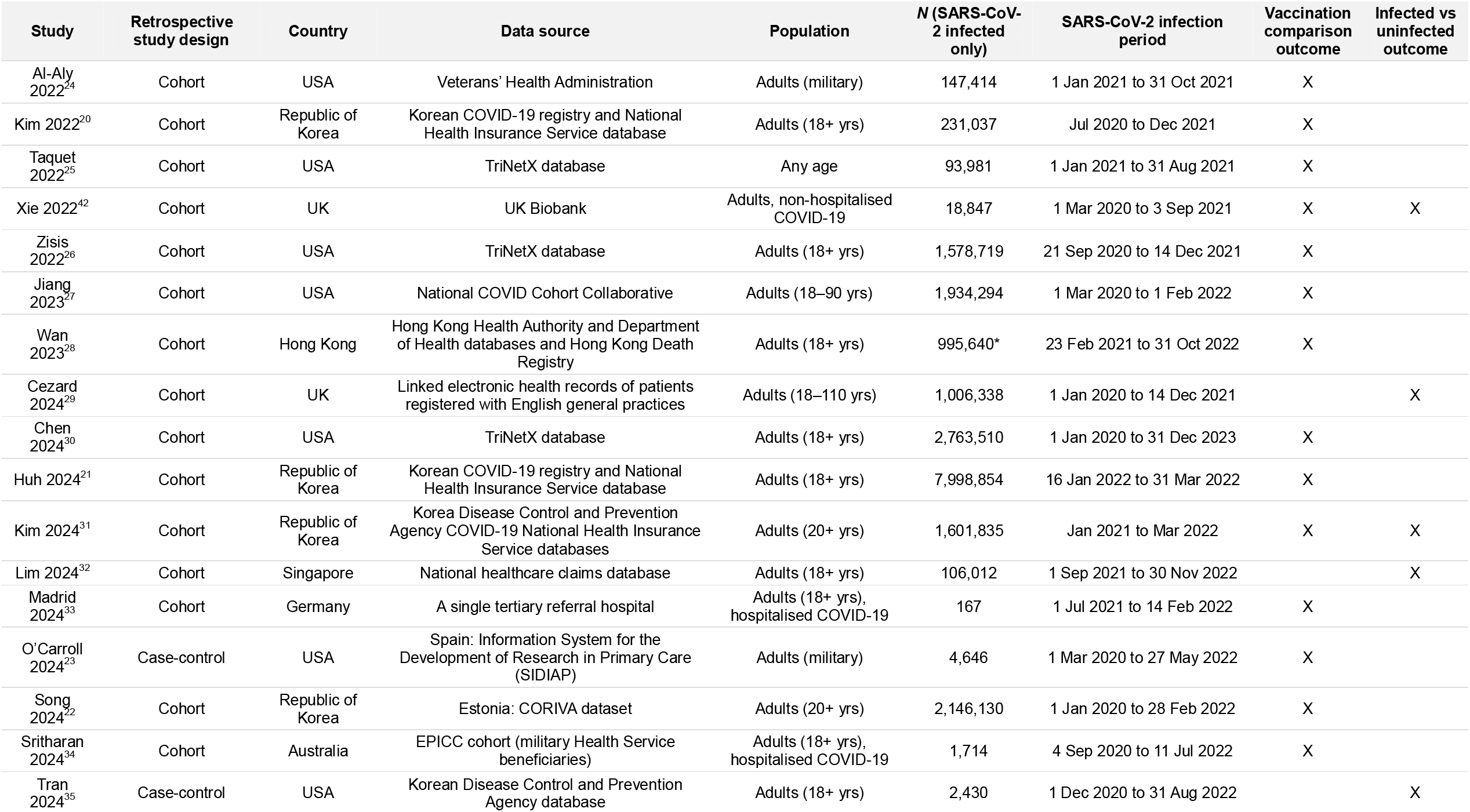
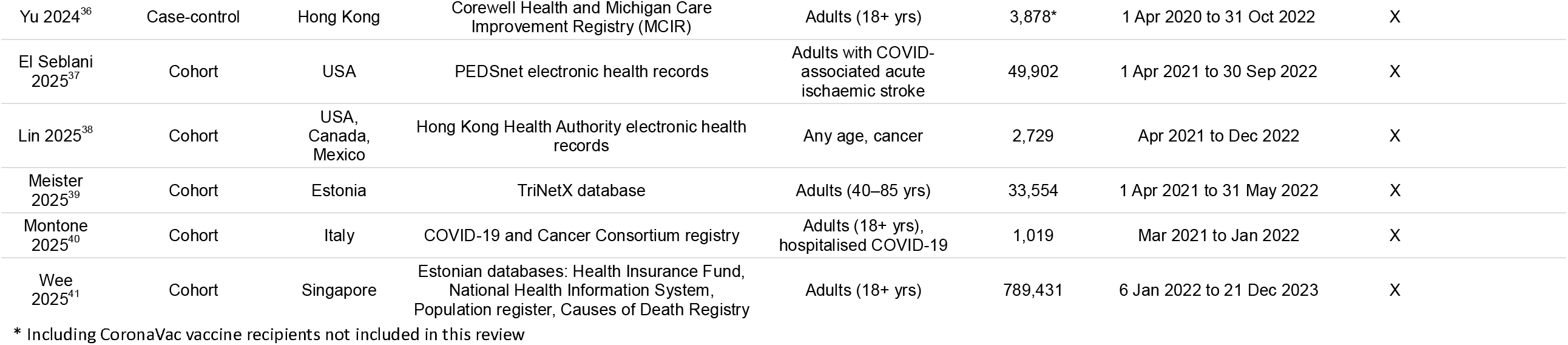
Characteristics of included studies.

### Search strategy

Literature searches of Embase, MEDLINE (ALL), and PubMed were conducted to identify studies published in peer-reviewed journals between 1 January 2021 and 11 September 2025. Search strategies were developed in accordance with the CRD guidance for undertaking reviews in health care^15^ and the Cochrane Handbook for Systematic Reviews of Interventions.^16^ Strategies combined controlled vocabulary terms (e.g. Medical Subject Headings [MeSH]) and free-text terms searched in titles and abstracts. Searches were structured using terms for COVID-19/SARS-CoV-2, named COVID-19 vaccines, and cardiovascular outcomes, and were combined with appropriate methodological filters, including filters for observational study designs. Searches were designed and executed by an experienced Information Specialist. The full search strategy is provided in Supplementary material online, Search strategies. In addition, reference lists of included original studies and relevant review articles were screened, and forward citation tracking of eligible publications was performed to identify additional studies.

### Study selection and data extraction

Studies were included if they met the following criteria:

1. reported the risk of cardiovascular, cerebrovascular or venous thromboembolic events after SARS-CoV-2 infection among individuals who had received COVID-19 vaccination compared with unvaccinated individuals or those who had received fewer vaccine doses, or compared monovalent and bivalent doses; or
2. reported the risk of cardiovascular, cerebrovascular or venous or thromboembolic events after SARS-CoV-2 infection compared with uninfected individuals, with results stratified by vaccination status.

Supplementary material online, Figure 1 illustrates the design of included studies.

**Figure 1.**
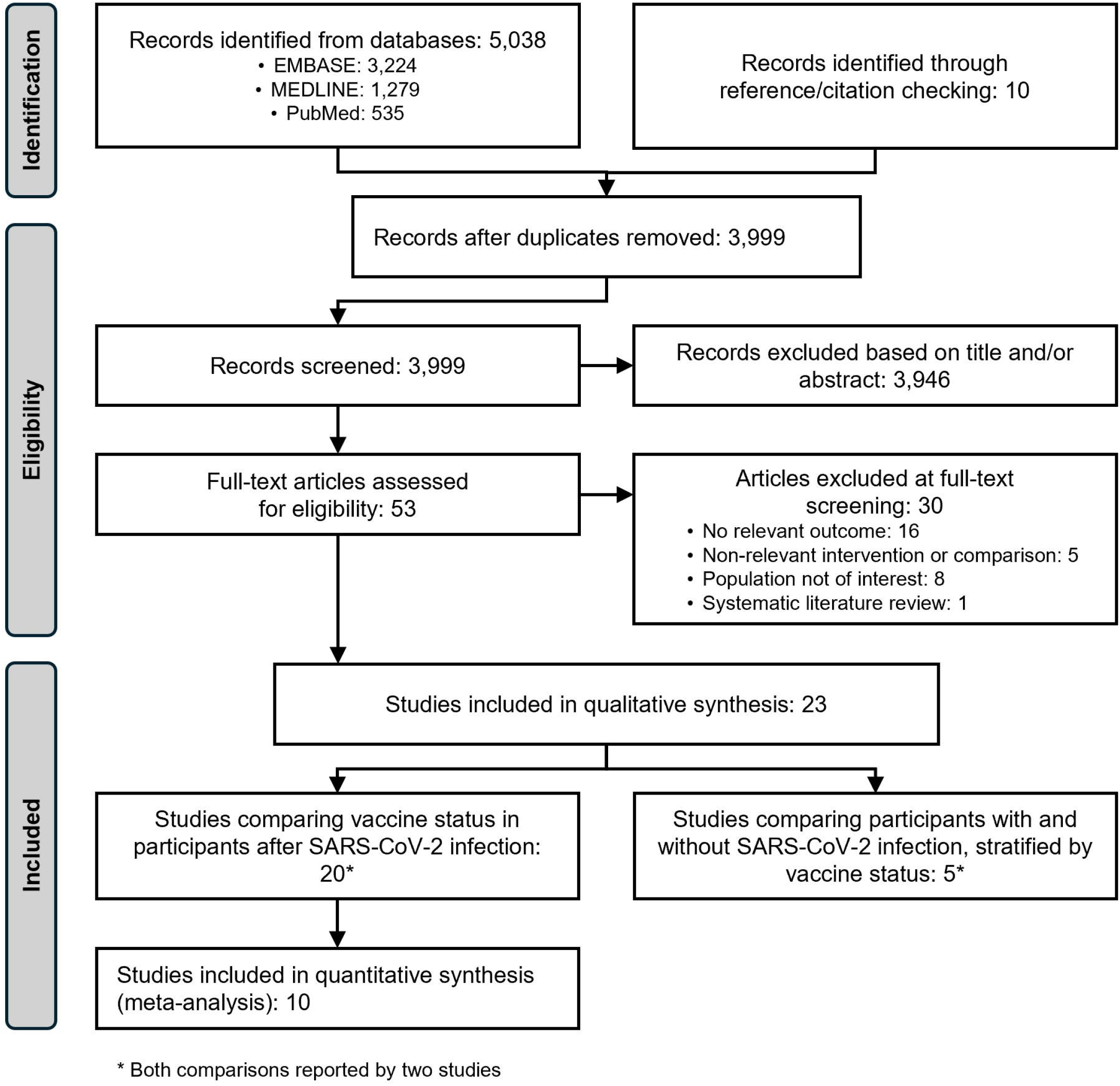
PRISMA flow diagram of literature search and study selection

Eligible vaccines were restricted to those authorised by the European Commission at the time of study conduct and included BNT162b2 (Pfizer–BioNTech), mRNA-1273 (Moderna), Ad26.COV2.S (Janssen), ChAdOx1-S (AstraZeneca), Nuvaxovid (Novavax), VLA2001 (Valneva), and VidPrevtyn Beta (Sanofi–GSK). Studies conducted in countries in which only these vaccines were administered were included even when vaccine type or manufacturer was not explicitly reported. Studies were also restricted to only those where vaccination occurred before the documented SARS-CoV-2 infection.

After deduplication, all records were independently screened by two reviewers (Z.M. and R.G.). Titles and abstracts were assessed for eligibility against the inclusion criteria, and records deemed potentially eligible underwent full-text review. Any disagreements were resolved by consensus.

Data extraction was performed using a predefined extraction form. A single reviewer extracted data from each included study, including study design, participant characteristics, vaccination and SARS-CoV-2–related characteristics, and outcomes of interest. All reported risk estimates were extracted across outcomes, subgroups, follow-up periods, vaccine doses, and vaccination comparisons. A second reviewer independently verified the accuracy and completeness of the extracted data. Only adjusted or weighted analyses were extracted. At least two attempts were made to contact the authors of publications with missing or unclear data.

### Quality assessment

Risk of bias was assessed independently by two reviewers (Z.M. and R.G.) using the Risk of Bias in Non-randomised Studies of Interventions (ROBINS-E) tool.^18^ Confounding factors assessed were age, sex, comorbidities, severity of SARS-CoV-2 infection, and socioeconomic factors.

The certainty of evidence was evaluated using the GRADE (Grading of Recommendations Assessment, Development and Evaluation) approach, which assesses five domains: risk of bias, inconsistency, indirectness, imprecision, and publication bias.^19^ Two reviewers (Z.M. and R.G.) independently assessed each domain for all outcomes.

Discrepancies between reviewers for both ROBINS-E and GRADE assessments were resolved through discussion to achieve consensus.

### Statistical analysis

All studies identified through the systematic review that reported risks of any outcomes after SARS-CoV-2 infection among individuals with one vaccination status compared with individuals with a different vaccination status were assessed for suitability for inclusion in pairwise meta-analyses (Supplementary material online, Meta-analysis feasibility assessment). Studies that reported the risk of outcomes after SARS-CoV-2 infection compared with uninfected individuals were not considered for meta-analyses, as they did not report a direct vaccination comparison. Meta-analyses were conducted for six outcomes — composite cardiovascular (cardiac) and cerebrovascular events, stroke, coronary syndromes, heart failure, arrhythmia, and venous thromboembolism — and restricted to studies comparing the risk of these outcomes post–SARS-CoV-2 infection in vaccinated individuals with that in unvaccinated individuals. Effect estimates were log-transformed, and study weights were calculated using the inverse-variance method (weight = 1/variance). Random-effects meta-analyses were performed using the DerSimonian and Laird method to estimate pooled hazard ratios (HRs) with corresponding 95% confidence intervals (CI).

Statistical heterogeneity was assessed using Cochran’s Q test, with statistical significance defined as p < 0.05, and quantified using the *I*^2^ statistic. Interpretation of *I*^2^ values followed thresholds recommended in the Cochrane Handbook for Systematic Reviews of Interventions.^16^ Publication bias was not formally assessed because all meta-analyses included fewer than ten studies.

When multiple post-infection time points or vaccine dose numbers were reported within a study, the time point or dose most comparable to those reported in other included studies was selected. For each study, the most fully adjusted effect estimates were used. Sensitivity analyses were conducted to assess the robustness of the findings and included leave-one-out analyses, comparisons using the lowest and highest vaccine dose numbers, shortest and longest post-infection risk periods, exclusion of studies with potential population overlap, and the addition, removal, or substitution of alternative outcome definitions. A prespecified subgroup analysis was performed for ischaemic and haemorrhagic stroke.

All statistical analyses were conducted using R version 4.1.1 (R Foundation for Statistical Computing) with the meta package.

## Results

### Systematic review of the literature

A total of 5,038 records were identified through database searches, with an additional ten records identified through reference list screening and citation tracking. Following title and abstract screening, 54 articles underwent full-text review for eligibility, of which 23 were included in the systematic review^6,20–41^ (Figure 1).

### Characteristics of included studies

The main characteristics of the included studies are displayed in Table 1, with additional details provided in Supplementary material online, Table 2.

**Table 2.**
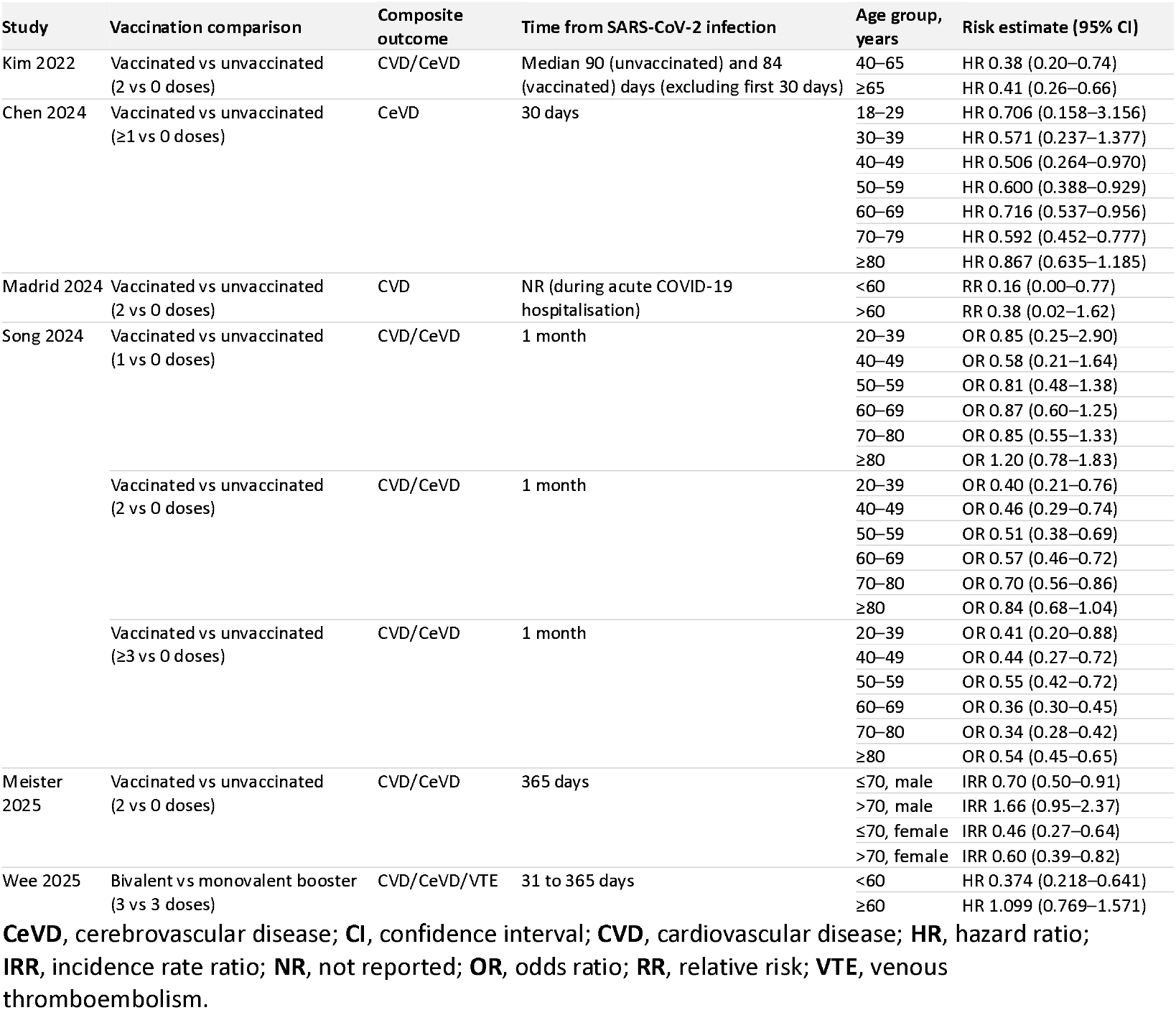
Risk of composite outcomes reported by different age groups.

Of the included 23 studies, 20 (18 cohort^6,20–22,24–28,30,31,33,34,37–41^ and two case-control^23,36^) assessed the association between vaccination and the risk of post-SARS-CoV-2 CVD/CeVD/VTE. Nineteen of these studies compared different number of vaccine doses (including no vaccination),^6,20–28,30,31,33,34,36–40^ and one study compared bivalent and monovalent booster doses.^41^ Five studies (four cohort and one case-control) compared the risk of CVD/CeVD/VTE events following SARS-CoV-2 infection versus uninfected individuals stratified by vaccination status.^6,29,31,32,35^

Twenty-one studies restricted inclusion to adults, while two did not report age-based inclusion criteria.^6,38^ Only individuals hospitalised for severe COVID-19 were included in three studies,^33,34,40^ and only non-hospitalised infections were included in one study.^6^ One study was restricted to individuals with COVID-19-associated acute ischaemic stroke,^37^ and one study included only individuals with cancer.^38^ Individuals with an outcome event prior to the study index date were excluded by 11 studies.^20–22,28–31,36,39,41,42^

Most studies included SARS-CoV-2 infections occurring in 2021 and 2022 (Supplementary material online, Figure 2); two studies included only Omicron variant infections,^21,41^ six included only pre-Omicron variant infections,^6,20,24–26,29^ and the remaining studies included infections spanning both periods.^22,23,27,28,30–40^ Vaccine platform and product were reported in 19 studies (Supplementary material online, Table 3). Four studies included mRNA-based (BNT162b2 and mRNA-1273) vaccines only,^32,35,38,41^ and two additional studies included both BNT162b2 and CoronaVac (whole inactivated virus vaccine) vaccination, with outcomes reported exclusively for individuals vaccinated with BNT162b2.^28,36^

**Figure 2.**
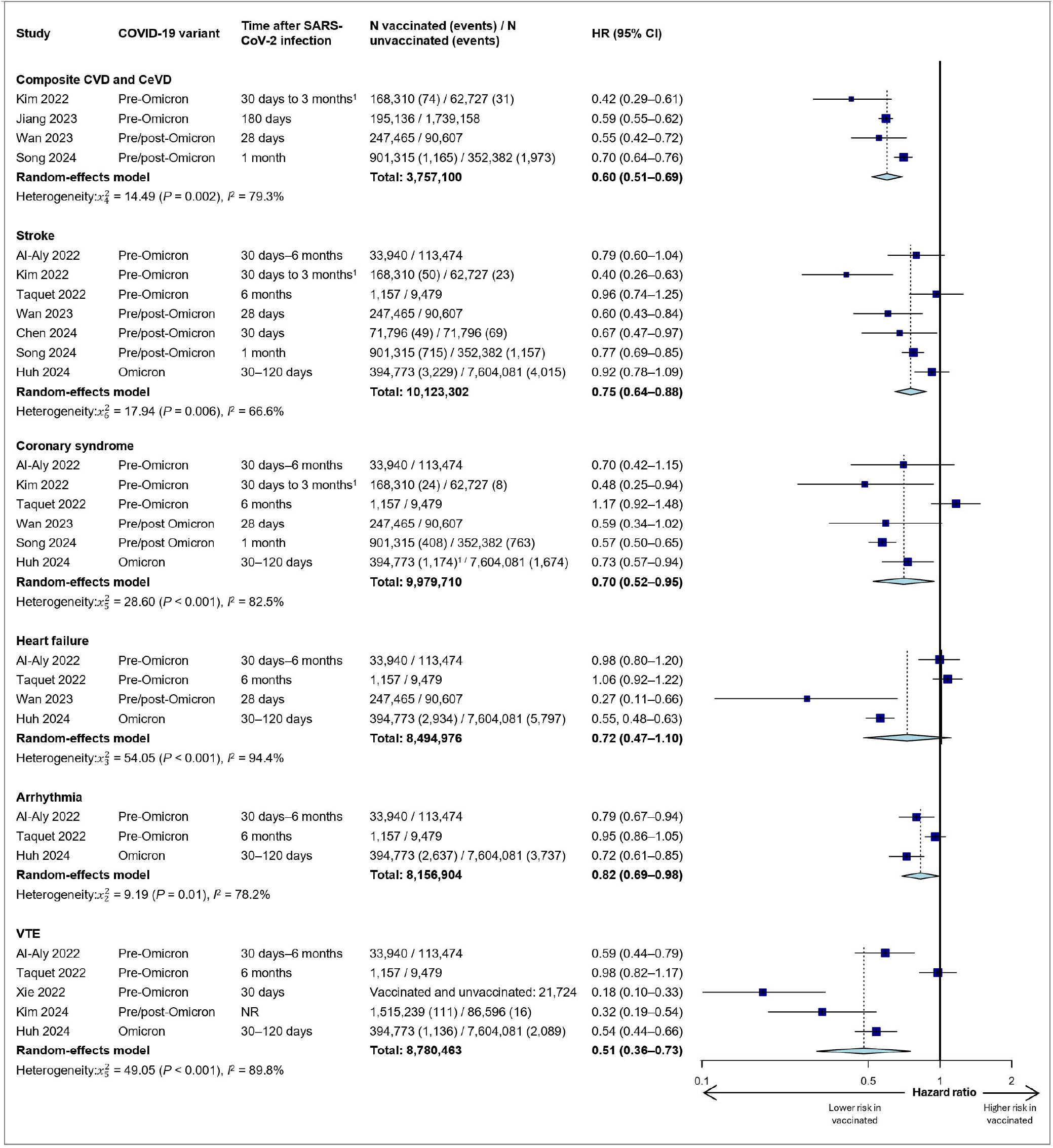
Pooled HRs (random-effects) of the risks of CVD/CeVD/VTE events after SARS-COV-2 infection in COVID-19-vaccinated individuals compared with unvaccinated individuals.

Risk of bias assessment using ROBINS-E classified 13 studies as having low risk of bias.^20–22,24,29–32,35,36,39,41,42^ Three were judged to have some concerns^25,28,37^ and five to have high risk of bias,^23,26,27,38,40^ due to residual confounding related to incomplete adjustment for COVID-19 severity and socioeconomic factors (Supplementary material online, Table 4). Two studies, both restricted to individuals hospitalised with severe COVID-19 were judged to be at very high risk of bias due to substantial missing or incomplete vaccination records and potential bias in outcome measurement, as events were ascertained during the index hospitalisation for acute infection.^33,34^

### Impact of COVID-19 vaccination on risk of CVD/CeVD/VTE events post-SARS-CoV-2 infection

Overall, the included studies indicated that pre-infection COVID-19 vaccination was associated with a reduced risk of CVD/CeVD/VTE events following SARS-CoV-2 infection. Significantly lower risks were observed across all assessed outcomes among fully vaccinated individuals (at least two vaccine doses) compared with unvaccinated individuals by 11 studies,^20,22,23,26,27,31,36–39,42^ while five reported significant risk reductions for a subset of outcomes.^21,24,28,30,33^ Three studies reported no significant association between COVID-19 vaccination and selected outcomes, including cardiac, stroke, hypertension, or venous thromboembolism events,^25^ composite cardiovascular and pulmonary embolism outcomes,^34^ or myocardial injury as assessed by high-sensitivity cardiac troponin I levels;^40^ two of these studies included a relatively small population of only individuals hospitalised for SARS-CoV-2 infection.^34,40^ Among 13 studies reporting composite outcomes, all but one^34^ demonstrated significantly lower composite risks in vaccinated compared with unvaccinated individuals (Supplementary material online, Figure 3). Increasing numbers of vaccine doses were associated with progressively lower risks of outcomes compared with no vaccination.^22,23,25,27,28,30,36^ Additionally, two studies reported lower risks among individuals receiving booster doses compared with full vaccination alone (Supplementary material online, Figure 4).^21,22^ A single study comparing the risk of CVD/CeVD/VTE events in individuals receiving either bivalent or monovalent mRNA boosters reported those with bivalent vaccination had a significantly lower risk of post-acute thrombotic disorders than those with monovalent vaccination, while bivalent vaccination associations with CVD/CeVD outcomes were directionally protective but largely not statistically significant over 31–365 days after infection.^41^

**Figure 3.**
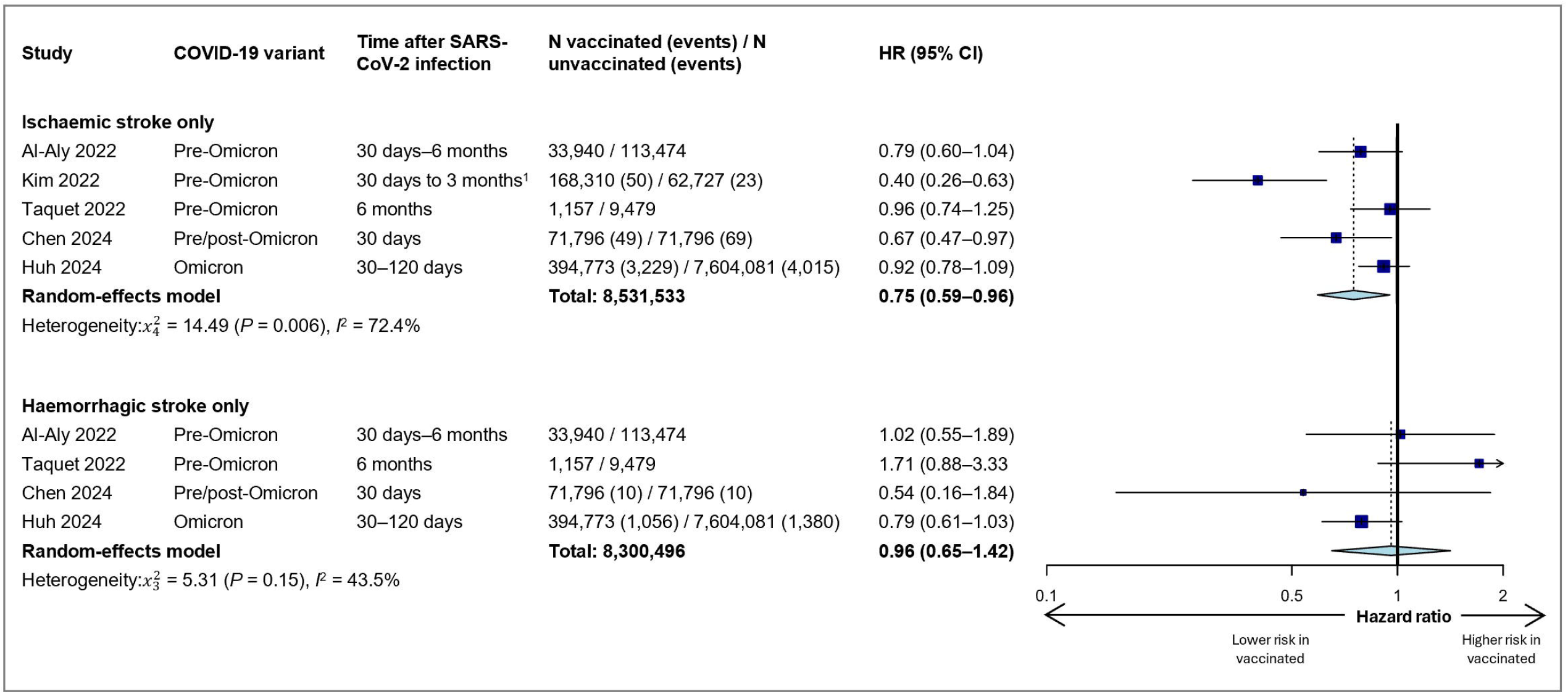
Pooled HRs (random-effects) of the risks of ischaemic stroke and haemorrhagic stroke after SARS-COV-2 infection in COVID-19-vaccinated individuals compared with unvaccinated individuals.

Outcome assessment periods ranged from three days to one year after infection; studies evaluating temporal patterns reported either attenuation of the protective association over time, with risk reductions still persisting at later follow-up,^22,28^ or broadly stable risk estimates across early and later post-infection periods.^26,30,37^ The impact of vaccination on risk of CVD/CeVD/VTE events was also reported by SARS-CoV-2 infection severity (five studies; mixed findings reported across studies^20,21,24,30(p202),39^) and variants (five studies; no substantial change in reduced risk across variants^23,30,33,37,38^), vaccine type (two studies; one reporting similar outcome risks for mRNA and non-mRNA vaccines^31^ and one reporting a significantly reduced risk of outcomes in individuals vaccinated with BNT162b2 but non-significant for mRNA-1273^37^) (Supplementary material online, Tables 6–8). Studies also reported risks by whether individuals had a history of the myocardial infarction or stroke outcome (one study; risks only significantly reduced in those with no outcome history^20^), by sex (five studies; similar risks reported for both sexes with the exception of one study reporting a significantly reduced risk only in females^20,22,30,39,41(p202)^), and by comorbidity status (five studies; similar risks reported across comorbidity statuses^20,22,30,33,41^) and immunocompromised status (two studies; one reporting similar risk to the entire study population^33^ and one reporting a significantly reduced risk only in non-immunocompromised people^24^) (Supplementary material online, Tables 9–12).

Most studies did not assess effect modification by age; however, several suggested potential age-related differences in vaccine-associated protection (Supplementary material online, Table 13). Associations between vaccination and composite CVD/CeVD/VTE outcomes only across different adult age groups were reported by six studies (Table 2). Four studies reported broadly similar risk estimates across age groups.^20,21,25,30^ In contrast, Madrid *et al*.^33^ reported significant risk reductions in younger individuals but not in those aged ≥ 60 years, and Meister *et al*.^39^ observed no protective association in men over 70 years, speculating that this may reflect a higher burden of pre-existing or undiagnosed cardiovascular disease in this group. Furthermore, Song *et al*.^22^ reported that, while odds ratios for CVD/CeVD following two vaccine doses increased with age, receipt of three or more doses was associated with lower odds ratios in middle-aged and older individuals. Montone *et al*.^40^ reported that full vaccination (two or more doses) was protective against myocardial injury in older patients (≥ 76 years) hospitalised for COVID-19. In contrast, among hospitalised patients aged 60 years or younger, vaccination was identified as an independent positive predictor of myocardial injury; however, the prevalence of this was still rare in this age group (3.8%). The authors suggested that these age-related differences may reflect distinct mechanisms of injury. In older individuals, myocardial injury may primarily relate to pre-existing cardiovascular disease or acute infection-related damage. In younger individuals, it may instead be associated with autoimmune mechanisms triggered by SARS-CoV-2 infection that could be amplified by previous COVID-19 immunisation.^40^

### Risk of CVD/CeVD/VTE events post-SARS-CoV-2 infection compared with no infection, stratified by vaccine status

Across five studies including adults only (one of which only included individuals with non-hospitalised COVID-19), risks of CVD/CeVD/VTE events were higher in individuals following SARS-CoV-2 infection compared with uninfected controls, with risks highest in the early post-infection period and declining over time; however, risks were consistently lower among vaccinated versus unvaccinated individuals (Supplementary material online, Table 14).^29,31,32(p20),35,42^ In two studies that stratified outcomes by infection severity, risks following severe or hospitalised COVID-19 were substantially higher than those following non-severe or non-hospitalised disease for both vaccinated and unvaccinated groups, although vaccinated individuals had lower risks regardless of severity strata.^29,31^

### Meta-analyses: impact of COVID-19 vaccination on risk of post-SARS-CoV-2 infection CVD/CeVD/VTE events

Meta-analyses evaluating risks of post-SARS-CoV-2 infection events in vaccinated versus unvaccinated individuals were conducted for six outcomes: composite CVD and CeVD outcomes, stroke, coronary syndrome, heart failure, arrhythmia, and venous thromboembolism (Supplementary material online, Table 15). A total of ten studies from the pre- and post-Omicron eras were included in these meta-analyses.

Figure 2 presents forest plots of study-specific and pooled HR estimates (random-effects). Vaccination was associated with significantly lowered pooled HRs of composite CVD and CeVD outcomes (HR 0.60, 95% CI 0.51–0.69; four studies), stroke (HR 0.75, 95% CI 0.64–0.88; seven studies), coronary syndrome (HR 0.70, 95% CI 0.52–0.95; six studies), arrhythmia (HR 0.82, 95% CI 0.69–0.98; three studies), and VTE (HR 0.51, 95% CI 0.36–0.73; five studies). Vaccination was associated with a non-significantly lowered pooled HR of heart failure (HR 0.72, 95% CI 0.47–1.10; four studies).

Between-study heterogeneity (the extent to which variability in effect estimates across included studies reflects real differences beyond that expected from sampling error alone) was substantial for the meta-analyses of composite outcomes (*I*^2^=79.3%), stroke (*I*^2^=66.6%), coronary syndrome (*I*^2^=82.5%), arrhythmia (*I*^2^=78.2%), and venous thromboembolism (*I*^2^=89.8%), and considerable for the meta-analysis of heart failure (*I*^2^=94.4%).

In leave-one-out sensitivity analyses, exclusion of individual studies did not alter the statistical significance for the composite outcomes, stroke, heart failure and venous thromboembolism analyses. Pooled estimates for coronary syndrome and arrhythmia were sensitive to removal of individual studies (Supplementary material online, Table 16). All additional sensitivity analyses showed no change in statistical significance, except for the lowest-dose sensitivity analyses for the stroke and arrhythmia meta-analyses, the highest-dose sensitivity analysis for the coronary syndrome meta-analysis, and the sensitivity analysis excluding myocardial infarction outcomes only (two studies) from the coronary syndrome meta-analysis (Supplementary material online, Table 17). Overall, these analyses indicate that the findings of the composite outcomes, stroke, heart failure, and VTE meta-analyses were robust to potentially influential analytical decisions, whereas results for coronary syndrome and arrhythmia were more sensitive to specific assumptions.

Stroke subgroup analyses demonstrated a significantly lower risk of ischaemic stroke among vaccinated individuals, whereas no significant association was observed for haemorrhagic stroke (Figure 3). The pooled HR for five studies reporting risk of ischaemic stroke was 0.75 (95% CI 0.59– 0.56), which aligns with the overall stroke analysis. In contrast, the pooled risk for four studies of haemorrhagic stroke was 0.96 (95% CI 0.65–1.42).

Certainty of evidence (GRADE) for composite CVD and CeVD, stroke, and VTE was low, primarily due to observational study designs, while evidence for coronary syndromes, arrhythmias, and heart failure was very low, owing to inconsistency (the extent to which results vary across studies) and imprecision (whether the effect estimate is sufficiently precise, reflected by the width of CIs) (Supplementary material online, Table 18). Although pooled estimates consistently favoured vaccination, variability in effect sizes and limited precision reduced confidence in the magnitude of these associations.

## Discussion

To the best of our knowledge, this is the first systematic review and meta-analysis to examine how COVID-19 vaccination affects the risk of cardiovascular, cerebrovascular, and venous thromboembolism events following SARS-CoV-2 infection. Our synthesis of 23 studies indicates that vaccination is associated with a reduced risk of these events, both in the acute infection period (up to four weeks post-infection) and in the months after infection. This association was observed across differing levels of COVID-19 severity and circulating SARS-CoV-2 variants, with evidence suggesting the risk is decreased further with increasing number of vaccinations. While further research is needed to investigate the impact in various demographics and high-risk populations, vaccination was consistently associated with a lower risk of CVD/CeVD/VTE events independent of sex, age groups, comorbidities, or immunocompromised status. Since the search date of this review, at least one additional study has been published reporting similar findings, with fully vaccinated individuals demonstrating a lower risk of CVD/CeVD/VTE events occurring at least 30 days after infection compared with unvaccinated infected individuals (adjusted HR 0.46, 95% CI 0.27–0.80).^43^

In our meta-analyses of ten studies including more than 13.6 million infected individuals, vaccination was associated with significantly lower pooled risks of post-infection stroke, coronary syndrome, arrhythmia, venous thromboembolism, and composite cardiovascular and cerebrovascular outcomes compared with no vaccination. In contrast, vaccination was not associated with a statistically significant reduction in post-infection heart failure risk in the pooled analysis of four studies. However, this analysis was limited by the small number of contributing studies and substantial heterogeneity in effect estimates. Notably, long-term cohort studies suggest that vaccination reduces the risk of post-infection heart failure compared with unvaccinated populations. A multinational cohort study conducted in the UK, Spain, and Estonia reported a significantly lower risk for up to one year.^44^ Similarly, a US study found that adults with pre-existing coronary artery disease or heart failure had significantly reduced risks of heart failure hospitalisation at both one and two years.^45^

In the stroke subgroup meta-analyses, vaccination was associated with a reduced risk of ischaemic stroke, while no directional association was observed for haemorrhagic stroke. This difference may be due to the distinct pathophysiological mechanisms underlying these stroke subtypes. Ischaemic stroke is primarily caused by vascular occlusion due to atherothrombosis, cardioembolism, or small-vessel disease. COVID-19 may elevate ischaemic stroke risk by promoting inflammation, endothelial dysfunction, and hypercoagulability, alongside cardiac injury and arrhythmias—particularly atrial fibrillation—as well as destabilisation of arterial plaques, thrombo-embolic events, and cerebral hypoperfusion.^46,47^

In contrast, haemorrhagic stroke generally arises from pathways less directly linked to SARS-CoV-2 infection such as vessel rupture associated with aneurysms or vascular malformations, with anticoagulant therapy acting as an additional contributing risk factor.^48^ Although ischaemic strokes may develop into haemorrhagic events, this is relatively rare, affecting roughly 2–7% of all cases.^49^ Due to the low incidence of haemorrhagic stroke after SARS-CoV-2 infection, the studies reviewed may lack sufficient power to identify changes in risk.^50,51^

### Mechanisms of vaccination impact on risk of cardiovascular disease

Cardiac injury following SARS-CoV-2 infection may arise from systemic inflammation, cytokine-mediated plaque instability and myocardial damage, activation of coagulation pathways, sympathetic activation, and stress-related cardiomyopathy.^52,53^ Thrombotic coronary events may arise following SARS-CoV-2 infection through similar mechanisms to ischaemic stroke. ^2,54^ Specifically, acute cardiovascular risk is amplified through infection-induced endothelial dysfunction and systemic thrombo-inflammation involving tissue factor and platelet activation which promotes venous and arterial micro- and macrothrombosis. Collectively, these processes may contribute to development of acute myocardial infarction, stroke, and heart failure.

The potential mechanisms by which vaccination reduces the risk of post-SARS-CoV-2 cardiovascular events are likely multifactorial. Aside from reducing the risk of SARS-CoV-2 infection itself, vaccination is associated with reduced disease severity^55,56^ and duration.^57^ Vaccination may protect against post-SARS-CoV-2 cardiovascular events by promoting rapid viral control,^58^ attenuating the infection-induced hyperinflammatory response^59–61^ and reducing the risk of COVID-19–associated coagulopathy and immunothrombosis.^2^ Through this, vaccination may reduce endothelial injury and lower the risk of downstream cardiac and thrombotic complications post-infection.

### Clinical implications

Although COVID-19 vaccines have been associated with a small increased risk of cardiovascular adverse events, mainly myocarditis and pericarditis following vaccination, these events are rare and occur significantly more frequently following SARS-CoV-2 infection.^62–64^ This review highlights the role of vaccination beyond prevention of SARS-CoV-2 infection, with evidence that they can protect against post-infection cardiovascular sequelae.

The risk of severe outcomes due to COVID-19 increases with age and multimorbidities.^65,66^ Vaccination is associated with a substantial reduction in the risk of hospitalisation,^67,68^ mortality,^69,70^ and post-infection multimorbidity^71^ in these populations. Consequently, COVID-19 vaccination is recommended for individuals with cardiovascular disease by major European and American clinical guidelines.^72–78^ People with pre-existing cardiovascular disease are also at increased risk of new cardiovascular events or exacerbations after SARS-CoV-2 infection.^79^ Compared with non-infected controls, patients hospitalised with COVID-19 who have pre-existing coronary artery disease have a significantly higher risk of new MACE (HR 1.58, 95% CI 1.38–1.80), myocardial infarction (HR 1.85, 95% CI 1.40–2.46), congestive heart failure (HR 1.51, 95% CI 1.27–1.79]), and stroke (HR 1.62, 95% CI 1.21–2.17]) over a follow-up of up to four years after infection.^80^ Those with a history of cardiovascular disease are also reported to have a six-fold higher risk of in-hospital death or MACE after COVID-19 hospitalisation (51% in those with previous cardiovascular disease compared with 15% in those without; OR 6.0 [95% CI 4.3–8.4]).^81^

Conditions such as diabetes and chronic kidney disease (CKD) increase susceptibility to severe COVID-19^82^ and are independently associated with elevated cardiovascular risk.^83,84^ Moreover, SARS-CoV-2 infection may worsen or precipitate new-onset diabetes or CKD,^86,87^ creating a bidirectional relationship that may further increase the likelihood of cardiovascular complications during and after infection.^87^ Vaccination may therefore confer particular benefit in individuals with these pre-existing conditions.

Herein we show that vaccination is consistently associated with a substantially reduced risk of new-onset cardiovascular events in populations without prior cardiovascular disease history. Of the 23 included studies, 11 excluded individuals with a history of cardiovascular disease; vaccination in all 11 studies was associated with a decreased risk of new-onset CVD/CeVD/VTE. Furthermore, in studies that assessed vaccination impact across different age groups, a significant reduction in these outcomes was observed in younger, lower-risk individuals. Taken together, these findings demonstrate that vaccination reduces post-infection cardiovascular risk across a broad population, extending beyond those with established cardiovascular disease or significant comorbidities. This protective effect was observed both during the pandemic era and, notably, in the current post-Omicron era.

Vaccination against respiratory pathogens such as influenza, respiratory syncytial virus, and pneumococcal disease has emerged as an important strategy for cardiovascular disease prevention,^88,89^ now endorsed by the European Society of Cardiology (ESC) as a fourth pillar of preventive cardiology alongside antihypertensives, lipid-lowering therapy, and glucose-lowering agents.^14^ COVID-19 vaccination has been incorporated into this framework, with recent ESC and American College of Cardiology (ACC) guidance recommending vaccination in high-risk groups, particularly those with established coronary artery disease, heart failure, and other cardiovascular comorbidities.^13,14^ The findings of the present systematic review and meta-analysis further support the role of COVID-19 vaccination in reducing the cardiovascular disease burden attributable to SARS-CoV-2, and support consideration of seasonal COVID-19 vaccination as part of comprehensive cardiovascular risk reduction.

### Review limitations

Several limitations of this review should be noted. Only observational studies were included due to the nature of the research question, and therefore causality cannot be inferred. Heterogeneity in study design, vaccinations, outcome definitions, follow-up duration, and control of confounding variables contributed to variability in effect estimates. While many studies included large population sizes, limited statistical power to detect rare outcomes or outcomes in subgroup analyses was frequently acknowledged by studies. Most studies relied on registry data, which may be subject to incomplete capture of exposures or outcomes, limited diagnostic validation, and incomplete information on socioeconomic and lifestyle factors.

Although all studies adjusted for potential confounders, these differed across studies, and not all controlled for key factors such as COVID-19 disease severity, socioeconomic status, or healthcare-seeking behaviour. A small number of studies acknowledged the potential for immortal time bias, noting that misclassification of pre-vaccination person-time, delayed exposure classification, or mismatched follow-up periods between comparison groups could have influenced effect estimates despite attempts to model vaccination as a time-varying exposure. In studies restricted to individuals with confirmed SARS-CoV-2 infection, conditioning on infection may also introduce collider bias, as infection risk is influenced by both vaccination status and baseline health characteristics, potentially distorting estimates of the association between vaccination and post-infection cardiovascular outcomes.

Limitations of the meta-analyses include the high between-study heterogeneity, and the low number of studies (less than five) included in the heart failure, arrhythmia, and composite outcomes analyses. The coronary syndromes and arrhythmia pooled estimates were sensitive to exclusion of individual studies. Subgroup analyses by vaccine type, demographic characteristics, or infection severity were not feasible due to insufficient comparable studies.

Only English-language studies were included. The search strategy required explicit naming of EMA-authorised COVID-19 vaccines to ensure exposure specificity and screening feasibility; consequently, relevant studies not including vaccine names in searchable fields may have been missed despite extensive hand-searching.

## Conclusions

COVID-19 vaccination is associated with a significant reduction of cardiovascular, cerebrovascular and venous thromboembolic events post-infection. The benefits of vaccination extend beyond preventing severe disease, offering protection against both pre-existing and new-onset cardiovascular events post-infection in adults. Given the elevated risk of cardiovascular complications, seasonal COVID-19 vaccines should be considered for individuals with known cardiovascular or cerebrovascular disease, as well as those with comorbidities, irrespective of age. These findings also support vaccination in older adults as a component of healthy ageing, and in younger adults who may have undiagnosed subclinical cardiovascular conditions exacerbated by infection.^90,91^

Taken together, these findings support COVID-19 vaccination as part of the fourth pillar of cardiovascular disease prevention, whereby seasonal vaccination may help reduce the cardiovascular burden of SARS-CoV-2 infection at a population level.

## Data Availability

All summary data generated during this study are presented in this published article or the supplemental material provided. Original data can be found in the published articles which can be retrieved from publicly available databases.

## Acknowledgements

The authors would like to thank Dr Amanda Gallagher (BioNTech SE) for medical writing support, including assisting authors with the development of the manuscript and editorial support, Ms Kate Misso (Maverex Ltd) for designing and running the search strategies, and Dr Daniel Mitchell (Maverex Ltd) for statistical support.

## Figure legends

**Figure 2**

Individual study estimates are shown as squares, with the size of the square proportional to study weight, and error bars indicating 95% CIs around the HR. The diamond represents the pooled effect size with its 95% CI.

^1^ Median 3 months starting from 30 days after infection.

CeVD, cerebrovascular disease; CVD, cardiovascular disease.

**Figure 3**

Individual study estimates are shown as squares, with the size of the square proportional to study weight, and error bars indicating 95% CIs around the HR. The diamond represents the pooled effect size with its 95% CI.

^1^ Median 3 months starting from 30 days after infection.

## References

1. Kang Y, Chen T, Mui D, et al. Cardiovascular manifestations and treatment considerations in COVID-19. Heart. 2020;106(15):1132–1141. doi:10.1136/heartjnl-2020-317056

2. Raman B, Bluemke DA, Lüscher TF, Neubauer S. Long COVID: post-acute sequelae of COVID-19 with a cardiovascular focus. Eur Heart J. 2022;43(11):1157–1172. doi:10.1093/eurheartj/ehac031

3. Kole C, Stefanou E, Karvelas N, Schizas D, Toutouzas KP. Acute and Post-Acute COVID-19 Cardiovascular Complications: A Comprehensive Review. Cardiovasc Drugs Ther. 2024;38(5):1017–1032. doi:10.1007/s10557-023-07465-w

4. Bikdeli B, Madhavan MV, Jimenez D, et al. COVID-19 and Thrombotic or Thromboembolic Disease: Implications for Prevention, Antithrombotic Therapy, and Follow-Up. JACC. 2020;75(23):2950–2973. doi:10.1016/j.jacc.2020.04.031

5. Romero Starke K, Kaboth P, Rath N, et al. Cardiovascular disease risk after a SARS-CoV-2 infection: A systematic review and meta-analysis. Journal of Infection. 2024;89(3):106215. doi:10.1016/j.jinf.2024.106215

6. Xie Y, Xu E, Bowe B, Al-Aly Z. Long-term cardiovascular outcomes of COVID-19. Nat Med. 2022;28(3):583–590. doi:10.1038/s41591-022-01689-3

7. Spetz M, Natt och Dag Y, Li H, Nyberg F, Rosvall M. Covid-19 and cardiovascular disease in a total population-study of long-term effects, social factors and Covid-19-vaccination. Nat Commun. 2025;16(1):10115. doi:10.1038/s41467-025-66270-1

8. Battistoni A, Volpe M, Morisco C, et al. Persistent increase of cardiovascular and cerebrovascular events in COVID-19 patients: a 3-year population-based analysis. Cardiovasc Res. 2024;120(6):623–629. doi:10.1093/cvr/cvae049

9. Al-Aly Z. Long COVID and its cardiovascular implications: a call to action. Eur Heart J. 2023;44(47):5001–5003. doi:10.1093/eurheartj/ehad723

10. Sliwa K, Singh K, Nikhare K, et al. Long COVID Syndrome, Mortality and Morbidity in Patients Hospitalized with COVID-19 From 16 Countries: The World Heart Federation Global COVID-19 Study. Glob Heart. 20(1):66. doi:10.5334/gh.1452

11. Mohammadi M, Kianifard N, Fazlzadeh A, et al. Association between influenza infection and cardiovascular diseases: A systematic review and meta-analysis. JRSM Cardiovasc Dis. 2025;14:20480040251407014. doi:10.1177/20480040251407014

12. Behrouzi B, Bhatt DL, Cannon CP, et al. Association of Influenza Vaccination With Cardiovascular Risk: A Meta-analysis. JAMA Netw Open. 2022;5(4):e228873. doi:10.1001/jamanetworkopen.2022.8873

13. Heidenreich P, Bhatt A, Nazir NT, Schaffner W, Vardeny O. 2025 Concise Clinical Guidance: An ACC Expert Consensus Statement on Adult Immunizations as Part of Cardiovascular Care. JACC. 2025;86(21):2085–2098. doi:10.1016/j.jacc.2025.07.003

14. Heidecker B, Libby P, Vassiliou VS, et al. Vaccination as a new form of cardiovascular prevention: a European Society of Cardiology clinical consensus statement: With the contribution of the European Association of Preventive Cardiology (EAPC), the Association for Acute CardioVascular Care (ACVC), and the Heart Failure Association (HFA) of the ESC. Eur Heart J. 2025;46(36):3518–3531. doi:10.1093/eurheartj/ehaf384

15. Centre for Reviews and Dissemination, University of York. CRD’s guidance on systematic reviews. Published online 2009.

16. Higgins JPT, Thomas J, Chandler J, Cumpston M, Li T, Page MJ. Cochrane Handbook for Systematic Reviews of Interventions. version 6.5 (updated August 2024). Cochrane; 2024. www.cochrane.org/handbook

17. Page MJ, McKenzie JE, Bossuyt PM, et al. The PRISMA 2020 statement: an updated guideline for reporting systematic reviews. BMJ. 2021;372:n71. doi:10.1136/bmj.n71

18. Higgins JPT, Morgan RL, Rooney AA, et al. A tool to assess risk of bias in non-randomized follow-up studies of exposure effects (ROBINS-E). Environment International. 2024;186:108602. doi:10.1016/j.envint.2024.108602

19. Guyatt GH, Oxman AD, Vist GE, et al. GRADE: an emerging consensus on rating quality of evidence and strength of recommendations. BMJ. 2008;336(7650):924–926. doi:10.1136/bmj.39489.470347.AD

20. Kim YE, Huh K, Park YJ, Peck KR, Jung J. Association Between Vaccination and Acute Myocardial Infarction and Ischemic Stroke After COVID-19 Infection. JAMA. 2022;328(9):887–889. doi:10.1001/jama.2022.12992

21. Huh K, Kim YE, Bae GH, et al. Vaccination and the risk of post-acute sequelae after COVID-19 in the Omicron-predominant period. Clinical Microbiology and Infection. 2024;30(5):666–673. doi:10.1016/j.cmi.2024.01.028

22. Song J, Choi S, Jeong S, et al. Protective effect of vaccination on the risk of cardiovascular disease after SARS-CoV-2 infection. Clin Res Cardiol. 2024;113(2):235–245. doi:10.1007/s00392-023-02271-8

23. O’Carroll A, Richard SA, Byrne C, et al. Estimating the Effect of Coronavirus Disease 2019 (COVID-19) Vaccination and Infection Variant on Post-COVID-19 Venous Thrombosis or Embolism Risk. Open Forum Infect Dis. 2024;11(11):ofae557. doi:10.1093/ofid/ofae557

24. Al-Aly Z, Bowe B, Xie Y. Long COVID after breakthrough SARS-CoV-2 infection. Nat Med. 2022;28(7):7. doi:10.1038/s41591-022-01840-0

25. Taquet M, Dercon Q, Harrison PJ. Six-month sequelae of post-vaccination SARS-CoV-2 infection: A retrospective cohort study of 10,024 breakthrough infections. Brain, Behavior, and Immunity. 2022;103:154–162. doi:10.1016/j.bbi.2022.04.013

26. Zisis SN, Durieux JC, Mouchati C, Perez JA, McComsey GA. The Protective Effect of Coronavirus Disease 2019 (COVID-19) Vaccination on Postacute Sequelae of COVID-19: A Multicenter Study From a Large National Health Research Network. Open Forum Infect Dis. 2022;9(7):ofac228. doi:10.1093/ofid/ofac228

27. Jiang J, Chan L, Kauffman J, et al. Impact of Vaccination on Major Adverse Cardiovascular Events in Patients With COVID-19 Infection. JACC. 2023;81(9):928–930. doi:10.1016/j.jacc.2022.12.006

28. Wan EYF, Mok AHY, Yan VKC, et al. Association between BNT162b2 and CoronaVac vaccination and risk of CVD and mortality after COVID-19 infection: A population-based cohort study. Cell Reports Medicine. 2023;4(10):101195. doi:10.1016/j.xcrm.2023.101195

29. Cezard GI, Denholm RE, Knight R, et al. Impact of vaccination on the association of COVID-19 with cardiovascular diseases: An OpenSAFELY cohort study. Nat Commun. 2024;15(1):2173. doi:10.1038/s41467-024-46497-0

30. Chen SY, Hsieh TYJ, Hung YM, et al. Prior COVID-19 vaccination and reduced risk of cerebrovascular diseases among COVID-19 survivors. Journal of Medical Virology. 2024;96(5):e29648. doi:10.1002/jmv.29648

31. Kim HJ, Jeong S, Song J, et al. Risk of pulmonary embolism and deep vein thrombosis following COVID-19: a nationwide cohort study. MedComm. 2024;5(7):e655. doi:10.1002/mco2.655

32. Lim JT, Liang En W, Tay AT, et al. Long-term Cardiovascular, Cerebrovascular, and Other Thrombotic Complications in COVID-19 Survivors: A Retrospective Cohort Study. Clin Infect Dis. 2024;78(1):70–79. doi:10.1093/cid/ciad469

33. Madrid J, Agarwal P, Müller-Peltzer K, et al. Cardioprotective effects of vaccination in hospitalized patients with COVID-19. Clin Exp Med. 2024;24(1):103. doi:10.1007/s10238-024-01367-3

34. Sritharan HP, Bhatia KS, van Gaal W, Kritharides L, Chow CK, Bhindi R. Cardiovascular outcomes for people hospitalised with COVID-19 in Australia, and the effect of vaccination: an observational cohort study. Medical Journal of Australia. 2024;220(10):517–522. doi:10.5694/mja2.52307

35. Tran HNQ, Risk M, Nair GB, Zhao L. Risk benefit analysis to evaluate risk of thromboembolic events after mRNA COVID-19 vaccination and COVID-19. npj Vaccines. 2024;9(1):166. doi:10.1038/s41541-024-00960-7

36. Yu Q, Fan M, Lin CJ, et al. Risk factors for long-term cardiovascular post-acute sequelae of COVID-19 infection: A nested case-control study in Hong Kong. npj Cardiovasc Health. 2024;1(1):10. doi:10.1038/s44325-024-00011-z

37. El Seblani N, Gorenflo MP, Ortega-Gutierrez S, Reichwein RK, Xu R, Nagaraja N. Vaccination against COVID-19 and Outcomes in Patients with COVID-19 Infection and Stroke. npj Vaccines. 2025;10(1):133. doi:10.1038/s41541-025-01158-1

38. Lin EPY, Hsu CY, Mishra S, et al. Associations of COVID-19 vaccination with risks for post-infectious cardiovascular complications: an international cohort study in cancer patients with SARS-CoV-2 infection. The Lancet Regional Health – Americas. 2025;44. doi:10.1016/j.lana.2025.101038

39. Meister T, Maiväli Ü, Tenson K, et al. Dynamic effects of COVID-19 vaccination on major acute cardiovascular events and mortality following SARS-CoV-2 infection in a target trial emulation study. Sci Rep. 2025;15(1):27530. doi:10.1038/s41598-025-13043-x

40. Montone RA, Rinaldi R, Masciocchi C, et al. Vaccines and myocardial injury in patients hospitalized for COVID-19 infection: the CardioCOVID-Gemelli study. Eur Heart J Qual Care Clin Outcomes. 2025;11(1):59–67. doi:10.1093/ehjqcco/qcae016

41. Wee LE, Lim JT, Goel M, et al. Bivalent Boosters and Risk of Postacute Sequelae Following Vaccine-Breakthrough SARS-CoV-2 Omicron Infection: A Cohort Study. Clin Infect Dis. 2025;80(3):520–528. doi:10.1093/cid/ciae598

42. Xie J, Prats-Uribe A, Feng Q, et al. Clinical and Genetic Risk Factors for Acute Incident Venous Thromboembolism in Ambulatory Patients With COVID-19. JAMA Intern Med. 2022;182(10):1063–1070. doi:10.1001/jamainternmed.2022.3858

43. Soares P, Ruivinho C, Silva J, et al. Long-term cardiovascular events in individuals hospitalised with COVID-19: a retrospective cohort. BMC Infect Dis. 2025;25:1525. doi:10.1186/s12879-025-11762-0

44. Mercadé-Besora N, Li X, Kolde R, et al. The role of COVID-19 vaccines in preventing post-COVID-19 thromboembolic and cardiovascular complications. Heart. 2024;110(9):635–643. doi:10.1136/heartjnl-2023-323483

45. Akbar UA, Thyagaturu H, Taha A, et al. COVID-19 Vaccination and Cardiovascular Outcomes in Older Adults With Coronary Artery Disease and Heart Failure: Insights From a Large Propensity-Matched Cohort Study. Journal of the American Heart Association. 2025;14(21):e044546. doi:10.1161/JAHA.125.044546

46. Zhang S, Zhang J, Wang C, et al. COVID-19 and ischemic stroke: Mechanisms of hypercoagulability (Review). Int J Mol Med. 2021;47(3):21. doi:10.3892/ijmm.2021.4854

47. Aghayari Sheikh Neshin S, Shahjouei S, Koza E, et al. Stroke in SARS-CoV-2 Infection: A Pictorial Overview of the Pathoetiology. Front Cardiovasc Med. 2021;8. doi:10.3389/fcvm.2021.649922

48. Smith SD, Eskey CJ. Hemorrhagic Stroke. Radiologic Clinics. 2011;49(1):27–45. doi:10.1016/j.rcl.2010.07.011

49. Spronk E, Sykes G, Falcione S, et al. Hemorrhagic Transformation in Ischemic Stroke and the Role of Inflammation. Front Neurol. 2021;12. doi:10.3389/fneur.2021.661955

50. Siow I, Lee KS, Zhang JJY, Saffari SE, Ng A, Young B. Stroke as a Neurological Complication of COVID-19: A Systematic Review and Meta-Analysis of Incidence, Outcomes and Predictors. Journal of Stroke and Cerebrovascular Diseases. 2021;30(3):105549. doi:10.1016/j.jstrokecerebrovasdis.2020.105549

51. Nannoni S, de Groot R, Bell S, Markus HS. Stroke in COVID-19: A systematic review and metaanalysis. International Journal of Stroke. 2021;16(2):137–149. doi:10.1177/1747493020972922

52. Clerkin KJ, Fried JA, Raikhelkar J, et al. COVID-19 and Cardiovascular Disease. Circulation. 2020;141(20):1648–1655. doi:10.1161/CIRCULATIONAHA.120.046941

53. Helms J, Combes A, Aissaoui N. Cardiac injury in COVID-19. Intensive Care Med. 2022;48(1):111–113. doi:10.1007/s00134-021-06555-3

54. Conway EM, Mackman N, Warren RQ, et al. Understanding COVID-19-associated coagulopathy. Nat Rev Immunol. 2022;22(10):639–649. doi:10.1038/s41577-022-00762-9

55. Mohammed I, Nauman A, Paul P, et al. The efficacy and effectiveness of the COVID-19 vaccines in reducing infection, severity, hospitalization, and mortality: a systematic review. Hum Vaccin Immunother. 2022;18(1):2027160. doi:10.1080/21645515.2022.2027160

56. Katz MA, Cohuet S, Bino S, et al. COVID-19 vaccine effectiveness against SARS-CoV-2-confirmed hospitalisation in the eastern part of the WHO European Region (2022–2023): a test-negative case-control study from the EuroSAVE network. The Lancet Regional Health – Europe. 2024;47. doi:10.1016/j.lanepe.2024.101095

57. Ronchini C, Gandini S, Pasqualato S, et al. Lower probability and shorter duration of infections after COVID-19 vaccine correlate with anti-SARS-CoV-2 circulating IgGs. PLOS ONE. 2022;17(1):e0263014. doi:10.1371/journal.pone.0263014

58. Puhach O, Adea K, Hulo N, et al. Infectious viral load in unvaccinated and vaccinated individuals infected with ancestral, Delta or Omicron SARS-CoV-2. Nat Med. 2022;28(7):1491–1500. doi:10.1038/s41591-022-01816-0

59. Chan L, Pinedo K, Stabile MA, et al. Prior vaccination prevents overactivation of innate immune responses during COVID-19 breakthrough infection. Science Translational Medicine. 2025;17(783):eadq1086. doi:10.1126/scitranslmed.adq1086

60. Painter MM, Johnston TS, Lundgreen KA, et al. Prior vaccination enhances immune responses during SARS-CoV-2 breakthrough infection with early activation of memory T cells followed by production of potent neutralizing antibodies. bioRxiv. Published online February 6, 2023:2023.02.05.527215. doi:10.1101/2023.02.05.527215

61. Zhu X, Gebo KA, Abraham AG, et al. Dynamics of inflammatory responses after SARS-CoV-2 infection by vaccination status in the USA: a prospective cohort study. Lancet Microbe. 2023;4(9):e692–e703. doi:10.1016/S2666-5247(23)00171-4

62. Ishisaka Y, Watanabe A, Aikawa T, et al. Overview of SARS-CoV-2 infection and vaccine associated myocarditis compared to non-COVID-19-associated myocarditis: A systematic review and metaanalysis. International Journal of Cardiology. 2024;395:131401. doi:10.1016/j.ijcard.2023.131401

63. Klamer TA, Linschoten M, Asselbergs FW. The benefit of vaccination against COVID-19 outweighs the potential risk of myocarditis and pericarditis. Neth Heart J. 2022;30(4):190–197. doi:10.1007/s12471-022-01677-9

64. Cotet IG, Mateescu DM, Ilie AC, et al. Systematic Review and Meta-Analysis of Myocarditis Prevalence and Diagnostics in COVID-19:Acute, Post-COVID, and MIS-C (2020–2025). Journal of Clinical Medicine. 2025;14(19):7008. doi:10.3390/jcm14197008

65. Makovski TT, Ghattas J, Monnier-Besnard S, et al. Multimorbidity and frailty are associated with poorer SARS-CoV-2-related outcomes: systematic review of population-based studies. Aging Clin Exp Res. 2024;36(1):40. doi:10.1007/s40520-023-02685-4

66. Starke KR, Reissig D, Petereit-Haack G, Schmauder S, Nienhaus A, Seidler A. The isolated effect of age on the risk of COVID-19 severe outcomes: a systematic review with meta-analysis. BMJ Glob Health. 2021;6(12). doi:10.1136/bmjgh-2021-006434

67. Harrison C, Frain S, Jalalinajafabadi F, Williams SG, Keavney B. The impact of COVID-19 vaccination on patients with congenital heart disease in England: a case-control study. Heart. 2024;110(23):1372–1380. doi:10.1136/heartjnl-2024-324470

68. Parenica J, Jarkovsky J, Benesova K, et al. COVID-19 vaccination decrease the risk of Intensive Care Unit hospitalisation in heart failure patients even in the time of Omicron variant: population based study. Eur Heart J. 2022;43(Supplement_2):ehac544.1077. doi:10.1093/eurheartj/ehac544.1077

69. Brodersen KD, Ulrichsen SP, Sørup S, et al. Recent COVID-19 and mortality after myocardial infarction: a Danish nationwide cohort study. Eur J Prev Cardiol. Published online August 28, 2025:zwaf538. doi:10.1093/eurjpc/zwaf538

70. Patel S, Ballout M, Khan S, et al. Patients With ST-Segment Elevation Myocardial Infarction and Cerebrovascular Accidents: Impact of COVID-19 Vaccination on Mortality. Cardiology Research. 2024;15(4):275. doi:10.14740/cr.v15i4.1688

71. Liu B, Song S, Liu W, et al. Post-COVID-19 multimorbidity incidence by prior vaccination status in people with a pre-existing comorbidity: A population-based cohort study. Journal of Infection. 2025;91(3). doi:10.1016/j.jinf.2025.106597

72. Arbelo E, Protonotarios A, Gimeno JR, et al. 2023 ESC Guidelines for the management of cardiomyopathies: Developed by the task force on the management of cardiomyopathies of the European Society of Cardiology (ESC). Eur Heart J. 2023;44(37):3503–3626. doi:10.1093/eurheartj/ehad194

73. Heidenreich PA, Bozkurt B, Aguilar D, et al. 2022 AHA/ACC/HFSA Guideline for the Management of Heart Failure. JACC. 2022;79(17):e263–e421. doi:10.1016/j.jacc.2021.12.012

74. McDonagh TA, Metra M, Adamo M, et al. 2021 ESC Guidelines for the diagnosis and treatment of acute and chronic heart failure: Developed by the Task Force for the diagnosis and treatment of acute and chronic heart failure of the European Society of Cardiology (ESC) With the special contribution of the Heart Failure Association (HFA) of the ESC. Eur Heart J. 2021;42(36):3599–3726. doi:10.1093/eurheartj/ehab368

75. Byrne RA, Rossello X, Coughlan JJ, et al. 2023 ESC Guidelines for the management of acute coronary syndromes: Developed by the task force on the management of acute coronary syndromes of the European Society of Cardiology (ESC). Eur Heart J Acute Cardiovasc Care. 2024;13(1):55–161. doi:10.1093/ehjacc/zuad107

76. Vrints C, Andreotti F, Koskinas KC, et al. 2024 ESC Guidelines for the management of chronic coronary syndromes: Developed by the task force for the management of chronic coronary syndromes of the European Society of Cardiology (ESC) Endorsed by the European Association for Cardio-Thoracic Surgery (EACTS). Eur Heart J. 2024;45(36):3415–3537. doi:10.1093/eurheartj/ehae177

77. Virani SS, Newby LK, Arnold SV, et al. 2023 AHA/ACC/ACCP/ASPC/NLA/PCNA Guideline for the Management of Patients With Chronic Coronary Disease. JACC. 2023;82(9):833–955. doi:10.1016/j.jacc.2023.04.003

78. Humbert M, Kovacs G, Hoeper MM, et al. 2022 ESC/ERS Guidelines for the diagnosis and treatment of pulmonary hypertension: Developed by the task force for the diagnosis and treatment of pulmonary hypertension of the European Society of Cardiology (ESC) and the European Respiratory Society (ERS). Endorsed by the International Society for Heart and Lung Transplantation (ISHLT) and the European Reference Network on rare respiratory diseases (ERN-LUNG). Eur Heart J. 2022;43(38):3618–3731. doi:10.1093/eurheartj/ehac237

79. Denegri A, Dall’Ospedale V, Covani M, Pruc M, Szarpak L, Niccoli G. Cardiovascular Complications of COVID-19 Disease: A Narrative Review. Diseases. 2025;13(8):252. doi:10.3390/diseases13080252

80. Hadidchi R, Lee P, Qiu S, Changela S, Henry S, Duong TQ. Long-term outcomes of patients with pre-existing coronary artery disease after SARS-CoV-2 infection. eBioMedicine. 2025;116. doi:10.1016/j.ebiom.2025.105778

81. Tessitore E, Carballo D, Poncet A, et al. Mortality and high risk of major adverse events in patients with COVID-19 and history of cardiovascular disease. Open Heart. 2021;8(1):e001526. doi:10.1136/openhrt-2020-001526

82. Chapman A, Barouch DH, Lip GYH, et al. Risk of severe outcomes from COVID-19 in comorbid populations in the Omicron era: A systematic review and meta-analysis. International Journal of Infectious Diseases. 2025;158:107958. doi:10.1016/j.ijid.2025.107958

83. Dal Canto E, Ceriello A, Rydén L, et al. Diabetes as a cardiovascular risk factor: An overview of global trends of macro and micro vascular complications. Eur J Prev Cardiol. 2019;26(2_suppl):25–32. doi:10.1177/2047487319878371

84. Marx-Schütt K, Cherney DZI, Jankowski J, Matsushita K, Nardone M, Marx N. Cardiovascular disease in chronic kidney disease. Eur Heart J. 2025;46(23):2148–2160. doi:10.1093/eurheartj/ehaf167

85. Schiffl H, Lang SM. Long-term interplay between COVID-19 and chronic kidney disease. Int Urol Nephrol. 2023;55(8):1977–1984. doi:10.1007/s11255-023-03528-x

86. Kazakou P, Lambadiari V, Ikonomidis I, et al. Diabetes and COVID-19; A Bidirectional Interplay. Front Endocrinol. 2022;13. doi:10.3389/fendo.2022.780663

87. Viswanathan V, Puvvula A, Jamthikar AD, et al. Bidirectional link between diabetes mellitus and coronavirus disease 2019 leading to cardiovascular disease: A narrative review. World J Diabetes. 2021;12(3):215–237. doi:10.4239/wjd.v12.i3.215

88. Fröbert O, Cajander S, Udell JA. The ideal vaccine to prevent cardiovascular disease. Eur Heart J. 2023;44(7):621–623. doi:10.1093/eurheartj/ehac826

89. Rademacher J, Therre M, Hinze CA, Buder F, Böhm M, Welte T. Association of respiratory infections and the impact of vaccinations on cardiovascular diseases. Eur J Prev Cardiol. 2024;31(7):877–888. doi:10.1093/eurjpc/zwae016

90. Bergström G, Persson M, Adiels M, et al. Prevalence of Subclinical Coronary Artery Atherosclerosis in the General Population. Circulation. 2021;144(12):916–929. doi:10.1161/CIRCULATIONAHA.121.055340

91. Lindberg P, Wiqvist S, Juszczyk M, et al. Long COVID and risk of incident cardiovascular disease: a prospective cohort study using the Multimorbidity Integrated Registry Across Care Levels in Stockholm (MIRACLE-S) cohort. eClinicalMedicine. 2026;0(0). doi:10.1016/j.eclinm.2026.103846

