## Supplemental for "COVID-19 vaccination and the risk of cardiovascular and thromboembolic events after SARS-CoV-2 infection: a systematic review and meta-analysis"

**Supplementary material**

**Contents**

Figures	3

Figure 1 Study designs of cohort and case-control studies meeting review inclusion criteria	3

Figure 2 SARS-CoV-2 infection periods of included studies	4

Figure 3 Risks of composite CVD, CeVD, and VTE outcomes after SARS-CoV-2 infection in vaccinated individuals compared with unvaccinated individuals	5

Figure 4 Risks of CVD, CeVD, and VTE outcomes in booster vaccinated individuals compared with primary course vaccinated individuals	7

Tables	8

Table 1 PRISMA 2020 checklist^3^	8

Table 2 Study and baseline participant characteristics of included studies	11

Table 3 Vaccination characteristics of included studies	14

Table 4 Risk of bias judgement of included studies using ROBINS-E	17

Table 5 Vaccination impact on risk of CVD, CeVD, and VTE outcomes after SARS-CoV-2 infection (adjusted risks or populations weighted/matched for confounding variables)	19

Table 6 Vaccination impact on risk of CVD, CeVD, and VTE outcomes after SARS-CoV-2 infection by SARS-CoV-2 infection severity (adjusted risks or populations weighted/matched for confounding variables)	27

Table 7 Vaccination impact on risk of CVD, CeVD, and VTE outcomes after SARS-CoV-2 infection by SARS-CoV-2 variant (adjusted risks or populations weighted/matched for confounding variables)	28

Table 8 Vaccination impact on risk of CVD, CeVD, and VTE outcomes after SARS-CoV-2 infection by vaccine (adjusted risks or populations weighted/matched for confounding variables)	29

Table 9 Vaccination impact on risk of CVD, CeVD, and VTE outcomes after SARS-CoV-2 infection by prior CVD/CeVD/VTE disease status (adjusted risks or populations weighted/matched for confounding variables)	30

Table 10 Vaccination impact on risk of CVD, CeVD, and VTE outcomes after SARS-CoV-2 infection by sex (adjusted risks or populations weighted/matched for confounding variables)	30

Table 11 Vaccination impact on risk of CVD, CeVD, and VTE outcomes after SARS-CoV-2 infection by comorbidity status (adjusted risks or populations weighted/matched for confounding variables)	31

Table 12 Vaccination impact on risk of CVD, CeVD, and VTE outcomes after SARS-CoV-2 infection by immunocompromised status (adjusted risks or populations weighted/matched for confounding variables)	32

Table 13 Vaccination impact on risk of CVD, CeVD, and VTE outcomes after SARS-CoV-2 infection by age (adjusted risks or populations weighted/matched for confounding variables)	33

Table 14 Risk of CVD, CeVD, and VTE events after SARS-CoV-2 infection, stratified by vaccination status (adjusted risks or populations weighted/matched for confounding variables)	36

Table 15 Outcomes included in meta-analyses	40

Table 16 Meta-analysis leave-one-out sensitivity analyses	41

Table 17 Meta-analysis sensitivity analyses	42

Table 18 GRADE certainty of evidence assessment	43

Search strategies	44

EMBASE strategy	44

Medline strategy	46

PubMed strategy	49

Meta-analysis feasibility assessment	53

References	55

### Figures

#### Figure 1 Study designs of cohort and case-control studies meeting review inclusion criteria

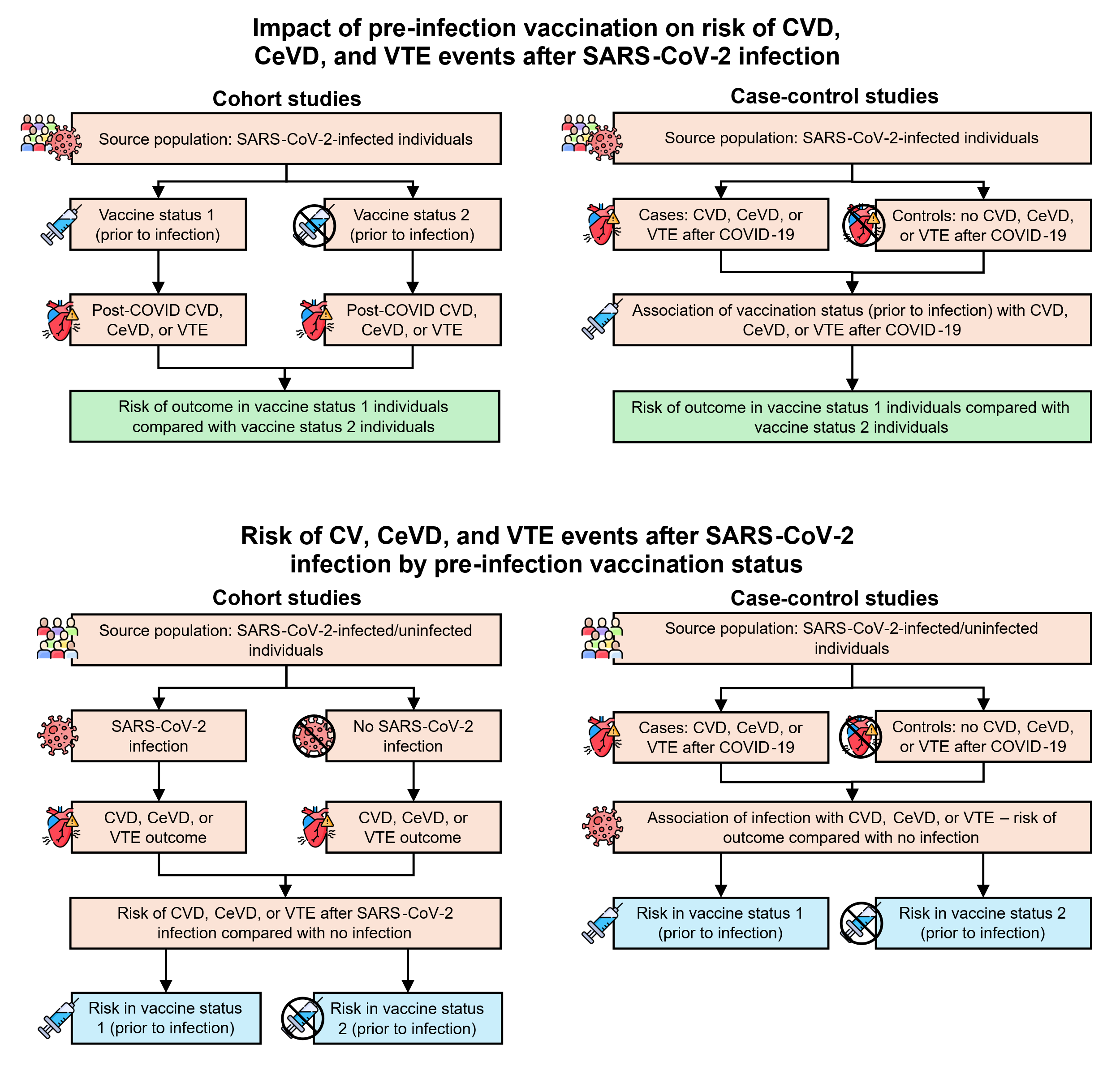

Figure depicts the different designs (retrospective cohort or retrospective case-control studies) of the studies included by this review. Studies reporting impact of vaccination on risk of outcomes after infection only included individuals with a record of SARS-CoV-2 infection. Studies reporting risk of outcomes after infection by vaccination status included both infected and uninfected individuals. Icons are illustrative only and do not indicate only vaccinated or unvaccinated; vaccine status groups include vaccinated, unvaccinated, specific numbers of doses, or monovalent/bivalent mRNA vaccines.

CeVD, cerebrovascular disease; CVD, cardiovascular disease; VTE, venous thromboembolism

#### Figure 2 SARS-CoV-2 infection periods of included studies

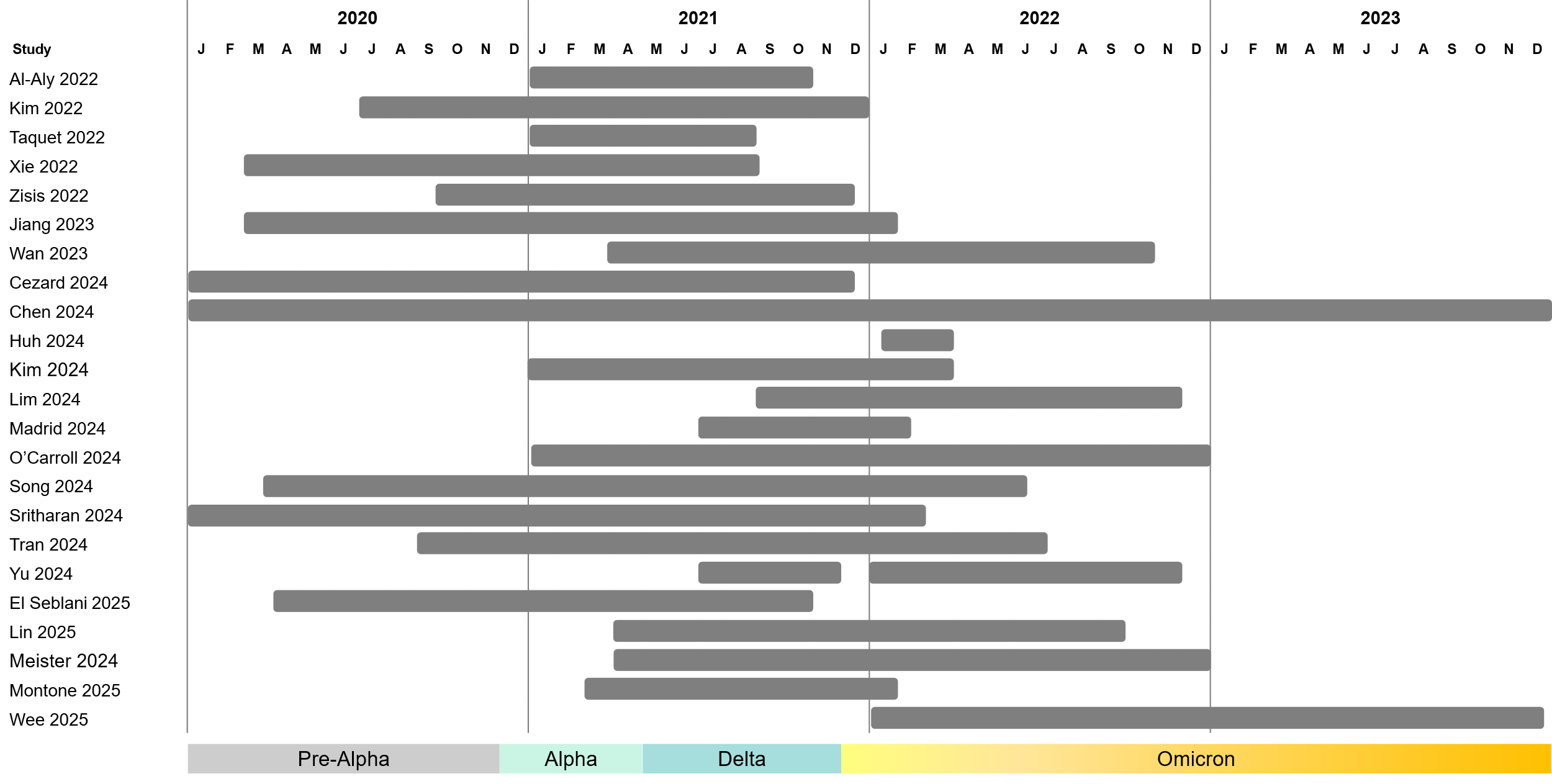

Figure shows the reported SARS-CoV-2 infection periods of all included studies. Estimated main variants are also presented; variant start dates are taken from dates they were declared a variant of concern.^1,2^

#### Figure 3 Risks of composite CVD, CeVD, and VTE outcomes after SARS-CoV-2 infection in vaccinated individuals compared with unvaccinated individuals

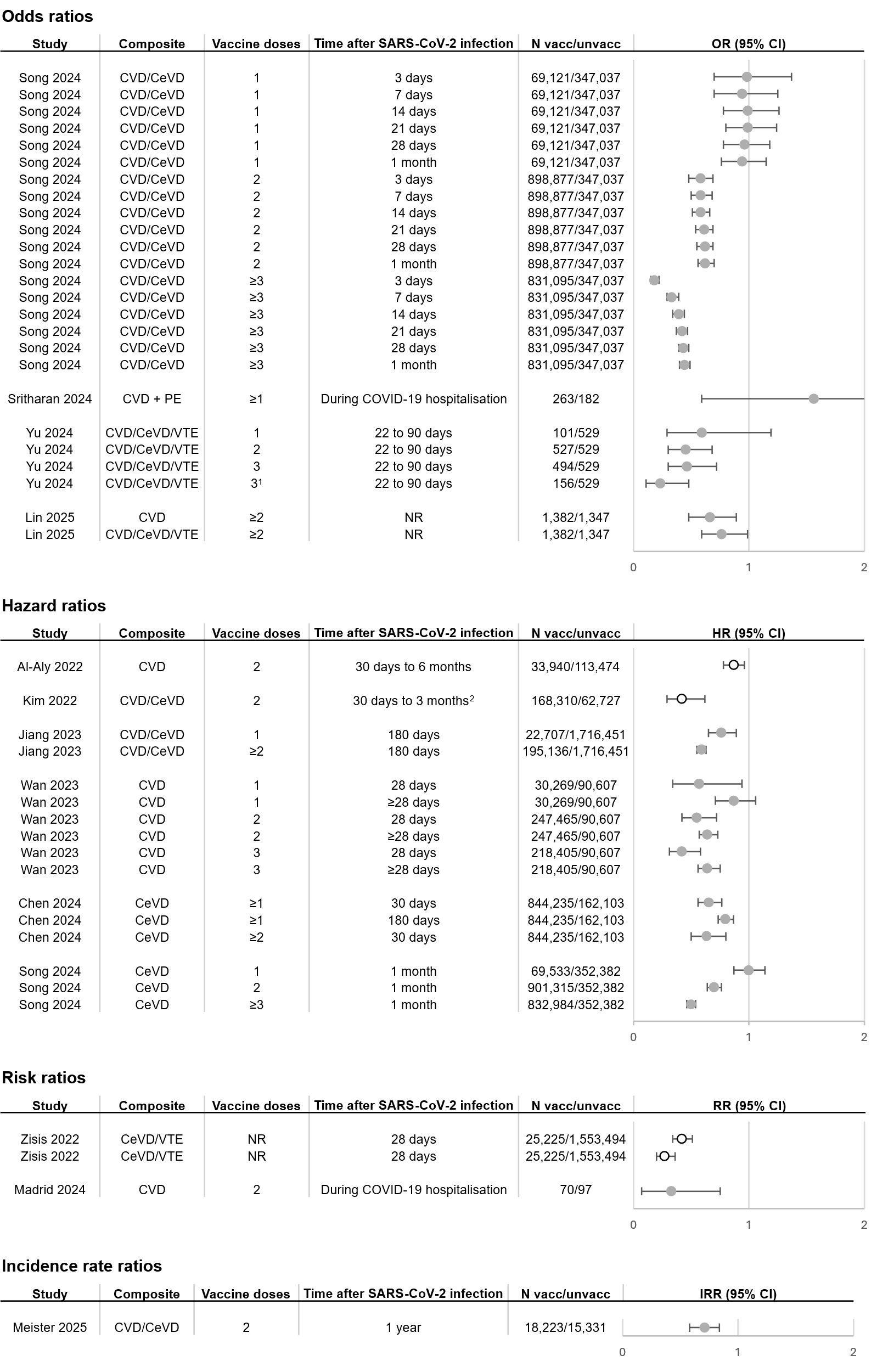

● Omicron and pre-Omicron variants; ○ Pre-Omicron variants only.

Risks of composite cardiovascular and/or cerebrovascular events after SARS-CoV-2 infection in individuals vaccinated before infection compared with unvaccinated individuals. Figure includes all studies identified by the systematic review that reported composite outcomes. Vaccine doses refer to the number of doses received by vaccinated individuals.

Composite outcome definitions:

- Song 2024: Coronary heart disease, stroke.
- Sritharan 2024: New onset atrial fibrillation or flutter, high grade atrioventricular block, new cardiomyopathy or heart failure, pericarditis, myocarditis or myopericarditis.
- Yu 2024: Stroke, myocardial infarction, heart failure, atrial fibrillation, coronary artery disease, myocarditis, pericarditis, deep vein thrombosis, cardiomyopathy, cardiovascular mortality.
- Lin 2025, CV: Acute myocardial infarction, other ischaemic heart disease, atrial fibrillation, ventricular fibrillation, other arrhythmias, cardiomyopathy, congestive heart failure.
- Lin 2025, CV/CBV/VTE: Acute myocardial infarction, other ischaemic heart disease, atrial fibrillation, ventricular fibrillation, other arrhythmias, cardiomyopathy, congestive heart failure, pulmonary embolism, deep vein thrombosis, superficial vein thrombosis, other thrombosis, cerebrovascular stroke.Al-Aly 2022: Acute coronary disease, atrial fibrillation, heart failure, hypertension, myocardial infarction, myocarditis, other dysrhythmias, pericarditis, tachycardia.
- Kim 2022: Acute myocardial infarction, ischaemic stroke.
- Jiang 2023: Major adverse cardiovascular events
- Wan 2023: Heart disease, stroke, heart failure.
- Chen 2024: Cerebral infarction, subarachnoid haemorrhage, intracerebral haemorrhage, other non-traumatic intracranial haemorrhage, occlusion and stenosis of precerebral arteries, occlusion and stenosis of cerebral arteries, other cerebrovascular diseases.

CeVD, cerebrovascular; CV, cardiovascular; CIs, confidence intervals; HR, hazard ratio; IRR, incidence rate ratio; OR, odds ratio; RR, risk ratio or relative risk; VTE, venous thromboembolism.

#### Figure 4 Risks of CVD, CeVD, and VTE outcomes in booster vaccinated individuals compared with primary course vaccinated individuals

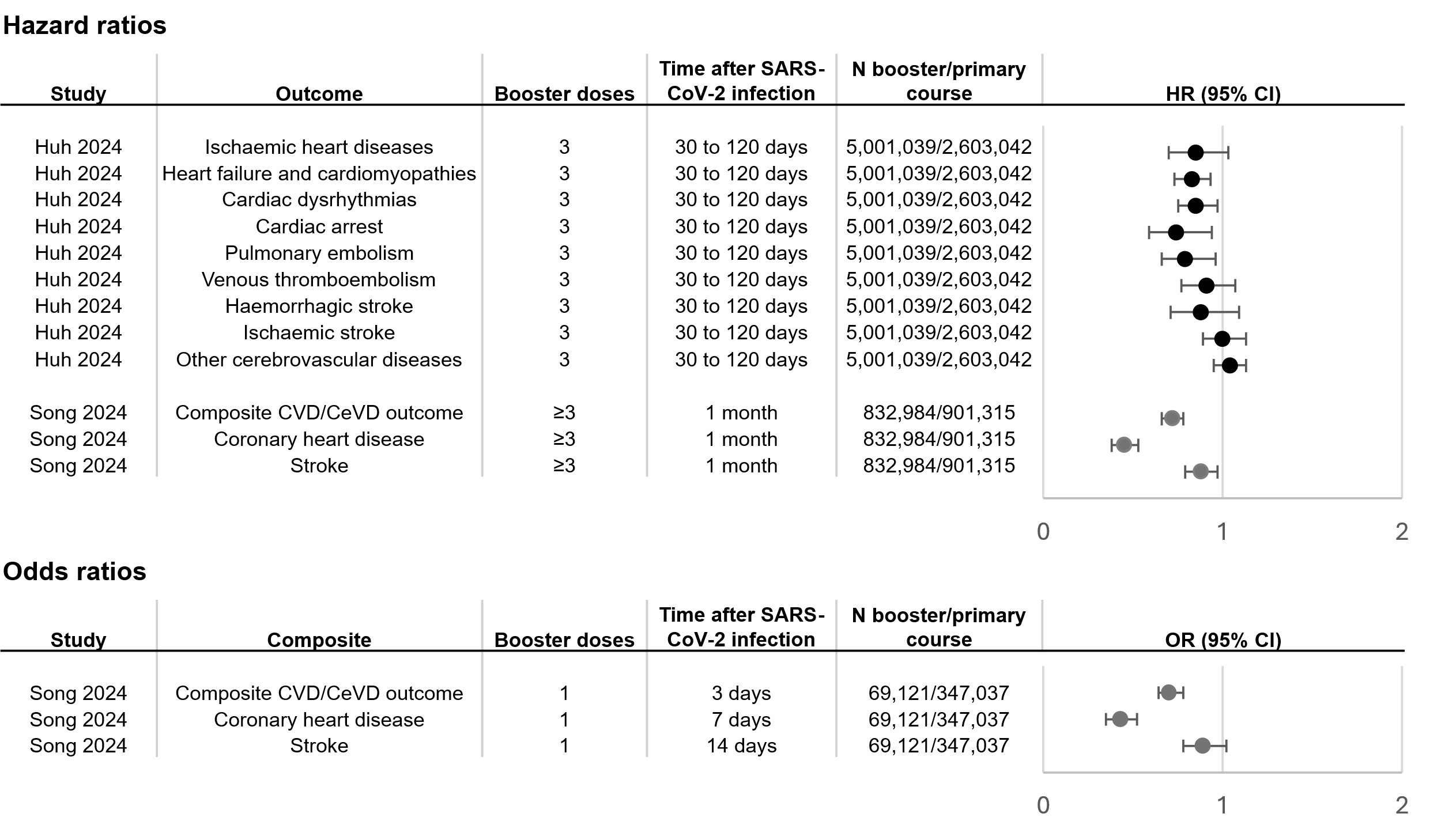

● Omicron variant only; ● Omicron and pre-Omicron variants.

Risks of cardiovascular, cerebrovascular, or venous thromboembolism events after SARS-CoV-2 infection in individuals with booster doe vaccination before infection compared with individuals with only primary course (two vaccine doses) before infection . Figure includes all studies identified by the systematic review that reported impact of booster dose vaccination compared with primary course vaccination. Booster doses refer to the number of vaccine doses received by booster dose-vaccinated individuals.

Composite CVD/CeVD outcome reported by Song 2024 includeds coronary heart disease and stroke.

### Tables

#### Table 1 PRISMA 2020 checklist^3^

| **Section and Topic** | **Item #** | **Checklist item** | **Location where item is reported** |
| --- | --- | --- | --- |
| **TITLE** | | |  |
| Title | 1 | Identify the report as a systematic review. | Title |
| **ABSTRACT** | | |  |
| Abstract | 2 | See the PRISMA 2020 for Abstracts checklist. | Abstract |
| **INTRODUCTION** | | |  |
| Rationale | 3 | Describe the rationale for the review in the context of existing knowledge. | Introduction |
| Objectives | 4 | Provide an explicit statement of the objective(s) or question(s) the review addresses. | Introduction |
| **METHODS** | | |  |
| Eligibility criteria | 5 | Specify the inclusion and exclusion criteria for the review and how studies were grouped for the syntheses. | Methods |
| Information sources | 6 | Specify all databases, registers, websites, organisations, reference lists and other sources searched or consulted to identify studies. Specify the date when each source was last searched or consulted. | Methods: search strategy |
| Search strategy | 7 | Present the full search strategies for all databases, registers and websites, including any filters and limits used. | Supplementary: search strategies |
| Selection process | 8 | Specify the methods used to decide whether a study met the inclusion criteria of the review, including how many reviewers screened each record and each report retrieved, whether they worked independently, and if applicable, details of automation tools used in the process. | Methods: study selection and data extraction |
| Data collection process | 9 | Specify the methods used to collect data from reports, including how many reviewers collected data from each report, whether they worked independently, any processes for obtaining or confirming data from study investigators, and if applicable, details of automation tools used in the process. | Methods: study selection and data extraction |
| Data items | 10a | List and define all outcomes for which data were sought. Specify whether all results that were compatible with each outcome domain in each study were sought (e.g. for all measures, time points, analyses), and if not, the methods used to decide which results to collect. | Methods: study selection and data extraction |
|  | 10b | List and define all other variables for which data were sought (e.g. participant and intervention characteristics, funding sources). Describe any assumptions made about any missing or unclear information. | Methods: study selection and data extraction |
| Study risk of bias assessment | 11 | Specify the methods used to assess risk of bias in the included studies, including details of the tool(s) used, how many reviewers assessed each study and whether they worked independently, and if applicable, details of automation tools used in the process. | Methods: quality assessment |
| Effect measures | 12 | Specify for each outcome the effect measure(s) (e.g. risk ratio, mean difference) used in the synthesis or presentation of results. | Methods: study selection and data extraction |
| Synthesis methods | 13a | Describe the processes used to decide which studies were eligible for each synthesis (e.g. tabulating the study intervention characteristics and comparing against the planned groups for each synthesis (item #5)). | Methods: statistical analysis |
|  | 13b | Describe any methods required to prepare the data for presentation or synthesis, such as handling of missing summary statistics, or data conversions. | Not applicable |
|  | 13c | Describe any methods used to tabulate or visually display results of individual studies and syntheses. | Not applicable |
|  | 13d | Describe any methods used to synthesize results and provide a rationale for the choice(s). If meta-analysis was performed, describe the model(s), method(s) to identify the presence and extent of statistical heterogeneity, and software package(s) used. | Methods: statistical analysis |
|  | 13e | Describe any methods used to explore possible causes of heterogeneity among study results (e.g. subgroup analysis, meta-regression). | Methods: statistical analysis |
|  | 13f | Describe any sensitivity analyses conducted to assess robustness of the synthesized results. | Methods: statistical analysis |
| Reporting bias assessment | 14 | Describe any methods used to assess risk of bias due to missing results in a synthesis (arising from reporting biases). | Methods: quality assessment |
| Certainty assessment | 15 | Describe any methods used to assess certainty (or confidence) in the body of evidence for an outcome. | Methods: quality assessment |
| **RESULTS** | | |  |
| Study selection | 16a | Describe the results of the search and selection process, from the number of records identified in the search to the number of studies included in the review, ideally using a flow diagram. | Results: systematic review of the literature |
|  | 16b | Cite studies that might appear to meet the inclusion criteria, but which were excluded, and explain why they were excluded. | Not applicable |
| Study characteristics | 17 | Cite each included study and present its characteristics. | Results: characteristics of included studies |
| Risk of bias in studies | 18 | Present assessments of risk of bias for each included study. | Results: characteristics of included studies |
| Results of individual studies | 19 | For all outcomes, present, for each study: (a) summary statistics for each group (where appropriate) and (b) an effect estimate and its precision (e.g. confidence/credible interval), ideally using structured tables or plots. | Results: impact of COVID-19 vaccination on risk of CVD/CeVD/VTE events post-SARS-CoV-2 infection; Results: risk of CVD/CeVD events post-SARS-CoV-2 infection compared with no infection, stratified by vaccine status; Supplementary: tables 5-14 |
| Results of syntheses | 20a | For each synthesis, briefly summarise the characteristics and risk of bias among contributing studies. | Meta-analyses: impact of COVID-19 vaccination on risk of post-SARS-CoV-2 infection CVD/CeVD/VTE events; Results: figures 2-3; Supplementary: table 15 |
|  | 20b | Present results of all statistical syntheses conducted. If meta-analysis was done, present for each the summary estimate and its precision (e.g. confidence/credible interval) and measures of statistical heterogeneity. If comparing groups, describe the direction of the effect. | Results: figures 2-3; |
|  | 20c | Present results of all investigations of possible causes of heterogeneity among study results. | Supplementary: table 17 |
|  | 20d | Present results of all sensitivity analyses conducted to assess the robustness of the synthesized results. | Supplementary: tables 16-17 |
| Reporting biases | 21 | Present assessments of risk of bias due to missing results (arising from reporting biases) for each synthesis assessed. | Not applicable |
| Certainty of evidence | 22 | Present assessments of certainty (or confidence) in the body of evidence for each outcome assessed. | Results: Meta-analyses: impact of COVID-19 vaccination on risk of post-SARS-CoV-2 infection CVD/CeVD/VTE events |
| **DISCUSSION** | | |  |
| Discussion | 23a | Provide a general interpretation of the results in the context of other evidence. | Discussion |
|  | 23b | Discuss any limitations of the evidence included in the review. | Discussion: review limitations |
|  | 23c | Discuss any limitations of the review processes used. | Discussion: review limitations |
|  | 23d | Discuss implications of the results for practice, policy, and future research. | Discussion: clinical implications |
| **OTHER INFORMATION** | | |  |
| Registration and protocol | 24a | Provide registration information for the review, including register name and registration number, or state that the review was not registered. | Methods |
|  | 24b | Indicate where the review protocol can be accessed, or state that a protocol was not prepared. | Methods |
|  | 24c | Describe and explain any amendments to information provided at registration or in the protocol. | Not applicable |
| Support | 25 | Describe sources of financial or non-financial support for the review, and the role of the funders or sponsors in the review. | Funding |
| Competing interests | 26 | Declare any competing interests of review authors. | Declarations |
| Availability of data, code and other materials | 27 | Report which of the following are publicly available and where they can be found: template data collection forms; data extracted from included studies; data used for all analyses; analytic code; any other materials used in the review. | Data availability |

#### Table 2 Study and baseline participant characteristics of included studies

| **Study** | **Excludes those with outcomes prior to study inclusion** | **COVID-19 variants(s) reported*** | **Subgroup** | **Unweighted** | | | | | **Weighting/ matching** | **Weighted / matched** | | | | |
| --- | --- | --- | --- | --- | --- | --- | --- | --- | --- | --- | --- | --- | --- | --- |
|  |  |  |  | **Age, mean (SD)** | **Age, median (IQR)** | **Male %** | **COVID-19 severity** | **Prior influenza vaccination** |  | **Age, mean (SD)** | **Age, median (IQR)** | **Male %** | **COVID-19 severity** | **Prior influenza vaccination** |
| Al-Aly 2022^4^ | No | NR (pre-Omicron) | Vaccinated, COVID-19 | 66.63 (13.84) | - | 91.0% | NR | In prior 2 years: 28.28% | Overlap weighting | 62.82 (14.87) | - | 88.85% | NR | NR |
|  |  |  | Unvaccinated, COVID-19 | 57.81 (15.94) | - | 86.92% | NR | In prior 2 years: 23.33% |  | 62.77 (14.71) | - | 88.65% | NR | NR |
| Kim 2022^5^ | Yes (past 30 days) | NR (pre-Omicron) | Vaccinated | - | 57 (42–66) | 47.04% | Severe: 3.15% Critical: 1.05% | NR | IPTW | - | 54 (38–65) | 47.21% | Severe: 2.84%  Critical: 0.95% | NR |
|  |  |  | Unvaccinated | - | 42 (31–58) | 48.48% | Severe: 9.78% Critical: 5.60% | NR |  | - | 52 (37–68) | 45.11% | Severe: 12.45%  Critical: 8.52% | NR |
| Taquet 2022^6^ | No | Pre-Omicron | Vaccinated | 57.0 (17.9) | - | 40.6% | NR | NR | PS matching | 56.5 (18.0) | - | 40.1% | NR | NR |
|  |  |  | Unvaccinated | 51.9 (23.1) | - | 41.7% | NR | At any time: 100% |  | 57.6 (20.6) | - | 39.2% | NR | At any time: 100% |
| Xie 2022^7^ | Yes (past 1 year) | Pre-Omicron | COVID-19 | 64.32 (8.03) | - | 43.8% | Non-hospitalised: 100% | NR | PS matching | 64.33 (8.03) | - | 43.8% | NR | NR |
|  |  |  | Uninfected | 67.96 (8.03) | - | 41.5% | Non-hospitalised: 100% | NR |  | 64.31 (7.92) | - | 44.0% | NR | NR |
| Zisis 2022^8^ | No | Pre-Omicron | Vaccinated | 54.82 (17.77) | - | 40.16% | NR | NR | PS matching | 54.82 (7.77) | - | 40.16% | NR | NR |
|  |  |  | Unvaccinated | 42.91 (21.84) | - | 43.95% | NR | NR |  | 55.06 (17.96) | - | 40.02% | NR | NR |
| Jiang 2023^9^ | No | NR (pre-Omicron, Omicron) | All | 45.2 | - | 44.1% | NR | NR | None | - | - | - | - |  |
| Wan 2023^10^ | Yes | Omicron, pre-Omicron | Unvaccinated | 56.4 (21.6) | - | 41.6% | Severe: 0.6% | NR | IPTW | 49.0 (20.4) | - | 43.8% | Severe: 0.2% | NR |
|  |  |  | 1 dose vaccinated | 56.8 (19.5) | - | 42.3% | Severe: 0.4% | NR |  | 49.7 (19.3) | - | 44.1% | Severe: 0.2% | NR |
|  |  |  | 2 dose vaccinated | 48.8 (17.1) | - | 43.1% | Severe: 0.1% | NR |  | 50.8 (17.5) | - | 43.4% | Severe: 0.2% | NR |
|  |  |  | 3 dose vaccinated | 50.3 (15.8) | - | 44.5% | Severe: 0.1% | NR |  | 50.8 (16.2) | - | 43.6% | Severe: 0.2% | NR |
| Cezard 2024^11^ | Yes | Delta | Vaccinated (COVID-19 and uninfected) | NR | NR | 47.9% | NR | NR | None | - | - | - | - | - |
|  |  |  | Unvaccinated (COVID-19 and uninfected) | NR | NR | 58.00% | NR | NR |  | - | - | - | - | - |
| Chen 2024^12^ | Yes | Pre-Omicron, Omicron | Vaccinated | 51.0 (17.5) | - | 37.2% | NR | Record of quadrivalent (IIV4) vaccination: 6.0% Record of vaccination: 5.2% | PS matching | 51.0 (17.5) | - | 37.2% | NR | Record of quadrivalent (IIV4) vaccination: 6.0% Record of vaccination: 5.2% |
|  |  |  | Unvaccinated | 46.2 (17.6) | - | 41.1% | NR | Record of quadrivalent (IIV4) vaccination: 2.2% Record of vaccination: 1.5% |  | 50.6 (17.3) | - | 36.2% | NR | Record of quadrivalent (IIV4) vaccination: 6.0% Record of vaccination: 4.7% |
| Huh 2024^13^ | Yes (past 1 year) | Omicron BA.1 and BA.2 | Vaccinated (2 or 3 doses) | 43.90 (16.31) | - | 43.71% | Mild: 99.44% Severe: 0.52% Critical: 0.04% | In 2020 and 2021: 39.86% | IPTW | 43.90 (16.77) | - | 43.61% | Mild: 99.28% Severe: 0.66% Critical: 0.06% | In 2020 and 2021: 39.28% |
|  |  |  | Unvaccinated | 43.66 (17.43) | - | 41.54% | Mild: 96.70% Severe: 2.97% Critical: 0.33% | In 2020 and 2021: 27.63% |  | 43.74 (77.99) | - | 42.67% | Mild: 99.18%  Severe: 0.76%  Critical:0.06% | In 2020 and 2021: 38.90% |
|  |  |  | 2 doses | 36.67 (12.69) | - | 45.94% | Mild: 99.58% Severe: 0.39% Critical: 0.03% | In 2020 and 2021: 28.82% |  | 43.53 (27.99) | - | 41.72% | Mild: 99.27%  Severe: 0.69%  Critical:0.04% | In 2020 and 2021: 39.50% |
|  |  |  | 3 doses | 47.66 (16.72) | - | 42.56% | Mild: 99.36% Severe: 0.59% Critical: 0.04% | In 2020 and 2021: 45.61% |  | 43.65 (20.71) | - | 43.36% | Mild: 99.43%  Severe: 0.54%  Critical:0.04% | In 2020 and 2021: 39.80% |
| Kim 2024^14^ | Yes | NR (pre-Omicron Omicron) | All | 47.6 (13.9) | - | 60.8% | Mild-low risk: 57.7%  Mild-high risk: 40.9%  Serious: 1.3%  Severely serious: 0.1% | NR | 1:10 exact matching (infected / uninfected) | - | - | - | - | - |
| Lim 2024^15^ | No | Delta, Omicron | COVID-19 | 51 (17.25) | - | 55.8% | NR | NR | IPTW | 48 (17.0) | - | 48.7% | NR | NR |
|  |  |  | Uninfected | 48 (17.7) | - | 48.1% | NR | NR |  | 48 (17.7) | - | 48.6% | NR | NR |
| Madrid 2024^16^ | No | Delta, Omicron | Vaccinated | 67.21 (95% CI 63.97; 70.59) | - | 70% | Hospitalised: 100% | NR | None | - | - | - | - | - |
|  |  |  | Unvaccinated | 51.10 (95% CI 48.12; 54.04) | - | 68% | Hospitalised: 100% | NR |  | - | - | - | - | - |
| O’Carroll 2024^17^ | No | Pre-Delta, Delta, Omicron | All | - | 36.0 (28.0–44.8) | 63.3% | Outpatient only: 91.3% | NR | None | - | - | - | - | - |
| Song 2024^18^ | Yes | Original, Alpha, Beta, Delta, Omicron | COVID+ (overall) | 44.39 (16.19) | - | 46.8% | NR | NR | None | - | - | - | - | - |
|  |  |  | COVID+, unvaccinated | - | - | - | - | - | 1:3 exact matching (infected/ uninfected) | 44.01 (16.17) | - | 48.6% | NR | NR |
|  |  |  | COVID+, 1 dose | - | - | - | - | - |  | 44.15 (14.88) | - | 56.6% | NR | NR |
|  |  |  | COVID+, 2 doses | - | - | - | - | - |  | 40.27 (14.76) | - | 46.6% | NR | NR |
|  |  |  | COVID+, ≥3 doses | - | - | - | - | - |  | 49.02 (16.53) | - | 45.3% | NR | NR |
|  |  |  | Non-infected matched group | - | - | - | - | - |  | 44.33 (16.10) | - | 46.7% | NR | NR |
| Sritharan 2024^19^ | No | Alpha, Delta, Omicron | Vaccinated | 66.5 (18.4) | - | 56.0% | Hospitalised: 100% | NR | None | - | - | - | - | - |
|  |  |  | Unvaccinated | 53.3 (19.2) | - | 54.7% | Hospitalised: 100% | NR |  | - | - | - | - | - |
| Tran 2024^20^ | No | NR (pre-Omicron, Omicron) | Cases (thromboembolic events outcome, COVID-19 and uninfected) | NR | NR | 51.66% | NR | NR | None | - | - | - | - | - |
|  |  |  | Controls (physical injury, COVID-19 and uninfected) | NR | NR | 42.37% | NR | NR |  | - | - | - | - | - |
| Yu 2024^21^ | Yes | NR (pre-Omicron, Omicron) | Cases (≥1 cardiac-related outcome) | - | - | - | - | - | 1:10 random matching | 70.4 (13.4) | - | 60% | NR | NR |
|  |  |  | Controls (no cardiac-related outcome) | - | - | - | - | - |  | 68.7 (12.0) | - | 61% | NR | NR |
| El Seblani 2025^22^ | No | Delta, Omicron | Vaccinated | 71 (12) | - | 51% | NR | NR | PS matching | 71 (12) | - | 52% | NR | NR |
|  |  |  | Unvaccinated | 65 (15) | - | 55% | NR | NR |  | 71 (12) | - | 51% | NR | NR |
| Lin 2025^23^ | No | Alpha, Delta, Omicron | Vaccinated | - | 67 (58–76) | 46% | NR | NR | IPTW | 65.7 (1.37) | - | 46.2% | NR | NR |
|  |  |  | Unvaccinated | - | 62 (52–72) | 42% | NR | NR |  | 64.6 (1.40) | - | 47.1% | NR | NR |
| Meister 2025^24^ | Yes (past 5 years) | Alpha, Delta, Omicron | Vaccinated | 55 (11.3) | - | 42.2% | NR | NR | IPTW | NR | - | NR | NR | NR |
|  |  |  | Unvaccinated | 54 (11.0) | - | 57.8% | NR | NR |  | NR | - | NR | NR | NR |
| Montone 2025^25^ | No | NR (pre-Omicron, Omicron) | Overall population | 67.7 (14.8) | - | 60.8% | Hospitalised: 100% | NR | None | - | - | - | - | - |
|  |  |  | Patients with myocardial injury | 77.4 (13.1) | - | 64.1% | Hospitalised: 100% | NR |  | - | - | - | - | - |
|  |  |  | Patients without myocardial injury | 66.1 (14.5) | - | 60.3% | Hospitalised: 100% | NR |  | - | - | - | - | - |
| Wee 2025^26^ | Yes | Omicron | Ancestral booster | NR | NR | 46.78% | NR | NR | Overlap weighting | NR | NR | 46.24% | NR | NR |
|  |  |  | Bivalent booster | NR | NR | 45.26% | NR | NR |  | NR | NR | 46.24% | NR | NR |

* Where variant not reported (NR), reviewers have estimated whether circulating variants are pre-Omicron or Omicron-onwards, based on COVID-19 infection periods (estimate in parentheses).

IPTW, inverse probability of treatment weighting; IQR, interquartile range; NR, not reported; PS, propensity score; SD, standard deviation.

#### Table 3 Vaccination characteristics of included studies

| **Study** | **Vaccines** | **% vaccinated individuals receiving different vaccines (EMA-authorised vaccines only)** | **Minimum time after last dose to be considered vaccinated** | **Time from last vaccination to COVID-19 or post-COVID-19 outcome** |
| --- | --- | --- | --- | --- |
| Al-Aly 2022^4^ | BNT162b2  mRNA-1273  Ad26.COV2.S | BNT162b2: 47.9%  mRNA-1273: 40.4%  Ad26.COV2.S: 11.6% | 14 days | NR |
| Kim 2022^5^ | BNT162b2  mRNA-1273  ChAdOx1  Ad26.COV2.S | NR | 7 days | NR |
| Taquet 2022^6^ | BNT162b2  mRNA-1273  Ad26.COV2.S | BNT162b2: 65.1%  mRNA-1273: 9.0%  Ad26.COV2.S: 1.6%  Unknown: 24.4% | 14 days | NR |
| Xie 2022^7^ | NR | NR | NR | NR |
| Zisis 2022^8^ | NR | NR | NR | NR |
| Jiang 2023^9^ | BNT162b2  mRNA-1273  Ad26.COV2.S | NR | 14 days | NR |
| Wan 2023^10^ | BNT162b2  CoronaVac* | BNT162b2: 100% (excluding CoronaVac vaccinations) | NR | NR |
| Cezard 2024^11^ | NR | NR | NR | NR |
| Chen 2024^12^ | NR | NR | 7 days | Max time to COVID-19: 1 year |
| Huh 2024^13^ | BNT162b2  mRNA-1273  ChAdOx1 | NR | NR | NR |
| Kim 2024^14^ | BNT162b2  mRNA-1273  ChAdOx1  Ad26.COV2.S | NR | NR | NR |
| Lim 2024^15^ | BNT162b2  mRNA-1273 | NR | NR | NR |
| Madrid 2024^16^ | BNT162b2  mRNA-1273  ChAdOx1  Ad26.COV2.S | NR | 14 days | NR |
| O’Carroll 2024^17^ | BNT162b2  mRNA-1273  Ad26.COV2.S | NR | 14 days | NR |
| Song 2024^18^ | BNT162b2  mRNA-1273  ChAdOx1  Ad26.COV2.S | NR | 14 days | NR |
| Sritharan 2024^19^ | BNT162b2  mRNA-1273  ChAdOx1 | BNT162b2: 19.8%  mRNA-1273: 1.0%  ChAdOx1: 33.7%  Unknown: 48.2% | NR | NR |
| Tran 2024^20^ | BNT162b2  mRNA-1273 | NR | NR | NR |
| Yu 2024^21^ | BNT162b2  CoronaVac* | BNT162b2: 100% (excluding CoronaVac vaccinations) | 14 days | NR |
| El Seblani 2025^22^ | BNT162b2  mRNA-1273  ChAdOx1  Ad26.COV2.S | BNT162b2: 68.7%^5^  mRNA-1273: 27.9%^5^  ChAdOx1 or Ad26.COV2.S: 3.3%^5^ | 1 month | NR |
| Lin 2025^23^ | BNT162b2  mRNA-1273 | NR | NR | NR |
| Meister 2025^24^ | BNT162b2  mRNA-1273  ChAdOx1  Ad26.COV2.S | BNT162b2: ~90% | 14 days | NR |
| Montone 2025^25^ | BNT162b2  mRNA-1273  ChAdOx1  Ad26.COV2.S | BNT162b2: 18.1%  mRNA-1273: 1.1%  ChAdOx1: 2.6%  Ad26.COV2.S: 0.5%  Missing data: 12.3% | 14 days | Mean (SD) time to COVID-19: 122.9 (84.7) days |
| Wee 2025^26^ | BNT162b2  mRNA-1273 | NR | 7 days | Time to COVID-19, % bivalent-vaccinated individuals:  0-3 months: 27.11%  3-6 months: 35.71%  6-12 months: 37.18%  Time to COVID-19, % monovalent-vaccinated individuals:  0-3 months: 43.25%  3-6 months: 36.98%  6-12 months: 19.77% |

* Analysis including CoronaVac-vaccinated individuals not included in this review.

NR, not reported; SD, standard deviation.

#### Table 4 Risk of bias judgement of included studies using ROBINS-E

| Study | Overall risk of bias | Confounding | Measurement of the exposure | Selection of participants | Post-exposure interventions | Missing data | Measurement of outcome | Selection of reported result |
| --- | --- | --- | --- | --- | --- | --- | --- | --- |
| Al-Aly 2022 | Low | Low | Low | Low | Low | Low | Low | Low |
| Kim 2022 | Low | Low | Low | Low | Low | Low | Low | Low |
| Taquet 2022 | Some concerns | Some concerns^2^ | Low | Low | Low | Low | Low | Low |
| Xie 2022 | Low | Low | Low | Low | Low | Low | Low | Low |
| Zisis 2022 | High | High^3^ | Low | Low | Low | Low | Low | Low |
| Jiang 2023 | High | High^3^ | Low | Low | Low | Low | Low | Low |
| Wan 2023 | Some concerns | Some concerns^4^ | Low | Low | Low | Low | Low | Low |
| Cezard 2024 | Low | Low | Low | Low | Low | Low | Low | Low |
| Chen 2024 | Low | Low | Low | Low | Low | Low | Low | Low |
| Huh 2024 | Low | Low | Low | Low | Low | Low | Low | Low |
| Kim 2024 | Low | Low | Low | Low | Low | Low | Low | Low |
| Lim 2024 | Low | Low | Low | Low | Low | Low | Low | Low |
| Madrid 2024 | Very high^1^ | High^5^ | Low | Low | Low | High^7^ | High^9^ | Low |
| O’Carroll 2024 | High | High^3^ | Low | Low | Low | Low | Low | Low |
| Song 2024 | Low | Low | Low | Low | Low | Low | Low | Low |
| Sritharan 2024 | Very high^1^ | Some concerns^4^ | Low | Low | Low | High^8^ | High^10^ | Low |
| Tran 2024^20^ | Low | Low | Low | Low | Low | Low | Low | Low |
| Yu 2024^21^ | Low | Low | Low | Low | Low | Low | Low | Low |
| El Seblani 2025 | Some concerns | Some concerns^4^ | Low | Low | Low | Low | Low | Low |
| Lin 2025 | High | High^3^ | Low | Low | Low | Low | Low | Low |
| Meister 2025 | Low | Low | Low | Low | Low | Low | Low | Low |
| Montone 2025 | High | High^6^ | Low | Low | Low | Low | Low | Low |
| Wee 2025 | Low | Low | Low | Low | Low | Low | Low | Low |

Due to the study design required to address the review question, the start of follow-up in the studies was considered to be SARS-CoV-2 infection rather than from the intervention (vaccination).

^1^ Favours intervention, i.e. may overestimate events for unvaccinated and underestimate events for vaccinated. This is driven by all patients in study being hospitalised for SARS-CoV-2 infection, and the outcomes assessed during the hospitalisation stay.

^2^ Did not control for COVID-19 severity.
^3^ Did not control for COVID-19 severity or socioeconomic factors.
^4^ Did not control for socioeconomic factors.
^5^ Did not control for sex or socioeconomic factors.
^6^ Did not control for sex, COVID-19 severity, or socioeconomic factors.
^7^ Although the authors excluded the 19% of the cohort with missing or incomplete vaccination data from the analysis, this may introduce selection bias. Exclusion due to missing data was likely not random and may have been related to both vaccination status and true cardiac outcomes. No sensitivity analyses were performed to test whether excluding patients with missing vaccination status, biomarker data, or confounders affected results.
^8^ Vaccination status only known for 34% of patients. Cardiovascular complications were only recorded if clinically indicated investigations were performed (e.g. troponin measured in 986/1714 patients [57.5%]), and authors acknowledged that this may have led to under-ascertainment of subclinical or less obvious cardiovascular outcomes. Exclusion due to missing data was likely related to the true value of outcomes (e.g. patients without vaccination records or troponin testing may have had systematically different risks).
^9^ Clinicians may be aware of unvaccinated status; unvaccinated patients may have been investigated more aggressively if perceived as higher risk.
^10^ Clinicians may be aware of unvaccinated status, unvaccinated patients may have been investigated more aggressively if perceived as higher risk.

#### Table 5 Vaccination impact on risk of CVD, CeVD, and VTE outcomes after SARS-CoV-2 infection (adjusted risks or populations weighted/matched for confounding variables)

| **Study** | **Time from SARS-CoV-2 infection** | **Vaccination comparison** | **Outcome** | **Risk estimate (95% CI)** |
| --- | --- | --- | --- | --- |
| Al-Aly 2022 | 30 days to 6 months | Vaccinated vs unvaccinated (2 vs 0 doses) | Composite CVD (“Cardiovascular outcome”: Acute coronary disease, atrial fibrillation, heart failure, hypertension, myocardial infarction, myocarditis, other dysrhythmias, pericarditis, tachycardia) | HR 0.87 (0.78–0.96) |
|  |  |  | Acute coronary disease | HR 0.92 (0.75–1.13) |
|  |  |  | Atrial fibrillation | HR 0.79 (0.67–0.94) |
|  |  |  | Heart failure | HR 0.98 (0.80–1.19) |
|  |  |  | Hypertension | HR 0.92 (0.79–1.08) |
|  |  |  | Myocardial infarction | HR 0.70 (0.42–1.15) |
|  |  |  | Myocarditis | HR 0.05 (0.01–0.38) |
|  |  |  | Other dysrhythmias | HR 0.52 (0.36–0.75) |
|  |  |  | Pericarditis | HR 0.82 (0.31–2.19) |
|  |  |  | Tachycardia | HR 0.57 (0.44–0.74) |
|  |  |  | Acute haemorrhagic cerebrovascular disease | HR 1.02 (0.55–1.89) |
|  |  |  | Stroke (ischaemic) | HR 0.79 (0.60–1.04) |
|  |  |  | Deep vein thrombosis | HR 0.59 (0.44–0.78) |
|  |  |  | Pulmonary embolism | HR 0.31 (0.24–0.41) |
| Kim 2022 | Median 90 (unvaccinated) and 84 (vaccinated) days (excluding first 30 days) | Vaccinated vs unvaccinated (2 vs 0 doses) | Composite CVD/CeVD: (“Composite cardiovascular events”: Acute myocardial infarction, ischaemic stroke) | HR 0.42 (0.29–0.62) |
|  |  |  | Acute myocardial infarction | HR 0.48 (0.25–0.94) |
|  |  |  | Ischaemic stroke | HR 0.40 (0.26–0.63) |
| Taquet 2022 | 6 months | Vaccinated vs unvaccinated (≥1 vs 0 doses) | Arrhythmia | HR 0.99 (0.91–1.08) |
|  |  |  | Cardiac failure | HR 1.06 (0.95–1.20) |
|  |  |  | Cardiomyopathy | HR 1.18 (0.97–1.43) |
|  |  |  | Cerebral haemorrhage | HR 1.33 (0.81–2.16) |
|  |  |  | Coronary disease | HR 1.04 (0.86–1.27) |
|  |  |  | Hypercoagulopathy/deep vein thrombosis/pulmonary embolism | HR 0.86 (0.75–0.99) |
|  |  |  | Hypertension | HR 1.00 (0.95–1.06) |
|  |  |  | Ischaemic stroke | HR 1.01 (0.80–1.26) |
|  |  |  | Myocarditis | HR 1.15 (0.49–2.66) |
|  |  | Vaccinated vs unvaccinated (1 vs 0 doses) | Arrhythmia | HR 1.01 (0.88–1.17) |
|  |  |  | Cardiac failure | HR 1.03 (0.86–1.24) |
|  |  |  | Cardiomyopathy | HR 1.16 (0.85–1.57) |
|  |  |  | Cerebral haemorrhage | HR 2.06 (0.94–4.53) |
|  |  |  | Coronary disease | HR 1.13 (0.81–1.58) |
|  |  |  | Hypercoagulopathy/deep vein thrombosis/pulmonary embolism | HR 0.94 (0.75–1.18) |
|  |  |  | Hypertension | HR 1.00 (0.92–1.10) |
|  |  |  | Ischaemic stroke | HR 1.49 (1.04–2.14) |
|  |  |  | Myocarditis | HR 1.30 (0.41–4.09) |
|  |  | Vaccinated vs unvaccinated (2 vs 0 doses) | Arrhythmia | HR 0.95 (0.86–1.05) |
|  |  |  | Cardiac failure | HR 1.06 (0.92–1.22) |
|  |  |  | Cardiomyopathy | HR 1.19 (0.95–1.50) |
|  |  |  | Cerebral haemorrhage | HR 1.71 (0.88–3.33) |
|  |  |  | Coronary disease | HR 1.17 (0.92–1.48) |
|  |  |  | Hypercoagulopathy/deep vein thrombosis/pulmonary embolism | HR 0.98 (0.82–1.17) |
|  |  |  | Hypertension | HR 1.11 (1.04–1.19) |
|  |  |  | Ischaemic stroke | HR 0.96 (0.74–1.25) |
|  |  |  | Myocarditis | HR 0.94 (0.34–2.60) |
| Xie 2022 | 30 days | Unvaccinated/partially vaccinated (0-1 vs 2 doses) | Venous thromboembolism | HR 5.50 (3.00–10.08) |
| Zisis 2022 | 28 days | Vaccinated vs unvaccinated | Hypertension | RR 0.45 (0.38–0.54) |
|  |  |  | Heart disease | RR 0.49 (0.43–0.57) |
|  |  |  | Thrombosis (strokes and venous thromboembolism) | RR 0.49 (0.43–0.57) |
|  | 90 days | Vaccinated vs unvaccinated | Hypertension | RR 0.33 (0.26–0.42) |
|  |  |  | Heart disease | RR 0.35 (0.29–0.44) |
|  |  |  | Thrombosis (strokes and venous thromboembolism) | RR 0.27 (0.20–0.36) |
| Jiang 2023 | 180 days | Vaccinated vs unvaccinated (≥2 vs 0 doses) | Composite CVD/CeVD (“Major adverse cardiovascular events”) | HR 0.59 (0.55–0.63) |
|  |  | Partially vaccinated vs unvaccinated (1 vs 0 doses) | Composite CVD/CeVD (“Major adverse cardiovascular events”) | HR 0.76 (0.65–0.89) |
| Wan 2023 | 28 days | Single dose vs unvaccinated (1 vs 0 doses) | Composite CVD/CeVD (“Cardiovascular disease”: coronary heart disease, stroke, heart failure) | HR 0.57 (0.34–0.94) |
|  |  |  | Stroke | HR 0.46 (0.23–0.92) |
|  |  |  | Coronary artery disease | HR 0.72 (0.33–1.59) |
|  |  |  | Heart failure | HR 0.65 (0.15–2.84) |
|  |  |  | CVD mortality | HR 0.51 (0.24–1.11) |
|  |  | Primary course vs unvaccinated (2 vs 0 doses) | Composite CVD/CeVD (“Cardiovascular disease”: heart disease, stroke, heart failure) | HR 0.55 (0.42–0.72) |
|  |  |  | Stroke | HR 0.60 (0.43–0.84) |
|  |  |  | Coronary artery disease | HR 0.59 (0.34–1.02) |
|  |  |  | Heart failure | HR 0.27 (0.11–0.65) |
|  |  |  | CVD mortality | HR 0.37 (0.18–0.73) |
|  |  | Booster dose vs unvaccinated (3 vs 0 doses) | Composite CVD/CeVD (“Cardiovascular disease”: heart disease, stroke, heart failure) | HR 0.42 (0.31–0.58) |
|  |  |  | Stroke | HR 0.34 (0.23–0.50) |
|  |  |  | Coronary artery disease | HR 0.71 (0.41–1.23) |
|  |  |  | Heart failure | HR 0.20 (0.07–0.54) |
|  |  |  | CVD mortality | HR 0.23 (0.07–0.82) |
|  | ≥28 days | Single dose vs unvaccinated (1 vs 0 doses) | Composite CVD/CeVD (“Cardiovascular disease”: heart disease, stroke, heart failure) | HR 0.87 (0.71–1.06) |
|  |  |  | Stroke | HR 0.95 (0.71–1.29) |
|  |  |  | Coronary artery disease | HR 0.92 (0.66–1.29) |
|  |  |  | Heart failure | HR 0.58 (0.36–0.93) |
|  |  |  | CVD mortality | HR 0.36 (0.19–0.70) |
|  |  | Primary course vs unvaccinated (2 vs 0 doses) | Composite CVD/CeVD (“Cardiovascular disease”: heart disease, stroke, heart failure) | HR 0.64 (0.57–0.73) |
|  |  |  | Stroke | HR 0.65 (0.54–0.78) |
|  |  |  | Coronary artery disease | HR 0.81 (0.66–0.99) |
|  |  |  | Heart failure | HR 0.41 (0.31–0.55) |
|  |  |  | CVD mortality | HR 0.36 (0.23–0.57) |
|  |  | Booster dose vs unvaccinated (3 vs 0 doses) | Composite CVD/CeVD (“Cardiovascular disease”: heart disease, stroke, heart failure) | HR 0.64 (0.56–0.75) |
|  |  |  | Stroke | HR 0.58 (0.46–0.73) |
|  |  |  | Coronary artery disease | HR 0.90 (0.72–1.12) |
|  |  |  | Heart failure | HR 0.41 (0.27–0.61) |
|  |  |  | CVD mortality | HR 0.33 (0.21–0.50) |
| Chen 2024 | 30 days | Vaccinated vs unvaccinated (≥1 vs 0 doses) | Composite CeVD (“Cerebrovascular diseases”: subarachnoid haemorrhage, intracerebral haemorrhage, other nontraumatic intracranial haemorrhage, cerebral infarction, occlusion and stenosis of precerebral arteries, occlusion and stenosis of cerebral arteries, other cerebrovascular diseases, cerebrovascular disorders in diseases classified elsewhere, sequalae of cerebrovascular disease) | HR 0.655 (0.561–0.765) |
|  |  |  | Subarachnoid haemorrhage | HR 0.571 (0.237–1.377) |
|  |  |  | Intracerebral haemorrhage | HR 0.577 (0.289–1.152) |
|  |  |  | Other nontraumatic intracranial haemorrhage | HR 0.880 (0.444–1.741) |
|  |  |  | Cerebral infarction | HR 0.617 (0.483–0.788) |
|  |  |  | Occlusion and stenosis of precerebral arteries | HR 0.735 (0.553–0.977) |
|  |  |  | Occlusion and stenosis of cerebral arteries | HR 1.644 (0.481–5.618) |
|  |  |  | Other cerebrovascular diseases | HR 0.570 (0.423–0.770) |
|  |  |  | Sequalae of cerebrovascular disease | HR 0.390 (0.225–0.676) |
|  |  | Vaccinated vs unvaccinated (≥2 vs 0 doses) | Composite CeVD (“Cerebrovascular diseases”: subarachnoid haemorrhage, intracerebral haemorrhage, other nontraumatic intracranial haemorrhage, cerebral infarction, occlusion and stenosis of precerebral arteries, occlusion and stenosis of cerebral arteries, other cerebrovascular diseases, cerebrovascular disorders in diseases classified elsewhere, sequalae of cerebrovascular disease) | HR 0.632 (0.499–0.800) |
|  |  |  | Subarachnoid haemorrhage | HR 0.470 (0.141–1.560) |
|  |  |  | Intracerebral haemorrhage | HR 0.539 (0.158–1.841) |
|  |  |  | Other nontraumatic intracranial haemorrhage | HR 0.629 (0.224–1.767) |
|  |  |  | Cerebral infarction | HR 0.672 (0.466–0.969) |
|  |  |  | Occlusion and stenosis of precerebral arteries | HR 0.610 (0.394–0.945) |
|  |  |  | Other cerebrovascular diseases | HR 0.620 (0.399–0.962) |
|  |  |  | Sequalae of cerebrovascular disease | HR 0.335 (0.121–0.930) |
|  | 180 days | Vaccinated vs unvaccinated (≥1 vs 0 doses) | Composite CeVD (“Cerebrovascular diseases”: subarachnoid haemorrhage, intracerebral haemorrhage, other nontraumatic intracranial haemorrhage, cerebral infarction, occlusion and stenosis of precerebral arteries, occlusion and stenosis of cerebral arteries, other cerebrovascular diseases, cerebrovascular disorders in diseases classified elsewhere, sequalae of cerebrovascular disease) | HR 0.797 (0.734–0.865) |
|  |  |  | Subarachnoid haemorrhage | HR 0.925 (0.581–1.473) |
|  |  |  | Intracerebral haemorrhage | HR 0.669 (0.457–0.980) |
|  |  |  | Other nontraumatic intracranial haemorrhage | HR 0.730 (0.518–1.028) |
|  |  |  | Cerebral infarction | HR 0.700 (0.612–0.801) |
|  |  |  | Occlusion and stenosis of precerebral arteries | HR 0.806 (0.703–0.925) |
|  |  |  | Occlusion and stenosis of cerebral arteries | HR 1.213 (0.707–2.081) |
|  |  |  | Other cerebrovascular diseases | HR 0.860 (0.738–1.003) |
|  |  |  | Cerebrovascular disorders in diseases classified elsewhere | HR 0.674 (0.234–1.944) |
|  |  |  | Sequalae of cerebrovascular disease | HR 0.741 (0.569–0.966) |
| Huh 2024 | 30 to 120 days | Vaccinated vs unvaccinated (2-3 vs 0 doses) | Ischaemic heart diseases | HR 0.73 (0.57–0.94) |
|  |  |  | Heart failure and cardiomyopathies | HR 0.55 (0.48–0.63) |
|  |  |  | Cardiac dysrhythmias | HR 0.72 (0.61–0.85) |
|  |  |  | Cardiac arrest | HR 0.41 (0.33–0.51) |
|  |  |  | Pulmonary embolism | HR 0.66 (0.52–0.84) |
|  |  |  | Venous thromboembolism | HR 0.54 (0.44–0.66) |
|  |  |  | Haemorrhagic stroke | HR 0.79 (0.61–1.03) |
|  |  |  | Ischaemic stroke | HR 0.92 (0.78–1.09) |
|  |  |  | Other cerebrovascular diseases | HR 0.99 (0.86–1.14) |
|  |  | Booster dose vs primary course (3 vs 2 doses) | Ischaemic heart diseases | HR 0.85 (0.70–1.03) |
|  |  |  | Heart failure and cardiomyopathies | HR 0.83 (0.73–0.93) |
|  |  |  | Cardiac dysrhythmias | HR 0.85 (0.75–0.97) |
|  |  |  | Cardiac arrest | HR 0.74 (0.59–0.94) |
|  |  |  | Pulmonary embolism | HR 0.79 (0.66–0.96) |
|  |  |  | Venous thromboembolism | HR 0.91 (0.77–1.07) |
|  |  |  | Haemorrhagic stroke | HR 0.88 (0.71–1.09) |
|  |  |  | Ischaemic stroke | HR 1.00 (0.89–1.13) |
|  |  |  | Other cerebrovascular diseases | HR 1.04 (0.95–1.13) |
| Kim 2024 | NR | Vaccinated vs unvaccinated (≥2 vs 0-1 doses) | Pulmonary embolism | HR 0.14 (0.09–0.22) |
|  |  |  | Deep vein thrombosis | HR 0.32 (0.19–0.54) |
| Madrid 2024 | NR (during acute COVID-19 hospitalisation) | Vaccinated vs unvaccinated (2 vs 0 doses) | Composite CVD (“Acute cardiac events”: myocardial infarction, heart failure, arrhythmia, myocarditis) | RR 0.33 (0.07–0.75) |
|  |  |  | Pericardial effusion | RR 1.27 (0.00–60.00) |
|  |  |  | Pulmonary embolism | RR 1.10 (0.42–2.33) |
| O’Carroll 2024 | 90 days | Primary course vs unvaccinated (2 vs 0 doses) | Venous thromboembolism | OR 0.28 (0.13–0.62) |
|  |  | Booster dose vs unvaccinated (≥3 vs 0 doses) | Venous thromboembolism | OR 0.06 (0.01–0.46) |
| Song 2024 | ≤3 days | Single dose vs unvaccinated (1 vs 0 doses) | Composite CVD/CeVD (“Cardiovascular disease”: coronary heart disease, stroke) | OR 0.98 (0.70–1.37) |
|  |  | Primary course vs unvaccinated (2 vs 0 doses) | Composite CVD/CeVD (“Cardiovascular disease”: coronary heart disease, stroke) | OR 0.58 (0.48–0.69) |
|  |  | Booster dose vs unvaccinated (≥3 vs 0 doses) | Composite CVD/CeVD (“Cardiovascular disease”: coronary heart disease, stroke) | OR 0.18 (0.15–0.22) |
|  | ≤7 days | Single dose vs unvaccinated (1 vs 0 doses) | Composite CVD/CeVD (“Cardiovascular disease”: coronary heart disease, stroke) | OR 0.94 (0.70–1.25) |
|  |  | Primary course vs unvaccinated (2 vs 0 doses) | Composite CVD/CeVD (“Cardiovascular disease”: coronary heart disease, stroke) | OR 0.58 (0.50–0.68) |
|  |  | Booster dose vs unvaccinated (≥3 vs 0 doses) | Composite CVD/CeVD (“Cardiovascular disease”: coronary heart disease, stroke) | OR 0.33 (0.29–0.39) |
|  | ≤14 days | Single dose vs unvaccinated (1 vs 0 doses) | Composite CVD/CeVD (“Cardiovascular disease”: coronary heart disease, stroke) | OR 0.99 (0.78–1.26) |
|  |  | Primary course vs unvaccinated (2 vs 0 doses) | Composite CVD/CeVD (“Cardiovascular disease”: coronary heart disease, stroke) | OR 0.58 (0.51–0.66) |
|  |  | Booster dose vs unvaccinated (≥3 vs 0 doses) | Composite CVD/CeVD (“Cardiovascular disease”: coronary heart disease, stroke) | OR 0.39 (0.34–0.44) |
|  | ≤21 days | Single dose vs unvaccinated (1 vs 0 doses) | Composite CVD/CeVD (“Cardiovascular disease”: coronary heart disease, stroke) | OR 0.99 (0.80–1.24) |
|  |  | Primary course vs unvaccinated (2 vs 0 doses) | Composite CVD/CeVD (“Cardiovascular disease”: coronary heart disease, stroke) | OR 0.61 (0.54–0.69) |
|  |  | Booster dose vs unvaccinated (≥3 vs 0 doses) | Composite CVD/CeVD (“Cardiovascular disease”: coronary heart disease, stroke) | OR 0.42 (0.37–0.47) |
|  | ≤28 days | Single dose vs unvaccinated (1 vs 0 doses) | Composite CVD/CeVD (“Cardiovascular disease”: coronary heart disease, stroke) | OR 0.96 (0.78–1.18) |
|  |  | Primary course vs unvaccinated (2 vs 0 doses) | Composite CVD/CeVD (“Cardiovascular disease”: coronary heart disease, stroke) | OR 0.62 (0.55–0.69) |
|  |  | Booster dose vs unvaccinated (≥3 vs 0 doses) | Composite CVD/CeVD (“Cardiovascular disease”: coronary heart disease, stroke) | OR 0.43 (0.39–0.48) |
|  | 1 month | Single dose vs unvaccinated (1 vs 0 doses) | Composite CVD/CeVD (“Cardiovascular disease”: coronary heart disease, stroke) | OR 0.94 (0.76–1.15) |
|  |  |  |  | HR 1.00 (0.87–1.14) |
|  |  |  | Coronary heart disease | OR 0.81 (0.58–1.13) |
|  |  |  |  | HR 0.84 (0.67–1.05) |
|  |  |  | Stroke | OR 1.10 (0.85–1.42) |
|  |  |  |  | HR 1.11 (0.94–1.32) |
|  |  | Primary course vs unvaccinated (2 vs 0 doses) | Composite CVD/CeVD (“Cardiovascular disease”: coronary heart disease, stroke) | OR 0.62 (0.56–0.70) |
|  |  |  |  | HR 0.70 (0.64–0.76) |
|  |  |  | Coronary heart disease | OR 0.52 (0.44–0.62) |
|  |  |  |  | HR 0.57 (0.50–0.65) |
|  |  |  | Stroke | OR 0.68 (0.59–0.78) |
|  |  |  |  | HR 0.77 (0.69–0.85) |
|  |  | Booster dose vs unvaccinated (≥3 vs 0 doses) | Composite CVD/CeVD (“Cardiovascular disease”: coronary heart disease, stroke) | OR 0.44 (0.40–0.49) |
|  |  |  |  | HR 0.50 (0.46–0.54) |
|  |  |  | Coronary heart disease | OR 0.22 (0.19–0.27) |
|  |  |  |  | HR 0.26 (0.22–0.30) |
|  |  |  | Stroke | OR 0.61 (0.53–0.69) |
|  |  |  |  | HR 0.67 (0.60–0.75) |
|  |  | Booster dose vs primary course (≥3 vs 2 doses) | Composite CVD/CeVD (“Cardiovascular disease”: coronary heart disease, stroke) | OR 0.70 (0.64–0.78) |
|  |  |  |  | HR 0.72 (0.66–0.78) |
|  |  |  | Coronary heart disease | OR 0.43 (0.35–0.52) |
|  |  |  |  | HR 0.45 (0.38–0.53) |
|  |  |  | Stroke | OR 0.89 (0.78–1.02) |
|  |  |  |  | HR 0.88 (0.79–0.97) |
| Sritharan 2024 | NR (during acute COVID-19 hospitalisation) | Vaccinated vs unvaccinated (≥1 vs 0 doses) | Composite CVD/VTE (“Clinical cardiovascular events” : new onset atrial fibrillation or flutter, high grade atrioventricular block, new cardiomyopathy heart failure, pericarditis, myocarditis or myopericarditis, pulmonary embolism) | OR 1.56 (0.59–4.16) |
| Yu 2024 | 22 to 90 days | Single dose vs unvaccinated (1 vs 0 doses) | Composite CVD/CeVD/VTE (“Cardiac-related post-acute sequelae of COVID-19”: Stroke, MI, heart failure, atrial fibrillation, coronary artery disease, myocarditis and pericarditis, deep vein thrombosis, cardiomyopathy, cardiovascular mortality) | OR 0.59 (0.29–1.19) |
|  |  | Primary course vs unvaccinated (2 vs 0 doses) | Composite CVD/CeVD/VTE (“Cardiac-related post-acute sequelae of COVID-19”: Stroke, MI, heart failure, atrial fibrillation, coronary artery disease, myocarditis and pericarditis, deep vein thrombosis, cardiomyopathy, cardiovascular mortality) | OR 0.45 (0.30–0.68) |
|  |  | Booster dose vs unvaccinated (≥3 vs 0 doses) | Composite CVD/CeVD/VTE (“Cardiac-related post-acute sequelae of COVID-19”: Stroke, MI, heart failure, atrial fibrillation, coronary artery disease, myocarditis and pericarditis, deep vein thrombosis, cardiomyopathy, cardiovascular mortality) | OR 0.46 (0.30–0.72) |
|  |  | Booster dose vs unvaccinated (≥3 vs 0 doses; BNT162b2 booster after CoronaVac primary course) | Composite CVD/CeVD/VTE (“Cardiac-related post-acute sequelae of COVID-19”: Stroke, MI, heart failure, atrial fibrillation, coronary artery disease, myocarditis and pericarditis, deep vein thrombosis, cardiomyopathy, cardiovascular mortality) | OR 0.23 (0.11–0.48) |
| El Seblani 2025 | 7 days | Vaccinated vs unvaccinated (2 vs 0 doses) | Intercranial haemorrhage | HR 0.66 (0.53–0.82) |
|  |  |  | Venous thromboembolic events | HR 0.44 (0.35–0.56) |
|  |  |  | Acute myocardial infarction | HR 0.58 (0.46–0.73) |
|  | 30 days | Vaccinated vs unvaccinated (2 vs 0 doses) | Intercranial haemorrhage | HR 0.66 (0.53–0.81) |
|  |  |  | Venous thromboembolic events | HR 0.51 (0.41–0.63) |
|  |  |  | Acute myocardial infarction | HR 0.60 (0.49–0.75) |
| Lin 2025 | NR | Vaccinated vs unvaccinated (≥2 vs 0 doses) | Composite CVD (“Cardiac complications”: acute myocardial infarction, other ischaemic heart disease, atrial fibrillation, ventricular fibrillation, other arrhythmias, cardiomyopathy, and congestive heart failure) | OR 0.66 (0.48–0.89) |
|  |  |  | Composite CVD/CeVD (Cardiovascular events”: cardiac complications along with pulmonary embolism, deep vein thrombosis, superficial vein thrombosis, other thrombosis, and cerebrovascular stroke) | OR 0.76 (0.59–0.99) |
| Meister 2025 | 365 days | Vaccinated vs unvaccinated (2 vs 0 doses) | Composite CVD/CeVD (“Major acute cardiovascular events”: acute myocardial infarction, stroke) | IRR 0.71 (0.58–0.84) |
| Montone 2025 | 30 days | Vaccinated vs unvaccinated (≥2 vs NR doses) | Myocardial injury (Elevation of hs-cTnI levels >99th percentile upper reference limit (>56 ng/L for a normal population) | OR 0.824 (0.556–1.222) |
| Wee 2025 | 31 to 365 days | Bivalent vs monovalent booster (3 vs 3 doses) | Composite CVD/CeVD/VTE (“All cardiovascular outcomes”: dysrhythmias, inflammatory heart disease, ischaemic heart disease, other heart disease (e.g. heart failure, cardiomyopathies), thrombotic disorders, cerebrovascular disease) | HR 0.77 (0.56–1.05)  HR 0.84 (0.61–1.16)^1^ |
|  |  |  | Dysrhythmias | HR 0.61 (0.33–1.11)  HR 0.59 (0.32–1.10)^1^ |
|  |  |  | Inflammatory heart disease | HR 0.28 (0.04–2.08)  HR 0.58 (0.07–4.67)^1^ |
|  |  |  | Ischaemic heart disease | HR 1.15 (0.71–1.85)  HR 1.27 (0.76–2.14)^1^ |
|  |  |  | Other heart disease (e.g. heart failure, cardiomyopathies) | HR 2.37 (1.14–4.93)  HR 1.91 (0.87–4.15)^1^ |
|  |  |  | Thrombotic disorders | HR 0.54 (0.29–0.99)  HR 0.59 (0.32–1.11)^1^ |
|  |  |  | Cerebrovascular disease | HR 0.66 (0.24–1.81)  HR 0.70 (0.25–1.94)^1^ |

^1^ Also includes time of infection (before/after Omicron XBB emergence), immunosuppression, prior chronic pulmonary disease in weighting.

CeVD, cerebrovascular disease; CI, confidence intervals; CVD, cardiovascular disease; HR, hazard ratio; IRR incidence rate ratio; L, litre; ng, nanograms; NR, not reported; OR, odds ratio; RR, relative risk or relative risk; VTE, venous thromboembolism.

#### Table 6 Vaccination impact on risk of CVD, CeVD, and VTE outcomes after SARS-CoV-2 infection by SARS-CoV-2 infection severity (adjusted risks or populations weighted/matched for confounding variables)

| **Study** | **Time from SARS-CoV-2 infection** | **Comparison** | **Outcome** | **COVID-19 severity** | **Risk estimate (95% CI)** |
| --- | --- | --- | --- | --- | --- |
| Al-Aly 2022 | 30 days to 6 months | Vaccinated vs unvaccinated (2 vs 0 doses) | Composite CVD (“Cardiovascular outcome”: Acute coronary disease, atrial fibrillation, heart failure, hypertension, myocardial infarction, myocarditis, other dysrhythmias, pericarditis, tachycardia) | Hospitalised | HR 0.90 (0.78–1.05) |
|  |  |  |  | ICU | HR 0.78 (0.63–0.98) |
|  |  |  |  | Non-hospitalised | HR 1.07 (0.98–1.17) |
| Kim 2022 | Median 90 (unvaccinated) and 84 (vaccinated) days (excluding first 30 days) | Vaccinated vs unvaccinated (2 vs 0 doses) | Composite CVD/CeVD: (“Composite cardiovascular events”: Acute myocardial infarction, ischaemic stroke) | Severe/critical | HR 0.66 (0.20–2.23) |
|  |  |  |  | Non-severe/critical | HR 0.37 (0.25–0.55) |
| Chen 2024 | 30 days | Vaccinated vs unvaccinated (≥1 vs 0 doses) | Composite CeVD (“Cerebrovascular diseases”: subarachnoid haemorrhage, intracerebral haemorrhage, other nontraumatic intracranial haemorrhage, cerebral infarction, occlusion and stenosis of precerebral arteries, occlusion and stenosis of cerebral arteries, other cerebrovascular diseases, cerebrovascular disorders in diseases classified elsewhere, sequalae of cerebrovascular disease) | Severe | HR 1.205 (0.926–1.568) |
|  |  |  |  | Non-severe | HR 0.700 (0.557–0.878) |
| Huh 2024 | 30 to 120 days | Vaccinated vs unvaccinated (2–3 vs 0 doses) | Ischaemic heart diseases | Non-severe | HR 0.73 (0.56–0.95) |
|  |  |  | Heart failure and cardiomyopathies | Non-severe | HR 0.53 (0.46–0.61) |
|  |  |  | Cardiac dysrhythmias | Non-severe | HR 0.72 (0.60–0.85) |
|  |  |  | Cardiac arrest | Non-severe | HR 0.38 (0.30–0.49) |
|  |  |  | Ischaemic stroke | Non-severe | HR 0.67 (0.52–0.87) |
|  |  |  | Haemorrhagic stroke | Non-severe | HR 0.53 (0.43–0.65) |
|  |  |  | Other cerebrovascular disease | Non-severe | HR 0.79 (0.60–1.03) |
|  |  |  | Venous thromboembolism | Non-severe | HR 0.92 (0.78–1.10) |
|  |  |  | Pulmonary embolism | Non-severe | HR 1.02 (0.94–1.10) |
|  |  | Booster dose vs primary course (3 vs 2 doses) | Ischaemic heart diseases | Non-severe | HR 0.86 (0.71–1.05) |
|  |  |  | Heart failure and cardiomyopathies | Non-severe | HR 0.82 (0.72–0.93) |
|  |  |  | Cardiac dysrhythmias | Non-severe | HR 0.85 (0.75–0.98) |
|  |  |  | Cardiac arrest | Non-severe | HR 0.75 (0.58–0.97) |
|  |  |  | Ischaemic stroke | Non-severe | HR 0.77 (0.63–0.94) |
|  |  |  | Haemorrhagic stroke | Non-severe | HR 0.89 (0.76–1.06) |
|  |  |  | Other cerebrovascular disease | Non-severe | HR 0.87 (0.70–1.08) |
|  |  |  | Venous thromboembolism | Non-severe | HR 1.01 (0.89–1.14) |
|  |  |  | Pulmonary embolism | Non-severe | HR 1.02 (0.94–1.12) |
| Meister 2025 | 365 days | Vaccinated vs unvaccinated (2 vs 0 doses) | Composite CVD/CeVD (“Major acute cardiovascular events”: acute myocardial infarction, stroke) | Non-severe | IRR 0.73 (0.59–0.87) |

CeVD, cerebrovascular disease; CI, confidence intervals; CVD, cardiovascular disease; HR, hazard ratio; ICU, intensive care unit; IRR, incidence rate ratio; VTE, venous thromboembolism.

#### Table 7 Vaccination impact on risk of CVD, CeVD, and VTE outcomes after SARS-CoV-2 infection by SARS-CoV-2 variant (adjusted risks or populations weighted/matched for confounding variables)

| **Study** | **Time from SARS-CoV-2 infection** | **Comparison** | **Outcome** | **COVID-19 variant** | **Risk estimate (95% CI)** |
| --- | --- | --- | --- | --- | --- |
| Chen 2024 | 30 days | Vaccinated vs unvaccinated (≥1 vs 0 doses) | Composite CeVD (“Cerebrovascular diseases”: subarachnoid haemorrhage, intracerebral haemorrhage, other nontraumatic intracranial haemorrhage, cerebral infarction, occlusion and stenosis of precerebral arteries, occlusion and stenosis of cerebral arteries, other cerebrovascular diseases, cerebrovascular disorders in diseases classified elsewhere, sequalae of cerebrovascular disease) | Delta | HR 0.512 (0.315–0.834) |
|  |  |  |  | Omicron | HR 0.685 (0.531–0.883) |
|  |  |  | Intracerebral haemorrhage | Delta | HR 0.191 (0.022–1.636) |
|  |  |  |  | Omicron | HR 0.823 (0.298–2.269) |
|  |  |  | Cerebral infarction | Delta | HR 0.605 (0.262–1.399) |
|  |  |  |  | Omicron | HR 0.665 (0.452–0.979) |
|  |  |  | Occlusion and stenosis of precerebral arteries | Delta | HR 0.578 (0.240–1.395) |
|  |  |  |  | Omicron | HR 0.937 (0.594–1.478) |
|  |  |  | Other cerebrovascular diseases | Delta | HR 0.221 (0.074–0.656) |
|  |  |  |  | Omicron | HR 0.489 (0.295–0.809) |
|  |  |  | Sequalae of cerebrovascular disease | Delta | HR 1.285 (0.287–5.741) |
|  |  |  |  | Omicron | HR 1.203 (0.448–3.230) |
| Madrid 2024 | NR (during acute COVID-19 hospitalisation) | Vaccinated vs unvaccinated (2 vs 0 doses) | Composite CVD (“Acute cardiac events”: myocardial infarction, heart failure, arrhythmia, myocarditis) | Delta | RR 0.26 (0.05–0.64) |
| O’Carroll 2024 | 90 days | Vaccinated vs unvaccinated (2 vs 0 doses) | Venous thromboembolism | Delta | OR 0.12 (0.03–0.42) |
|  |  |  |  | Omicron | OR 0.28 (0.02–4.88) |
| El Seblani 2025 | 7 days | Vaccinated vs unvaccinated (2 vs 0 doses) | Intercranial haemorrhage | Delta | HR 0.52 (0.37–0.73) |
|  |  |  |  | Omicron | HR 0.79 (0.59–1.04) |
|  |  |  | Venous thromboembolic events | Delta | HR 0.33 (0.23–0.48) |
|  |  |  |  | Omicron | HR 0.44 (0.33–0.60) |
|  |  |  | Myocardial infarction | Delta | HR 0.50 (0.36–0.70) |
|  |  |  |  | Omicron | HR 0.51 (0.38–0.69) |
|  | 30 days | Vaccinated vs unvaccinated (2 vs 0 doses) | Intercranial haemorrhage | Delta | HR 0.51 (0.37–0.71) |
|  |  |  |  | Omicron | HR 0.83 (0.63–1.09) |
|  |  |  | Venous thromboembolic events | Delta | HR 0.36 (0.26–0.50) |
|  |  |  |  | Omicron | HR 0.52 (0.40–0.68) |
|  |  |  | Myocardial infarction | Delta | HR 0.52 (0.38–0.71) |
|  |  |  |  | Omicron | HR 0.55 (0.41–0.72) |
| Lin 2025 | NR | Vaccinated vs unvaccinated (≥2 vs 0 doses) | Composite CVD (“Cardiac complications”: acute myocardial infarction, other ischaemic heart disease, atrial fibrillation, ventricular fibrillation, other arrhythmias, cardiomyopathy, and congestive heart failure)  Composite CVD (“Cardiac complications”: acute myocardial infarction, other ischaemic heart disease, atrial fibrillation, ventricular fibrillation, other arrhythmias, cardiomyopathy, and congestive heart failure) | Alpha | OR 0.53 (0.25–1.13) |
|  |  |  |  | Delta | OR 0.70 (0.48–1.04) |
|  |  |  |  | Omicron | OR 0.57 (0.31–1.07) |
|  |  |  | Composite CVD/CeVD (Cardiovascular events”: cardiac complications along with pulmonary embolism, deep vein thrombosis, superficial vein thrombosis, other thrombosis, and cerebrovascular stroke) | Alpha | OR 0.50 (0.25–0.99) |
|  |  |  |  | Delta | OR 0.75 (0.54–1.04) |
|  |  |  |  | Omicron | OR 0.88 (0.50–1.55) |

CeVD, cerebrovascular disease; CI, confidence intervals; CVD, cardiovascular disease; HR, hazard ratio; NR, not reported; OR, odds ratio; VTE, venous thromboembolism.

#### Table 8 Vaccination impact on risk of CVD, CeVD, and VTE outcomes after SARS-CoV-2 infection by vaccine (adjusted risks or populations weighted/matched for confounding variables)

| **Study** | **Time from SARS-CoV-2 infection** | **Comparison** | **Outcome** | **Vaccine** | **Risk estimate (95% CI)** |
| --- | --- | --- | --- | --- | --- |
| Kim 2024 | NR | Vaccinated vs unvaccinated (≥2 vs 0-1 doses) | Pulmonary embolism | BNT162b2, mRNA-1273 | HR 0.12 (0.07–0.20) |
|  |  |  |  | ChAdOx1, AD26.COV2-S | HR 0.17 (0.10–0.29) |
|  |  |  | Deep vein thrombosis | BNT162b2, mRNA-1273 | HR 0.33 (0.19–0.56) |
|  |  |  |  | ChAdOx1, AD26.COV2-S | HR 0.31 (0.18–0.56) |
| El Seblani 2025 | 7 days | Vaccinated vs unvaccinated (2 vs 0 doses) | Intercranial haemorrhage | BNT162b2 | HR 0.62 (0.48–0.81) |
|  |  |  |  | mRNA-1273 | HR 0.85 (0.60–1.30) |
|  |  |  | Venous thromboembolism | BNT162b2 | HR 0.44 (0.35–0.56) |
|  |  |  |  | mRNA-1273 | HR 0.85 (0.56–1.31) |
|  |  |  | Myocardial infarction | BNT162b2 | HR 0.58 (0.46–0.73) |
|  |  |  |  | mRNA-1273 | HR 0.74 (0.51–1.08) |
|  | 30 days | Vaccinated vs unvaccinated (2 vs 0 doses) | Intercranial haemorrhage | BNT162b2 | HR 0.65 (0.50–0.84) |
|  |  |  |  | mRNA-1273 | HR 0.82 (0.56–1.20) |
|  |  |  | Venous thromboembolism | BNT162b2 | HR 0.51 (0.41–0.63) |
|  |  |  |  | mRNA-1273 | HR 0.79 (0.54–1.16) |
|  |  |  | Myocardial infarction | BNT162b2 | HR 0.60 (0.49–0.75) |
|  |  |  |  | mRNA-1273 | HR 0.81 (0.57–1.16) |

CeVD, cerebrovascular disease; CI, confidence intervals; CVD, cardiovascular disease; HR, hazard ratio; NR, not reported; VTE, venous thromboembolism..

#### Table 9 Vaccination impact on risk of CVD, CeVD, and VTE outcomes after SARS-CoV-2 infection by prior CVD/CeVD/VTE disease status (adjusted risks or populations weighted/matched for confounding variables)

| **Study** | **Time from SARS-CoV-2 infection** | **Comparison** | **Outcome** | **Prior CVD/CeVD/VTE status** | **Risk estimate (95% CI)** |
| --- | --- | --- | --- | --- | --- |
| Kim 2022 | Median 90 (unvaccinated) and 84 (vaccinated) days (excluding first 30 days) | Vaccinated vs unvaccinated (≥2 vs 0-1 doses) | Composite CVD/CeVD: (“Composite cardiovascular events”: Acute myocardial infarction, ischaemic stroke) | Previous history of outcome | HR 0.33 (0.10–1.07) |
|  |  |  |  | No previous history of outcome | HR 0.44 (0.29–0.65) |

CeVD, cerebrovascular disease; CI, confidence intervals; CVD, cardiovascular disease; VTE, venous thromboembolism.

#### Table 10 Vaccination impact on risk of CVD, CeVD, and VTE outcomes after SARS-CoV-2 infection by sex (adjusted risks or populations weighted/matched for confounding variables)

| **Study** | **Time from SARS-CoV-2 infection** | **Comparison** | **Outcome** | **Sex** | **Risk estimate (95% CI)** |
| --- | --- | --- | --- | --- | --- |
| Kim 2022 | Median 90 (unvaccinated) and 84 (vaccinated) days (excluding first 30 days) | Vaccinated vs unvaccinated (2 vs 0 doses) | Composite CVD/CeVD: (“Composite cardiovascular events”: Acute myocardial infarction, ischaemic stroke) | Male | HR 0.41 (0.26–0.66) |
|  |  |  |  | Female | HR 0.42 (0.23–0.76) |
| Chen 2024 | 30 days | Vaccinated vs unvaccinated (≥1 vs 0 doses) | Composite CeVD (“Cerebrovascular diseases”: subarachnoid haemorrhage, intracerebral haemorrhage, other nontraumatic intracranial haemorrhage, cerebral infarction, occlusion and stenosis of precerebral arteries, occlusion and stenosis of cerebral arteries, other cerebrovascular diseases, cerebrovascular disorders in diseases classified elsewhere, sequalae of cerebrovascular disease) | Male | HR 0.566 (0.453–0.707) |
|  |  |  |  | Female | HR 0.616 (0.496–0.765) |
| Meister 2024 | 365 days | Vaccinated vs unvaccinated (2 vs 0 doses) | Composite CVD/CeVD (“Major acute cardiovascular events”: acute myocardial infarction, stroke) | Male | IRR 1.03 (0.78–1.27) |
|  |  |  |  | Female | IRR 0.56 (0.41–0.71) |
| Song 2024 | 1 month | Single dose vs unvaccinated (1 vs 0 doses) | Composite CVD/CeVD (“Major acute cardiovascular events”: acute myocardial infarction, stroke) | Male | OR 0.94 (0.71–1.25) |
|  |  |  |  | Female | OR 0.91 (0.67–1.24) |
|  |  | Primary course vs unvaccinated (2 vs 0 doses) | Composite CVD/CeVD (“Major acute cardiovascular events”: acute myocardial infarction, stroke) | Male | OR 0.65 (0.56–0.76) |
|  |  |  |  | Female | OR 0.60 (0.51–0.70) |
|  |  | Booster dose vs unvaccinated (≥3 vs 0 doses) | Composite CVD/CeVD (“Major acute cardiovascular events”: acute myocardial infarction, stroke) | Male | OR 0.43 (0.37–0.50) |
|  |  |  |  | Female | OR 0.46 (0.40–0.53) |
| Wee 2025 | 31 to 365 days | Bivalent vs monovalent booster (3 vs 3 doses) | Composite CVD/CeVD/VTE (“All cardiovascular outcomes”: dysrhythmias, inflammatory heart disease, ischaemic heart disease, other heart disease (e.g. heart failure, cardiomyopathies), thrombotic disorders, cerebrovascular disease) | Male | HR 0.667 (0.432–1.031) |
|  |  |  |  | Female | HR 0.902 (0.577–1.408) |
|  |  |  | Dysrhythmias | Male | HR 0.495 (0.202–1.215) |
|  |  |  |  | Female | HR 0.729 (0.32–1.66) |
|  |  |  | Ischaemic heart disease | Male | HR 1.032 (0.523–2.037) |
|  |  |  |  | Female | HR 1.247 (0.634–2.451) |
|  |  |  | Other heart disease (e.g. heart failure, cardiomyopathies) | Male | HR 1.032 (0.523–2.037) |
|  |  |  |  | Female | HR 2.015 (0.664–6.119) |
|  |  |  | Thrombotic disorders | Male | HR 0.471 (0.206–1.076) |
|  |  |  |  | Female | HR 0.656 (0.269–1.598) |

CeVD, cerebrovascular disease; CI, confidence intervals; CVD, cardiovascular disease; HR, hazard ratio; IRR, incidence rate ratio; OR, odds ratio; VTE, venous thromboembolism.

#### Table 11 Vaccination impact on risk of CVD, CeVD, and VTE outcomes after SARS-CoV-2 infection by comorbidity status (adjusted risks or populations weighted/matched for confounding variables)

| **Study** | **Time from SARS-CoV-2 infection** | **Comparison** | **Outcome** | **Comorbidity status** | **Risk estimate (95% CI)** |
| --- | --- | --- | --- | --- | --- |
| Kim 2022 | Median 90 (unvaccinated) and 84 (vaccinated) days (excluding first 30 days) | Vaccinated vs unvaccinated (2 vs 0 doses) | Composite CVD/CeVD: (“Composite cardiovascular events”: Acute myocardial infarction, ischaemic stroke) | ≥5 CCI score | HR 0.54 (0.24–1.22) |
|  |  |  |  | <5 CCI score | HR 0.40 (0.26–0.60) |
|  |  |  |  | Diabetes | HR 0.47 (0.25–0.91) |
|  |  |  |  | No diabetes | HR 0.38 (0.24–0.61) |
|  |  |  |  | Hypertension | HR 0.34 (0.18–0.62) |
|  |  |  |  | No hypertension | HR 0.50 (0.31–0.80) |
|  |  |  |  | Dyslipidaemia | HR 0.09 (0.03–0.34) |
|  |  |  |  | No dyslipidaemia | HR 0.54 (0.37–0.80) |
| Chen 2024 | 30 days | Vaccinated vs unvaccinated (≥1 vs 0 doses) | Composite CeVD (“Cerebrovascular diseases”: subarachnoid haemorrhage, intracerebral haemorrhage, other nontraumatic intracranial haemorrhage, cerebral infarction, occlusion and stenosis of precerebral arteries, occlusion and stenosis of cerebral arteries, other cerebrovascular diseases, cerebrovascular disorders in diseases classified elsewhere, sequalae of cerebrovascular disease) | Obesity | HR 0.848 (0.616–1.166) |
|  |  |  |  | No obesity | HR 0.617 (0.494–0.770) |
|  |  |  |  | Diabetes | HR 0.701 (0.539–0.912) |
|  |  |  |  | No diabetes | HR 0.647 (0.525–0.796) |
|  |  |  |  | Chronic kidney disease | HR 0.808 (0.592–1.102) |
|  |  |  |  | No chronic kidney disease | HR 0.607 (0.501–0.736) |
|  |  |  |  | Hypertensive diseases | HR 0.777 (0.644–0.938) |
|  |  |  |  | No hypertensive diseases | HR 0.582 (0.391–0.865) |
| Madrid 2024 | NR (during acute COVID-19 hospitalisation) | Vaccinated vs unvaccinated (2 vs 0 doses) | Composite CVD (“Acute cardiac events”: myocardial infarction, heart failure, arrhythmia, myocarditis) | Non-healthy subjects | RR 0.22 (0.03–0.54) |
|  |  |  |  | No cancer | RR 0.18 (0.01–0.59) |
|  |  |  |  | No vascular disease | RR 0.33 (0.05–0.80) |
|  |  |  |  | No neurological disease | RR 0.37 (0.06–0.91) |
|  |  |  |  | No organ transplant | RR 0.45 (0.09–0.96) |
| Song 2024 | 1 month | Single dose vs unvaccinated (1 vs 0 doses) | Composite CVD/CeVD (“Cardiovascular disease”: coronary heart disease, stroke) | Hypertension | OR 1.43 (0.64–3.18) |
|  |  |  |  | Diabetes | OR 1.18 (0.66–2.12) |
|  |  |  |  | Cancer | OR 1.18 (0.48–2.89) |
|  |  | Primary course vs unvaccinated (2 vs 0 doses) | Composite CVD/CeVD (“Cardiovascular disease”: coronary heart disease, stroke) | Hypertension | OR 0.69 (0.42–1.15) |
|  |  |  |  | Diabetes | OR 0.62 (0.43–0.88) |
|  |  |  |  | Cancer | OR 0.72 (0.43–1.22) |
|  |  | Booster dose vs unvaccinated (≥3 vs 0 doses) | Composite CVD/CeVD (“Cardiovascular disease”: coronary heart disease, stroke) | Hypertension | OR 0.53 (0.33–0.85) |
|  |  |  |  | Diabetes | OR 0.35 (0.25–0.50) |
|  |  |  |  | Cancer | OR 0.28 (0.16–0.48) |
| Wee 2025 | 31 to 365 days | Bivalent vs monovalent booster (3 vs 3 doses) | Composite CVD/CeVD/VTE (“All cardiovascular outcomes”: dysrhythmias, inflammatory heart disease, ischaemic heart disease, other heart disease (e.g. heart failure, cardiomyopathies), thrombotic disorders, cerebrovascular disease) | 0 CCI score | HR 0.681 (0.453–1.023) |
|  |  |  |  | ≥1 CCI score | HR 1.004 (0.624–1.616) |
|  |  |  | Dysrhythmias | 0 CCI score | HR 0.630 (0.229–1.736) |
|  |  |  |  | ≥1 CCI score | HR 0.627 (0.296–1.326) |
|  |  |  | Ischaemic heart disease | 0 CCI score | HR 0.862 (0.481–1.544) |
|  |  |  |  | ≥1 CCI score | HR 2.562 (1.131–5.802) |
|  |  |  | Other heart disease (e.g. heart failure, cardiomyopathies) | 0 CCI score | HR 1.618 (0.440–5.948) |
|  |  |  |  | ≥1 CCI score | HR 3.106 (1.285–7.505) |
|  |  |  | Thrombotic disorders | 0 CCI score | HR 0.423 (0.183–0.976) |
|  |  |  |  | ≥1 CCI score | HR 0.787 (0.332–1.864) |
|  |  |  | Cerebrovascular disease | 0 CCI score | HR 0.660 (0.158–2.764) |
|  |  |  |  | ≥1 CCI score | HR 0.754 (0.187–3.043) |

CCI, Charlson Comorbidity Index; CeVD, cerebrovascular disease; CI, confidence intervals; CVD, cardiovascular disease; HR, hazard ratio; NR, not reported; OR, odds ratio; RR, relative risk or relative risk; VTE, venous thromboembolism.

#### Table 12 Vaccination impact on risk of CVD, CeVD, and VTE outcomes after SARS-CoV-2 infection by immunocompromised status (adjusted risks or populations weighted/matched for confounding variables)

| **Study** | **Time from SARS-CoV-2 infection** | **Comparison** | **Outcome** | **Immunocompromised status** | **Risk estimate (95% CI)** |
| --- | --- | --- | --- | --- | --- |
| Al-Aly 2022 | 30 days to 6 months | Vaccinated vs unvaccinated (2 vs 0 doses) | Composite CVD (“Cardiovascular outcome”: Acute coronary disease, atrial fibrillation, heart failure, hypertension, myocardial infarction, myocarditis, other dysrhythmias, pericarditis, tachycardia) | Immunocompromised | HR 0.78 (0.63–0.97) |
|  |  |  |  | Not immunocompromised | HR 0.92 (0.85–1.00) |
| Madrid 2024 | NR (during acute COVID-19 hospitalisation) | Vaccinated vs unvaccinated (2 vs 0 doses) | Composite CVD (“Acute cardiac events”: myocardial infarction, heart failure, arrhythmia, myocarditis) | Immunocompromised | RR 0.46 (0.09–0.99) |

CeVD, cerebrovascular disease; CI, confidence intervals; CVD, cardiovascular disease; HR, hazard ratio; NR, not reported; RR, relative risk or relative risk; VTE, venous thromboembolism.

#### Table 13 Vaccination impact on risk of CVD, CeVD, and VTE outcomes after SARS-CoV-2 infection by age (adjusted risks or populations weighted/matched for confounding variables)

| **Study** | **Time from SARS-CoV-2 infection** | **Comparison** | **Outcome** | **Age group, years** | **Risk estimate (95% CI)** |
| --- | --- | --- | --- | --- | --- |
| Kim 2022 | Median 90 (unvaccinated) and 84 (vaccinated) days (excluding first 30 days) | Vaccinated vs unvaccinated (2 vs 0 doses) | Composite CVD/CeVD: (“Composite cardiovascular events”: Acute myocardial infarction, ischaemic stroke) | 40–65 | HR 0.38 (0.20–0.74) |
|  |  |  |  | ≥65 | HR 0.41 (0.26–0.66) |
| Taquet 2022 | 6 months | Vaccinated vs unvaccinated (≥1 vs 0 doses) | Arrhythmia | <60 | HR 0.97 (0.81–1.15) |
|  |  |  |  | ≥60 | HR 1.07 (0.97–1.19) |
|  |  |  | Cardiac failure | <60 | HR 0.93 (0.72–1.22) |
|  |  |  |  | ≥60 | HR 1.16 (1.02–1.33) |
|  |  |  | Cardiomyopathy | <60 | HR 1.49 (1.05–2.13) |
|  |  |  |  | ≥60 | HR 1.02 (0.81–1.28) |
|  |  |  | Cerebral haemorrhage | <60 | HR 0.43 (0.19–0.99) |
|  |  |  |  | ≥60 | HR 1.33 (0.77–2.31) |
|  |  |  | Coronary disease | <60 | HR 0.63 (0.39–1.01) |
|  |  |  |  | ≥60 | HR 1.34 (1.07–1.67) |
|  |  |  | Hypercoagulopathy/deep vein thrombosis/pulmonary embolism | <60 | HR 0.71 (0.55–0.91) |
|  |  |  |  | ≥60 | HR 1.04 (0.87–1.23) |
|  |  |  | Hypertension | <60 | HR 1.00 (0.90–1.11) |
|  |  |  |  | ≥60 | HR 1.11 (1.04–1.19) |
|  |  |  | Ischaemic stroke | <60 | HR 1.05 (0.64–1.72) |
|  |  |  |  | ≥60 | HR 1.03 (0.80–1.33) |
|  |  |  | Myocarditis | <60 | HR 3.81 (0.79–18.39) |
|  |  |  |  | ≥60 | HR 0.45 (0.14–1.48) |
| Chen 2024 | 30 days | Vaccinated vs unvaccinated (≥1 vs 0 doses) | Composite CeVD (“Cerebrovascular diseases”: subarachnoid haemorrhage, intracerebral haemorrhage, other nontraumatic intracranial haemorrhage, cerebral infarction, occlusion and stenosis of precerebral arteries, occlusion and stenosis of cerebral arteries, other cerebrovascular diseases, cerebrovascular disorders in diseases classified elsewhere, sequalae of cerebrovascular disease) | 18–29 | HR 0.706 (0.158–3.156) |
|  |  |  |  | 30–39 | HR 0.571 (0.237–1.377) |
|  |  |  |  | 40–49 | HR 0.506 (0.264–0.970) |
|  |  |  |  | 50–59 | HR 0.600 (0.388–0.929) |
|  |  |  |  | 60–69 | HR 0.716 (0.537–0.956) |
|  |  |  |  | 70–79 | HR 0.592 (0.452–0.777) |
|  |  |  |  | ≥80 | HR 0.867 (0.635–1.185) |
| Huh 2024 | 30 to 120 days | Vaccinated vs unvaccinated (2-3 vs 0 doses) | Ischaemic heart diseases | ≥65 | HR 0.62 (0.43–0.90) |
|  |  |  | Heart failure and cardiomyopathies | ≥65 | HR 0.61 (0.50–0.75) |
|  |  |  | Cardiac dysrhythmias | ≥65 | HR 0.93 (0.71–1.20) |
|  |  |  | Cardiac arrest | ≥65 | HR 0.44 (0.33–0.59) |
|  |  |  | Pulmonary embolism | ≥65 | HR 0.80 (0.54–1.19) |
|  |  |  | Venous thromboembolism | ≥65 | HR 0.69 (0.45–1.05) |
|  |  |  | Haemorrhagic stroke | ≥65 | HR 0.94 (0.63–1.41) |
|  |  |  | Ischaemic stroke | ≥65 | HR 0.97 (0.77–1.22) |
|  |  |  | Other cerebrovascular diseases | ≥65 | HR 0.90 (0.70–1.14) |
|  |  | Booster dose vs primary course (3 vs 2 doses) | Ischaemic heart diseases | ≥65 | HR 0.71 (0.51–0.99) |
|  |  |  | Heart failure and cardiomyopathies | ≥65 | HR 0.83 (0.69–1.00) |
|  |  |  | Cardiac dysrhythmias | ≥65 | HR 0.85 (0.69–1.05) |
|  |  |  | Cardiac arrest | ≥65 | HR 0.81 (0.58–1.12) |
|  |  |  | Pulmonary embolism | ≥65 | HR 0.81 (0.58–1.14) |
|  |  |  | Venous thromboembolism | ≥65 | HR 0.95 (0.62–1.46) |
|  |  |  | Haemorrhagic stroke | ≥65 | HR 0.89 (0.61–1.29) |
|  |  |  | Ischaemic stroke | ≥65 | HR 1.05 (0.86–1.28) |
|  |  |  | Other cerebrovascular diseases | ≥65 | HR 1.10 (0.90–1.34) |
| Madrid 2024 | NR (during acute COVID-19 hospitalisation) | Vaccinated vs unvaccinated (2 vs 0 doses) | Composite CVD (“Acute cardiac events”: myocardial infarction, heart failure, arrhythmia, myocarditis) | <60 | RR 0.16 (0.00–0.77) |
|  |  |  |  | >60 | RR 0.38 (0.02–1.62) |
| Song 2024 | 1 month | Single dose vs unvaccinated (1 vs 0 doses) | Composite CVD/CeVD (“Cardiovascular disease”: coronary heart disease, stroke) | 20–39 | OR 0.85 (0.25–2.90) |
|  |  |  |  | 40–49 | OR 0.58 (0.21–1.64) |
|  |  |  |  | 50–59 | OR 0.81 (0.48–1.38) |
|  |  |  |  | 60–69 | OR 0.87 (0.60–1.25) |
|  |  |  |  | 70–80 | OR 0.85 (0.55–1.33) |
|  |  |  |  | ≥80 | OR 1.20 (0.78–1.83) |
|  |  | Primary course vs unvaccinated (2 vs 0 doses) | Composite CVD/CeVD (“Cardiovascular disease”: coronary heart disease, stroke) | 20–39 | OR 0.40 (0.21–0.76) |
|  |  |  |  | 40–49 | OR 0.46 (0.29–0.74) |
|  |  |  |  | 50–59 | OR 0.51 (0.38–0.69) |
|  |  |  |  | 60–69 | OR 0.57 (0.46–0.72) |
|  |  |  |  | 70–80 | OR 0.70 (0.56–0.86) |
|  |  |  |  | ≥80 | OR 0.84 (0.68–1.04) |
|  |  | Booster dose vs unvaccinated (≥3 vs 0 doses) | Composite CVD/CeVD (“Cardiovascular disease”: coronary heart disease, stroke) | 20–39 | OR 0.41 (0.20–0.88) |
|  |  |  |  | 40–49 | OR 0.44 (0.27–0.72) |
|  |  |  |  | 50–59 | OR 0.55 (0.42–0.72) |
|  |  |  |  | 60–69 | OR 0.36 (0.30–0.45) |
|  |  |  |  | 70–80 | OR 0.34 (0.28–0.42) |
|  |  |  |  | ≥80 | OR 0.54 (0.45–0.65) |
| Meister 2025 | 365 days | Vaccinated vs unvaccinated (2 vs 0 doses) | Composite CVD/CeVD (“Major acute cardiovascular events”: acute myocardial infarction, stroke) | ≤70, male | IRR 0.70 (0.50–0.91) |
|  |  |  |  | >70, male | IRR 1.66 (0.95–2.37) |
|  |  |  |  | ≤70, female | IRR 0.46 (0.27–0.64) |
|  |  |  |  | >70, female | IRR 0.60 (0.39–0.82) |
| Montone 2025 | 30 days | Vaccinated vs unvaccinated (≥2 vs NR doses) | Myocardial injury (Elevation of hs-cTnI levels >99th percentile upper reference limit (>56 ng/L for a normal population) | ≤60 | OR 4.438 (1.284–15.344) |
|  |  |  |  | 61-75 | OR 1.248 (0.662–2.351) |
|  |  |  |  | ≥75 | OR 0.567 (0.341–0.945) |
| Wee 2025 | 31 to 365 days | Bivalent vs monovalent booster (3 vs 3 doses) | Composite CVD/CeVD/VTE (“All cardiovascular outcomes”: dysrhythmias, inflammatory heart disease, ischaemic heart disease, other heart disease (e.g. heart failure, cardiomyopathies), thrombotic disorders, cerebrovascular disease) | <60 | HR 0.374 (0.218–0.641) |
|  |  |  |  | ≥60 | HR 1.099 (0.769–1.571) |
|  |  |  | Dysrhythmias | <60 | HR 0.253 (0.060–1.062) |
|  |  |  |  | ≥60 | HR 0.639 (0.336–1.216) |
|  |  |  | Ischaemic heart disease | <60 | HR 0.471 (0.223–0.994) |
|  |  |  |  | ≥60 | HR 3.028 (1.691–5.422) |
|  |  |  | Other heart disease (e.g. heart failure, cardiomyopathies) | <60 | HR 6.643 (1.203–36.67) |
|  |  |  |  | ≥60 | HR 1.350 (0.486–3.752) |
|  |  |  | Thrombotic disorders | <60 | HR 0.065 (0.009–0.473) |
|  |  |  |  | ≥60 | HR 0.868 (0.471–1.600) |
|  |  |  | Cerebrovascular disease | <60 | HR 0.660 (0.075–5.808) |
|  |  |  |  | ≥60 | HR 0.617 (0.201–1.897) |

CeVD, cerebrovascular disease; CI, confidence intervals; CVD, cardiovascular disease; HR, hazard ratio; IRR, incidence rate ratio; NR, not reported; OR, odds ratio; RR, relative risk or relative risk; VTE, venous thromboembolism.

#### Table 14 Risk of CVD, CeVD, and VTE events after SARS-CoV-2 infection, stratified by vaccination status (adjusted risks or populations weighted/matched for confounding variables)

| **Study** | **Time from SARS-CoV-2 infection** | **Outcome** | **Vaccination status** | **Risk estimate (95% CI)** |
| --- | --- | --- | --- | --- |
| Xie 2022 | 30 days | Venous thromboembolism | Vaccinated (2 doses) | HR 5.95 (1.82–19.51) |
|  |  |  | Unvaccinated/partially vaccinated (0-1 doses) | HR 27.94 (15.11–51.65) |
| Cezard 2024 | 0 days | Composite CVD/CeVD (“All arterial thrombotic events”: acute myocardial infarction, ischaemic stroke, composite ‘arterial thrombotic event’) | Vaccinated | HR 71.8 (66.9–77.0) |
|  |  |  | Unvaccinated | HR 198.5 (168.8–233.4) |
|  |  | Acute myocardial infarction | Vaccinated | HR 64.5 (58.0–71.7) |
|  |  |  | Unvaccinated | HR 144.9 (111.8–187.8) |
|  |  | Ischaemic stroke | Vaccinated | HR 79.9 (72.7–87.7) |
|  |  |  | Unvaccinated | HR 212.0 (168.7–266.4) |
|  |  | Heart failure | Vaccinated | HR 74.4 (70.9–78.1) |
|  |  |  | Unvaccinated | HR 181.0 (157.9–207.5) |
|  |  | Angina | Vaccinated | HR 45.5 (42.3–48.9) |
|  |  |  | Unvaccinated | HR 144.5 (120.4–173.5) |
|  |  | Composite VTE (“All venous thrombotic events”: pulmonary embolism, deep vein thrombosis, composite ‘venous thrombotic event’) | Vaccinated | HR 72.3 (66.2–79.0) |
|  |  |  | Unvaccinated | HR 382.6 (336.8–434.6) |
|  |  | Pulmonary embolism | Vaccinated | HR 137.3 (123.7–152.4) |
|  |  |  | Unvaccinated | HR 1151.7 (990.9–1338.5) |
|  |  | Deep vein thrombosis | Vaccinated | HR 27.3 (22.5–33.0) |
|  |  |  | Unvaccinated | HR 75.5 (55.4–103.0) |
|  | 1 to 4 weeks | Composite CVD/CeVD (“All arterial thrombotic events”: acute myocardial infarction, ischaemic stroke, composite ‘arterial thrombotic event’) | Vaccinated | HR 2.09 (1.92–2.28) |
|  |  |  | Unvaccinated | HR 8.53 (7.20–10.1) |
|  |  | Acute myocardial infarction | Vaccinated | HR 1.97 (1.74–2.23) |
|  |  |  | Unvaccinated | HR 7.27 (5.65–9.35) |
|  |  | Ischaemic stroke | Vaccinated | HR 2.12 (1.88–2.39) |
|  |  |  | Unvaccinated | HR 7.40 (5.70–9.59) |
|  |  | Heart failure | Vaccinated | HR 2.55 (2.41–2.70) |
|  |  |  | Unvaccinated | HR 7.74 (6.56–9.13) |
|  |  | Angina | Vaccinated | HR 2.09 (1.95–2.25) |
|  |  |  | Unvaccinated | HR 6.33 (5.15–7.78) |
|  |  | Composite VTE (“All venous thrombotic events”: pulmonary embolism, deep vein thrombosis, composite ‘venous thrombotic event’) | Vaccinated | HR 4.87 (4.53–5.23) |
|  |  |  | Unvaccinated | HR 29.6 (26.7–32.9) |
|  |  | Pulmonary embolism | Vaccinated | HR 9.10 (8.36–9.90) |
|  |  |  | Unvaccinated | HR 82.8 (72.7–94.3) |
|  |  | Deep vein thrombosis | Vaccinated | HR 2.37 (2.07–2.72) |
|  |  |  | Unvaccinated | HR 6.49 (5.16–8.17) |
|  | 5 to 28 weeks | Composite CVD/CeVD (“All arterial thrombotic events”: acute myocardial infarction, ischaemic stroke, composite ‘arterial thrombotic event’) | Vaccinated | HR 1.08 (1.00–1.18) |
|  |  |  | Unvaccinated | HR 1.54 (1.16–2.04) |
|  |  | Acute myocardial infarction | Vaccinated | HR 1.05 (0.93–1.18) |
|  |  |  | Unvaccinated | HR 1.61 (1.11–2.35) |
|  |  | Ischaemic stroke | Vaccinated | HR 1.09 (0.96–1.22) |
|  |  |  | Unvaccinated | HR 1.27 (0.81–1.98) |
|  |  | Heart failure | Vaccinated | HR 1.14 (1.07–1.21) |
|  |  |  | Unvaccinated | HR 1.87 (1.47–2.40) |
|  |  | Angina | Vaccinated | HR 1.12 (1.04–1.20) |
|  |  |  | Unvaccinated | HR 1.34 (0.97–1.86) |
|  |  | Composite VTE (“All venous thrombotic events”: pulmonary embolism, deep vein thrombosis, composite ‘venous thrombotic event’) | Vaccinated | HR 1.53 (1.39–1.67) |
|  |  |  | Unvaccinated | HR 2.36 (1.87–2.99) |
|  |  | Pulmonary embolism | Vaccinated | HR 1.75 (1.53–2.00) |
|  |  |  | Unvaccinated | HR 4.15 (3.01–5.74) |
|  |  | Deep vein thrombosis | Vaccinated | HR 1.48 (1.30–1.67) |
|  |  |  | Unvaccinated | HR 1.61 (1.15–2.26) |
| Kim 2024 | NR | Pulmonary embolism | Vaccinated (≥2 doses) | HR 1.48 (1.15–1.89) |
|  |  |  | Booster dose (≥3 doses) | HR 1.31 (0.97–1.75) |
|  |  |  | Primary course (2 doses) | HR 2.01 (1.27–3.17) |
|  |  |  | Unvaccinated | HR 6.25 (3.67–10.65) |
|  |  | Deep vein thrombosis | Vaccinated (≥2 doses) | HR 1.15 (0.94–1.40) |
|  |  |  | Booster dose (≥3 doses) | HR 1.16 (0.93–1.44) |
|  |  |  | Primary course (2 doses) | HR 1.11 (0.69–1.76) |
|  |  |  | Unvaccinated | HR 3.05 (1.75–5.31) |
| Lim 2024 | Median 30 days | Composite CVD/CeVD/VTE (“Any cardiovascular, cerebrovascular, and other thrombotic complication”) | Booster dose (3 doses) | HR 1.10 (0.92–1.32) |
|  |  |  | Primary course (2 doses) | HR 1.11 (1.02–1.22) |
|  |  |  | Unvaccinated (0 doses) | HR 1.56 (1.29–1.90) |
|  |  | Composite CV/CeVD (“Major adverse cardiovascular/cerebrovascular events”) | Booster dose (3 doses) | HR 1.05 (0.81–1.35) |
|  |  |  | Primary course (2 doses) | HR 1.12 (0.99–1.26) |
|  |  |  | Unvaccinated (0 doses) | HR 1.51 (1.19–1.92) |
|  |  | Composite CeVD (“Cerebrovascular complications”) | Booster dose (3 doses) | HR 1.20 (0.89–1.62) |
|  |  |  | Primary course (2 doses) | HR 1.04 (0.88–1.22) |
|  |  |  | Unvaccinated (0 doses) | HR 1.25 (0.89–1.76) |
|  |  | All dysrhythmias | Booster dose (3 doses) | HR 1.38 (1.00–1.91) |
|  |  |  | Primary course (2 doses) | HR 1.18 (1.00–1.40) |
|  |  |  | Unvaccinated (0 doses) | HR 2.04 (1.53–2.72) |
|  |  | All ischaemic heart diseases | Booster dose (3 doses) | HR 0.98 (0.75–1.29) |
|  |  |  | Primary course (2 doses) | HR 1.00 (0.87–1.16) |
|  |  |  | Unvaccinated (0 doses) | HR 1.45 (1.07–1.95) |
|  |  | All other cardiac disorders | Booster dose (3 doses) | HR 0.93 (0.59–1.46) |
|  |  |  | Primary course (2 doses) | HR 1.43 (1.19–1.70) |
|  |  |  | Unvaccinated (0 doses) | HR 1.92 (1.39–2.65) |
|  |  | Stroke | Booster dose (3 doses) | HR 1.13 (0.80–1.59) |
|  |  |  | Primary course (2 doses) | HR 0.98 (0.82–1.17) |
|  |  |  | Unvaccinated (0 doses) | HR 1.27 (0.89–1.82) |
|  |  | Transient ischaemic attack | Booster dose (3 doses) | HR 1.25 (0.72–2.19) |
|  |  |  | Primary course (2 doses) | HR 1.20 (0.83–1.73) |
|  |  |  | Unvaccinated (0 doses) | HR 0.74 (0.23–2.38) |
|  |  | Atrial fibrillation | Booster dose (3 doses) | HR 1.07 (0.64–1.78) |
|  |  |  | Primary course (2 doses) | HR 1.14 (0.90–1.44) |
|  |  |  | Unvaccinated (0 doses) | HR 1.65 (1.11–2.46) |
|  |  | Sinus tachycardia | Booster dose (3 doses) | HR 0.95 (0.40–2.26) |
|  |  |  | Primary course (2 doses) | HR 0.82 (0.56–1.19) |
|  |  |  | Unvaccinated (0 doses) | HR 2.59 (1.39–4.81) |
|  |  | Sinus bradycardia | Booster dose (3 doses) | HR 1.39 (0.55–3.50) |
|  |  |  | Primary course (2 doses) | HR 1.40 (0.87–2.25) |
|  |  |  | Unvaccinated (0 doses) | HR 4.58 (1.49–14.07) |
|  |  | Other arrhythmia | Booster dose (3 doses) | HR 2.20 (1.40–3.46) |
|  |  |  | Primary course (2 doses) | HR 1.40 (1.05–1.86) |
|  |  |  | Unvaccinated (0 doses) | HR 2.15 (1.29–3.60) |
|  |  | Myocardial infarction | Booster dose (3 doses) | HR 1.25 (0.88–1.77) |
|  |  |  | Primary course (2 doses) | HR 0.66 (0.35–1.24) |
|  |  |  | Unvaccinated (0 doses) | HR 1.42 (0.99–2.03) |
|  |  | Acute coronary disease | Booster dose (3 doses) | HR 0.86 (0.63–1.19) |
|  |  |  | Primary course (2 doses) | HR 0.97 (0.82–1.15) |
|  |  |  | Unvaccinated (0 doses) | HR 1.11 (0.69–1.77) |
|  |  | Ischaemic cardiomyopathy | Booster dose (3 doses) | HR 1.65 (0.63–4.32) |
|  |  |  | Primary course (2 doses) | HR 0.81 (0.39–1.66) |
|  |  |  | Unvaccinated (0 doses) | HR 1.34 (0.52–3.46) |
|  |  | Angina | Booster dose (3 doses) | HR 0.82 (0.44–1.51) |
|  |  |  | Primary course (2 doses) | HR 1.32 (0.97–1.79) |
|  |  |  | Unvaccinated (0 doses) | HR 2.17 (1.09–4.32) |
|  |  | Heart failure | Booster dose (3 doses) | HR 0.70 (0.39–1.26) |
|  |  |  | Primary course (2 doses) | HR 1.45 (1.19–1.77) |
|  |  |  | Unvaccinated (0 doses) | HR 2.04 (1.44–2.90) |
|  |  | Non-ischaemic cardiomyopathy | Booster dose (3 doses) | HR 1.79 (0.82–3.93) |
|  |  |  | Primary course (2 doses) | HR 1.48 (0.98–2.23) |
|  |  |  | Unvaccinated (0 doses) | HR 1.67 (0.72–3.83) |
|  |  | Cardiac arrest | Booster dose (3 doses) | HR 1.58 (0.45–5.63) |
|  |  |  | Primary course (2 doses) | HR 0.75 (0.10–5.95) |
|  |  |  | Unvaccinated (0 doses) | HR 1.01 (0.29–3.52) |
|  |  | Cardiogenic shock | Primary course (2 doses) | HR 1.01 (0.45–2.27) |
|  |  |  | Unvaccinated (0 doses) | HR 0.61 (0.22–1.68) |
|  |  | Pulmonary embolism | Booster dose (3 doses) | HR 1.91 (0.67–5.47) |
|  |  |  | Primary course (2 doses) | HR 0.83 (0.45–1.52) |
|  |  |  | Unvaccinated (0 doses) | HR 1.52 (0.46–5.06) |
|  |  | Deep venous thrombosis | Booster dose (3 doses) | HR 0.47 (0.14–1.60) |
|  |  |  | Primary course (2 doses) | HR 1.27 (0.87–1.86) |
|  |  |  | Unvaccinated (0 doses) | HR 1.85 (0.95–3.60) |
|  |  | Deep venous thrombosis | Primary course (2 doses) | HR 2.57 (1.13–5.87) |
|  |  |  | Unvaccinated (0 doses) | HR 1.47 (0.21–10.35) |
|  |  | Arterial thromboses | Booster dose (3 doses) | HR 0.93 (0.51–1.69) |
|  |  |  | Primary course (2 doses) | HR 0.20 (0.03–1.53) |
|  |  |  | Unvaccinated (0 doses) | HR 1.30 (0.45–3.75) |
| Tran 2024 | 28 days | Composite CVD/CeVD/VTE (“Thromboembolic event”: venous thromboembolism, arterial thrombosis, cerebral venous sinus thrombosis, ischaemic stroke, and myocardial infarction) | Vaccinated (≥1 dose) | OR 2.786 (2.396–3.239) |
|  |  |  | Unvaccinated (0 doses) | OR 4.650 (4.183–5.170) |

CeVD, cerebrovascular disease; CI, confidence intervals; CVD, cardiovascular disease; HR, hazard ratio; NR, not reported; OR, odds ratio; VTE, venous thromboembolism.

#### Table 15 Outcomes included in meta-analyses

| **Study** | **Outcome description** | **Outcome identification details** |
| --- | --- | --- |
| **Composite CVD and CeVD** | | |
| Kim 2022 | Composite cardiovascular events (acute myocardial infarction, ischaemic stroke) | ICD-10 I21 (acute myocardial infarction), I63 (ischaemic stroke) |
| Jiang 2023 | Major adverse cardiovascular events | OMOP codeset ID 516422864). |
| Wan 2023 | Cardiovascular disease (coronary heart disease, stroke, heart failure) | ICD-9 419–414, 36.0, 36.1, V45.81 (coronary heart disease), 430-438 (stroke), 428, 398.91, 402.01, 402.11, 402.91, 404.01–404.03, 404.11–404.13, 404.91–404.93 (heart failure) |
| Song 2024 | Cardiovascular disease (coronary heart disease, stroke) | ICD-10 I20–I25 (coronary heart disease), ICD-10 I60–I69 (stroke) |
| **Stroke** | | |
| Al-Aly 2022 | Stroke^1^ | CCSR v 2021.1 referenced (code not reported, but likely for ‘cerebral infarction’) |
| Kim 2022 | Ischaemic stroke | ICD-10 I63 |
| Taquet 2022 | Ischaemic stroke | ICD-10 I63, H34.1 |
| Wan 2023 | Stroke | ICD-9 430-438 |
| Chen 2024 | Cerebral infarction | ICD-10 I63 |
| Song 2024 | Stroke | ICD-10 I60–I69 |
| Huh 2024 | Ischaemic stroke | ICD-10 I63 |
| **Coronary syndrome** | | |
| Al-Aly 2022 | Myocardial infarction | CCSR v 2021.1 referenced (code not reported) |
| Kim 2022 | Acute myocardial infarction | ICD-10 I21 |
| Taquet 2022 | Coronary disease | ICD-10 I21 (acute myocardial infarction), I22 (subsequent ST elevation and non-ST elevation myocardial infarction), I20.0 (unstable angina), I24 (other acute ischaemic heart disease), R57.0 (cardiogenic shock) |
| Wan 2023 | Coronary artery disease | ICD-9 410-414 (acute myocardial infarction, other acute and subacute form of ischaemic heart disease, old myocardial infarction, angina pectoris), 36.0 (removal of coronary artery obstruction and insertion of stent(s), 36.1 (bypass anastomosis for heart revascularization) |
| Song 2024 | Coronary heart diseases | ICD-10 I20 (angina pectoris), I21 (acute myocardial infarction), I22 (subsequent ST elevation and non-ST elevation myocardial infarction), I23 (certain current complications following acute myocardial infarction), I24 (other acute ischaemic heart disease), I25 (chronic ischaemic heart disease) |
| Huh 2024 | Ischaemic heart diseases | ICD-10 I21 (acute myocardial infarction), I22 (subsequent ST elevation and non-ST elevation myocardial infarction), I23 (certain current complications following acute myocardial infarction) |
| **Heart failure** | | |
| Al-Aly 2022 | Heart failure | CCSR v 2021.1 referenced (code not reported) |
| Taquet 2022 | Cardiac failure | ICD-10 I50, I11.0, I13.0 |
| Wan 2023 | Heart failure | ICD-9 428.x, 398.91, 402.01, 402.11,  402.91, 404.01–404.03, 404.11–404.13, 404.91–404.93 |
| Huh 2024 | Heart failure and cardiomyopathies | ICD-10 I40, I43, I50 |
| **Arrhythmia** | | |
| Al-Aly 2022 | Atrial fibrillation | CCSR v 2021.1 referenced (code not reported) |
| Taquet 2022 | Arrhythmia | ICD-10 I47, I48, I49, R00.0 |
| Huh 2024 | Cardiac dysrhythmias | ICD-10 I44, I45, I47, I48, I49 |
| **Venous thromboembolism** | | |
| Al-Aly 2022 | Deep vein thrombosis | CCSR v 2021.1 referenced (code not reported) |
| Taquet 2022 | Hypercoagulopathy, deep vein thrombosis, pulmonary embolism | ID-10 D68, I82, I26 |
| Xie 2022 | Venous thromboembolism | ICD-10 I26, I80, I81, I82 |
| Kim 2024 | Deep vein thrombosis | ICD-10 I801–8033 |
| Huh 2024 | Venous thromboembolism | ICD-10 I63.6, I67.6, I81, I82.2, I82.3, I82.8, I82.9 |

^1^ Assumed ‘ischaemic stroke’ by reviewers.

CCSR, Clinical Classifications Software Refined; CeVD, cerebrovascular disease; CVD, cardiovascular disease; ICD, International Classification of Diseases; OMOP, Observational Medical Outcomes Partnership.

#### Table 16 Meta-analysis leave-one-out sensitivity analyses

| **Leave-one-out analysis** | **Pooled HR (95% CI)** | ***P* value** |
| --- | --- | --- |
| **Composite CVD and CeVD** | | |
| **Main analysis** | **0.60 (0.51–0.69)** | **<0.0001** |
| Omitting Kim 2022 | 0.62 (0.54–0.72) | <0.0001 |
| Omitting Jiang 2023 | 0.57 (0.43–0.76) | <0.0001 |
| Omitting Wan 2023 | 0.60 (0.51–0.72) | <0.0001 |
| Omitting Song 2024 | 0.56 (0.48–0.65) | <0.0001 |
| **Stroke** | | |
| **Main analysis** | **0.75 (0.64–0.88)** | **0.0003** |
| Omitting Al-Aly 2022 | 0.74 (0.61–0.88) | 0.0011 |
| Omitting Kim 2022 | 0.80 (0.71–0.90) | 0.0003 |
| Omitting Taquet 2022 | 0.72 (0.60–0.85) | 0.0001 |
| Omitting Wan 2023 | 0.77 (0.65–0.91) | 0.0020 |
| Omitting Chen 2024 | 0.76 (0.64–0.90) | 0.0014 |
| Omitting Song 2024 | 0.73 (0.58–0.91) | 0.0047 |
| Omitting Huh 2024 | 0.71 (0.59–0.85) | 0.0002 |
| **Coronary syndrome** | | |
| **Main analysis** | **0.70 (0.52–0.95)** | **0.0199** |
| Omitting Al-Aly 2022 | 0.70 (0.50–0.98) | 0.0390 |
| Omitting Kim 2022 | 0.74 (0.53–1.01) | 0.0606 |
| Omitting Taquet 2022 | 0.60 (0.54–0.67) | <0.0001 |
| Omitting Wan 2023 | 0.72 (0.52–1.00) | 0.0526 |
| Omitting Song 2024 | 0.75 (0.55–1.03) | 0.0777 |
| Omitting Huh 2024 | 0.69 (0.47–1.02) | 0.0616 |
| **Heart failure** | | |
| **Main analysis** | **0.72 (0.47–1.10)** | **0.1239** |
| Omitting Al-Aly 2022 | 0.61 (0.34–1.10) | 0.1034 |
| Omitting Taquet 2022 | 0.60 (0.36–1.02) | 0.0605 |
| Omitting Wan 2023 | 0.83 (0.53–1.29) | 0.4031 |
| Omitting Huh 2024 | 0.89 (0.65–1.22) | 0.4749 |
| **Arrhythmia** | | |
| **Main analysis** | **0.82 (0.69–0.98)** | **0.0313** |
| Omitting Al-Aly 2022 | 0.83 (0.64–1.09) | 0.1892 |
| Omitting Taquet 2022 | 0.75 (0.67–0.85) | <0.0001 |
| Omitting Huh 2024 | 0.88 (0.73–1.05) | 0.1533 |
| **Venous thromboembolism** | | |
| **Main analysis** | **0.48 (0.30–0.75)** | **0.0014** |
| Omitting Al-Aly 2022 | 0.44 (0.25–0.80) | 0.0069 |
| Omitting Taquet 2022 | 0.40 (0.27–0.60) | <0.0001 |
| Omitting Xie 2022 | 0.59 (0.39–0.88) | 0.0105 |
| Omitting Kim 2024 | 0.52 (0.32–0.85) | 0.0086 |
| Omitting Huh 2024 | 0.45 (0.24–0.86) | 0.0151 |

CeVD, cerebrovascular disease; CI, confidence intervals; CV, cardiovascular disease; HR, hazard ratio.

#### Table 17 Meta-analysis sensitivity analyses

| **Sensitivity analysis** | **Pooled HR (95% CI)** | ***P* value** | **Q** | **Degrees of freedom** | **Q *P* value** | ***I*^2^** |
| --- | --- | --- | --- | --- | --- | --- |
| **Composite CVD and CeVD** | | | | | | |
| **Main analysis** | **0.60 (0.51–0.69)** | **<0.0001** | **14.49** | **3** | **0.0023** | **79.3%** |
| Lowest dose | 0.69 (0.50–0.95) | 0.0228 | 22.95 | 3 | <0.0001 | 86.9% |
| Highest dose | 0.51 (0.44–0.59) | <0.0001 | 14.14 | 3 | 0.0027 | 78.8% |
| Longest follow-up | 0.62 (0.55–0.70) | <0.0001 | 13.77 | 3 | 0.0032 | 78.2% |
| Add CVD- and CeVD-only composite outcomes (Al-Aly 2022, Chen 2024) | 0.64 (0.54–0.75) | <0.0001 | 46.73 | 5 | <0.0001 | 89.3% |
| **Stroke** | | | | | | |
| **Main analysis** | **0.75 (0.64–0.88)** | **0.0003** | **17.94** | **6** | **0.0064** | **66.6%** |
| Lowest dose | 0.79 (0.61–1.04) | 0.0887 | 39.85 | 6 | <0.0001 | 84.9% |
| Highest dose | 0.67 (0.53–0.83) | 0.0004 | 36.17 | 6 | <0.0001 | 83.4% |
| Longest follow-up | 0.75 (0.65–0.87) | 0.0001 | 19.30 | 6 | 0.0037 | 68.9% |
| Remove potential population overlap (Taquet 2022) | 0.72 (0.60–0.85) | 0.0001 | 15.46 | 5 | 0.0086 | 67.7% |
| **Coronary syndrome** | | | | | | |
| **Main analysis** | **0.70 (0.52–0.95)** | **0.0199** | **28.59** | **5** | **<0.0001** | **82.5%** |
| Lowest dose | 0.80 (0.67–0.97) | 0.0196 | 7.39 | 5 | 0.1934 | 32.3% |
| Highest dose | 0.61 (0.33–1.14) | 0.1241 | 128.91 | 5 | <0.0001 | 96.1% |
| Longest follow-up | 0.74 (0.57–0.96) | 0.0236 | 30.70 | 5 | <0.0001 | 83.7% |
| Replace with acute coronary disease outcome (Al-Aly 2022) | 0.74 (0.56–0.98) | 0.0383 | 35.65 | 5 | <0.0001 | 86.0% |
| Remove only myocardial infarction outcomes (Al-Aly 2022, Kim 2022) | 0.74 (0.51–1.07) | 0.1141 | 27.54 | 3 | <0.0001 | 89.1% |
| **Heart failure** | | | | | | |
| **Main analysis** | **0.72 (0.47–1.10)** | **0.1239** | **54.04** | **3** | **<0.0001** | **94.4%** |
| Lowest dose | 0.81 (0.53–1.22) | 0.3062 | 38.53 | 3 | <0.0001 | 92.2% |
| Highest dose | 0.70 (0.45–1.09) | 0.1152 | 55.38 | 3 | <0.0001 | 94.6% |
| Longest follow-up | 0.70 (0.46–1.06) | 0.0958 | 67.10 | 3 | <0.0001 | 95.5% |
| Remove outcomes pooled with cardiomyopathies (Huh 2024) | 0.89 (0.65–1.22) | 0.4749 | 9.01 | 2 | 0.0110 | 77.8% |
| **Arrhythmia** | | | | | | |
| **Main analysis** | **0.82 (0.69–0.98)** | **0.0313** | **9.19** | **2** | **0.0101** | **78.2%** |
| Lowest dose | 0.83 (0.68–1.03) | 0.0870 | 10.20 | 2 | 0.0061 | 80.4% |
| Replace with other dysrhythmias outcome (Al-Aly 2022) | 0.74 (0.55–0.99) | 0.0430 | 15.45 | 2 | 0.0004 | 87.1% |
| **Venous thromboembolism** | | | | | | |
| **Main analysis** | **0.48 (0.30–0.75)** | **0.0014** | **48.94** | **4** | **<0.0001** | **91.8%** |
| Lowest dose | 0.48 (0.31–0.73) | 0.0006 | 37.32 | 4 | <0.0001 | 89.3% |
| Remove potential population overlap (Kim 2024) | 0.52 (0.32–0.85) | 0.0086 | 41.18 | 3 | <0.0001 | 92.7% |
| Replace with pulmonary embolism outcome (Al-Aly 2022, Huh 2024, Kim 2024) | 0.36 (0.18–0.70) | 0.0029 | 109.21 | 4 | <0.0001 | 96.3% |

CeVD, cerebrovascular disease; CI, confidence intervals; CV, cardiovascular disease; HR, hazard ratio.

#### Table 18 GRADE certainty of evidence assessment

| **Meta-analysis** | **N studies** | **Study design** | **Risk of bias** | **Inconsistency** | **Indirectness** | **Imprecision** | **Dissemination bias** | **Other considerations** | **Pooled risk (95% CI)** | ***P* value** | **Certainty of Evidence (GRADE)** |
| --- | --- | --- | --- | --- | --- | --- | --- | --- | --- | --- | --- |
| Composite CV/CeVD | 4 | Observational | None | None | None | None | None | None | HR 0.60 (0.51–0.69) | <0.0001 | ꚚꚚOO  Low |
| Stroke | 7 | Observational | None | None | None | None | None | None | HR 0.75 (0.64–0.88) | 0.0003 | ꚚꚚOO  Low |
| Coronary syndrome | 6 | Observational | None | Serious | None | Serious | None | None | HR 0.70 (0.52–0.95) | 0.0199 | ꚚOOO  Very low |
| Heart failure | 4 | Observational | None | Serious | None | Very serious | None | None | HR 0.72 (0.47–1.10) | 0.1239 | ꚚOOO  Very low |
| Arrhythmia | 3 | Observational | None | None | None | Serious | None | None | HR 0.82 (0.69–0.98) | 0.0313 | ꚚOOO  Very low |
| Venous thromboembolism | 5 | Observational | None | None | None | None | None | None | HR 0.51 (0.36–0.73) | 0.0014 | ꚚꚚOO  Low |

Low certainty implies that the available evidence is limited and the true effect may be substantially different from the estimate.
Very low certainty indicates that the available evidence is insufficient to support any firm conclusions.

CeVD, cerebrovascular disease; CI, confidence intervals; CV, cardiovascular disease; GRADE, Grading of Recommendations Assessment Development, and Evaluation; HR, hazard ratio.

### Search strategies

#### EMBASE strategy

Embase (Ovid): 2021-2025/09/08

Searched 9.9.25

<https://ovidsp.ovid.com/ovidweb.cgi?T=JS&NEWS=N&PAGE=main&SHAREDSEARCHID=2ghL36CiCUE7QZZByJRK6dq7ldKBtFNCPPs9SL3lY97zyDctZiLaS7seIrRuVxinV>

1	exp coronavirus disease 2019/	482231

2	exp SARS coronavirus/	10972

3	exp Severe acute respiratory syndrome coronavirus 2/	142868

4	(coronavirinae/ or betacoronavirus/ or coronavirus infection/) and (epidemic/ or pandemic/)	11089

5	(Coronavirus$ or "covid 19" or "2019-ncov").ti,ab,bk,kw,kf,ot.	530141

6	("2019-ncov" or "2019ncov" or corona-virus$ or "cov19" or "cov-19" or "19nCoV" or "COVID19" or "COVID2019" or "Covid 2019").ti,ab,bk,kw,kf,ot.	21113

7	(ncov$ or "sars cov$" or sarscov$ or "sars coronavirus$" or coronovirus$ or corono$ virus$ or "19-nCoV$" or 19nCoV$).ti,ab,bk,kw,kf,ot.	197607

8	("SARS2$" or "SARS-2$" or SARScoronavirus$ or SARS-coronavirus$ or SARScoronovirus$ or SARS-coronovirus$).ti,ab,bk,kw,kf,ot.	3221

9	("HCoV-19$" or "HCoV19$" or "HCoV-2019$" or "HCoV2019$").ti,ab,bk,kw,kf,ot.	87

10	("2019 novel$" or Ncov$).ti,ab,bk,kw,kf,ot.	6563

11	("Severe Acute Respiratory Syndrome Coronavirus 2" or "Severe Acute Respiratory Syndrome Corona Virus 2").ti,ab,bk,kw,kf,ot.	43509

12	or/1-11	621348

13	Clinical study/	170805

14	Case control study/	241566

15	Family study/	26565

16	Longitudinal study/	254318

17	Retrospective study/	1892639

18	Prospective study/	1005104

19	Randomized controlled trials/	299597

20	18 not 19	992864

21	Cohort analysis/	1440401

22	(Cohort adj (study or studies)).mp.	611508

23	(Case control adj (study or studies)).tw.	191834

24	(follow up adj (study or studies)).tw.	83234

25	(observational adj (study or studies)).tw.	339443

26	(epidemiologic$ adj (study or studies)).tw.	133490

27	(cross sectional adj (study or studies)).tw.	439379

28	or/13-17,20-27	4868740

29	("real world" or RWE or RWD or "Real-life").ti,ab,ot,bk,kf,kw.	261245

30	(registry or register$ or survey).ti,ab.	1706333

31	(patient record$ or medical record$).ti,ab.	329679

32	(electronic health record$ or EHR).ti,ab,bk,kf,kw.	68516

33	(electronic medical record$ or EMR).ti,ab,bk,kf,kw.	88933

34	(dataset$ or data set$).ti,ab,bk,kf,kw.	545488

35	(administrat$ adj3 claim$).ti,ab,bk,kf,kw.	9929

36	(pragmatic trial$ or prct).ti,ab,ot,bk,kf,kw.	4165

37	or/29-36	2772987

38	28 or 37	6862668

39	animal/ or animal experiment/ or (rat or rats or mouse or mice or murine or rodent or rodents or hamster or hamsters or pig or pigs or porcine or rabbit or rabbits or animal or animals or dogs or dog or cat or cats or cows or cow or bovine or sheep or ovine or monkey or monkeys).ti,ab,ot,hw.	8348268

40	exp human/ or human experiment/	29003205

41	39 not (39 and 40)	6123936

42	38 not 41	6713755

43	42 not (letter or editorial or note).pt.	6605101

44	limit 43 to yr="2021 -Current"	2442557

45	tozinameran/	16522

46	((pfizer$ or biontech) adj2 (vaccin$ or jab or jabs or shot or shots or immunis$ or immuniz$)).ti,ab,ot,bk,kf,kw.	3266

47	(tozinameran or Comirnaty or "Pfizer-BioNTech" or "pf07302048" or "pf-07302048" or " BNT-162b2" or "BNT162b2" or "rbp 020.2" or " rbp020.2").ti,ab,ot,bk,kf,kw,hw,du,dy,tn,rn.	18649

48	(Pidacmeran or "BNT162C2" or "BNT-162C2" or Abdavomeran or "BNT-162B1" or "BNT162B1" or "BNT162A1" or "BNT-162A1").ti,ab,ot,bk,kf,kw,hw,du,dy,tn,rn.	157

49	"2417899-77-3".af.	15371

50	(pfizer or biontech or "Pfizer-BioNTech").mf.	60356

51	or/45-50	71847

52	elasomeran/	9380

53	davesomeran plus elasomeran/	61

54	elasomeran plus imelasomeran/	88

55	(Elasomeran or moderna or Spikevax or davesomeran or imelasomeran or andusomeran or "mRNA 1273" or "mRNA1273" or "mRNA-1273.211" or "mRNA 1273.211" or Spikevax or "M-1273" or "M1273" or "CX-024414" or "CX024414" or "TAK-919" or "TAK919" or "mRNA1273" or "mRNA-1273" or "messenger RNA1273" or "messenger RNA 1273").ti,ab,ot,bk,kf,kw,hw,du,dy,tn,rn.	12480

56	("2430046-03-8" or "2457298-05-2").af.	8343

57	moderna.mf.	6093

58	or/52-57	13802

59	Ibacovavec/	3606

60	(Jcovden or ibacovavec or "Ad26.COV2.S" or "JNJ-78436735" or "JNJ78436735" or "Ad26COVS1" or "VAC31518" or "VAC-31518").ti,ab,ot,bk,kf,kw,hw,du,dy,tn,rn.	3653

61	((Janssen or "J&J" or "Johnson & Johnson") adj2 (vaccin$ or jab or jabs or shot or shots or immunis$ or immuniz$)).ti,ab,ot,bk,kf,kw.	499

62	"2541607-46-7".af.	1692

63	or/59-62	3922

64	Vaxzevria/	8491

65	(Vaxzevria or Covishield or "AZD1222" or "AZD-1222" or "AZD2816" or "AZD-2816" or "ChAdOx1" or "ChAdOx-1" or "ChAdOx1-s" or "ChAdOx1s" or "ChAdOx-1-s" or "ChAdOx-1s").ti,ab,ot,bk,kf,kw,hw,du,dy,tn,rn.	9141

66	(("Oxford-AstraZeneca" or AstraZeneca or AZ) adj2 (vaccin$ or jab or jabs or shot or shots or immunis$ or immuniz$)).ti,ab,ot,bk,kf,kw.	1147

67	"2420395-83-9".af.	7669

68	or/64-67	9576

69	Nvx-cov2373 vaccine/	1131

70	(Novavax or CovoVax or Nuvaxovid or "NVXCoV2373" or "NVX-CoV2373" or "nvx cov 2373" or "nvx cov2373" or "NVXCoV-2373" or " NVX-CoV2601" or " NVXCoV2601" or " NVX-CoV-2601" or " NVXCoV-2601" or "sars-cov-2 rs" or "sarscov-2 rs" or "sarscov2 rs" or "sars-cov-2rs" or "sarscov2rs" or "sarscov-2rs" or "tak 019" or "tak019").ti,ab,ot,bk,kf,kw,hw,du,dy,tn,rn.	1440

71	"2502099-58-1".af.	0

72	or/69-71	1440

73	valneva COVID-19 vaccine/	77

74	("VLA2001" or "VLA2101" or "VLA-2001" or "VLA-2101" or Valneva).ti,ab,ot,bk,kf,kw,hw,du,dy,tn,rn.	162

75	"2695500-31-1".af.	0

76	or/73-75	162

77	vidprevtyn beta/	23

78	("vidprevtyn beta" or "vidprevtynbeta" or "vidprevtynb" or "vidprevtyn-b" or "VAT0000" or "VAT00008" or "VAT-0000" or "VAT-00008").ti,ab,ot,bk,kf,kw,hw,du,dy,tn,rn.	26

79	(("sp/gsk" or "subunit B.1.351" or "Sanofi GSK") adj2 (vaccin$ or jab or jabs or shot or shots or immunis$ or immuniz$)).ti,ab,ot,bk,kf,kw.	4

80	"2696235-99-9".af.	0

81	or/77-80	26

82	or/51,58,63,68,72,76,81	79809

83	exp cardiovascular disease/	5878734

84	exp cerebrovascular disease/	1034068

85	(CVD or CBV or CHD or CBA or CBAs or TIA or TIAs or IHD or MI or CVA or CVAs or AF).ti,ab,ot,bk,kf,kw.	459271

86	(("Peripheral vascular" or Cerebrovascular or cardiovascular or cardio-vascular or Cerebro-vascular) adj2 (accident$ or failure$ or attack$ or clot or clots or embolus or emboli or disease$ or comorbidit$ or co-morbidit$)).ti,ab,ot,bk,kf,kw.	502360

87	((Congenital or coronary or ischaemic or ischemic or heart or congestive or cardiac or Atherosclerotic or cardiovascular or cardio-vascular) adj3 (failure$ or disease$ or attack$ or embolus or emboli or clot or clots)).ti,ab,ot,bk,kf,kw.	1374971

88	(Atrial fibrillat$ or Thrombosis or thrombotic or thromboses or Stroke or strokes or "blood clot" or "blood clots" or Hypertension or Myocardial infarction$ or Transient Ischemic Attack$ or "pulmonary embolus" or "pulmonary emboli" or Congenital Heart Problem$).ti,ab,ot,bk,kf,kw.	2036923

89	((Circulatory or Cardiac or vascular) adj2 (injur$ or damag$ or patholog$ or failure$ or comorbidit$ or co-morbidit$ or complication$ or disorder$ or syndrome$ or disease$ or lesion$ or disturbance$ or syndrome$ or event$)).ti,ab,ot,bk,kf,kw.	413901

90	((cardiovascular or cardio-vascular or angiocardiovascular or angio-cardiovascular or angio-cardio-vascular or cerebrovascular or cerebro-vascular or "cerebral vascular" or heart or cardiac or myocardial or cerebral) adj3 (complication$ or disorder$ or syndrome$ or comorbidit$ or co-morbidit$ or disease$ or lesion$ or disturbance$ or event$ or syndrome$ or damag$ or injur$ or failure$ or patholog$)).ti,ab,ot,bk,kf,kw.	1533653

91	(cerebrovasculopath$ or cerebro-vasculopath$ or cerebroangiopath$ or cerebro-angiopath$ or cerebral vasculopath$ or cerebral angiopath$ or brain vasculopath$ or brain angiopath$ or Angiocardiopath$ or Angio-cardiopath$).ti,ab,ot,bk,kf,kw.	1296

92	co-morbid$.ti,ab,ot,bk,kf,kw.	89695

93	comorbidity/	485703

94	comorbid$.ti,ab,ot,bk,kf,kw.	499933

95	(multimorbid$ or multi-morbid$ or polymorbid$ or poly-morbid$).ti,ab,ot,bk,kf,kw.	19451

96	(pluralpathol$ or plural-pathol$ or pluralmorbid$ or plural-morbid$ or "dual diagnosis" or "dual diagnoses" or "plural diagnosis" or "plural diagnoses").ti,ab,ot,bk,kf,kw.	4193

97	exp multiple chronic conditions/	13277

98	((multifactorial or multi-factorial or multiple or concurrent or simultaneous) adj3 (condition$ or disease$ or syndrome$ or disorder$ or illness$)).ti,ab,ot,bk,kf,kw.	149002

99	((CVD or CVB or CVA or CV or Cerebrovascular or cardiovascular or cardio-vascular or Cerebro-vascular) adj5 (hospital$ or mortalit$ or death$ or admission$)).ti,ab,ot,bk,kf,kw.	156580

100	or/83-99	6973647

101	12 and 82 and 44 and 100	3224

*COVID facet based on terms from:*

 World Health Organization (26 May 2021) WHO COVID-19 Database Search Strategy. Systematic search of the COVID-19 literature performed Monday through Friday for the WHO Database. Search strategy as of 26 May 2021. Searches performed by Tomas Allen, Kavita Kothari, and Martha Knuth. Available from: <https://www.who.int/docs/default-source/coronaviruse/who-covid-19-database/who-covid-19_sources_searchstrategy_20210526.pdf?sfvrsn=65209cc2_5>

Canadian Agency for Drugs and Technologies in Health (2.9.21) CADTH COVID-19 Search Strings: COVID-19 — EMBASE (Internet). Available from: <https://covid.cadth.ca/literature-searching-tools/cadth-covid-19-search-strings/>

NICE (18 December 2020) [accessed 17.8.21] COVID-19 rapid guideline: managing the long-term effects of COVID-19 [NG188]. Search history record [PDF]. NICE: London. Available from: <https://www.nice.org.uk/guidance/ng188/evidence/search-strategies-pdf-8957634445>

*Observational study design filter adapted from:*

Scottish Intercollegiate Guidelines Network (SIGN). Search filters: observational studies. Embase. Edinburgh: SIGN, Last Modified 24/04/17 Available from: <https://www.sign.ac.uk/what-we-do/methodology/search-filters/>

Observational Studies - Embase. In: CADTH Search Filters Database. Ottawa: CADTH; 2023 [accessed 7.11.23]: <https://searchfilters.cadth.ca/link/37>

#### Medline strategy

Medline ALL (Ovid): 2021-2025/09/10

Searched 11.9.25

<https://ovidsp.ovid.com/ovidweb.cgi?T=JS&NEWS=N&PAGE=main&SHAREDSEARCHID=2gzlliVf9Xi7Zk2D4rYu1oQnEDMne9uE1lZWjO39kSFIFELmPN1OONPMxBuxB2cAo>

1	exp COVID-19/	300744

2	exp Severe acute respiratory syndrome-related coronavirus/	202375

3	coronaviridae/ or exp coronavirus/	215485

4	Betacoronavirus/	33581

5	Coronavirus Infections/	47062

6	or/3-5	222511

7	epidemics/ or pandemics/	155476

8	Disease Outbreaks/	96068

9	or/7-8	246624

10	6 and 9	93067

11	(Coronavirus$ or "covid 19" or 2019-ncov).ti,ab,kw,kf,ot.	458886

12	("2019-ncov" or "2019ncov" or corona-virus$ or "cov19" or "cov-19" or "19nCoV" or "COVID19" or "COVID2019" or "Covid 2019").ti,ab,kw,kf,ot.	11185

13	(ncov$ or "sars cov$" or sarscov$ or "sars coronavirus$" or coronovirus$ or corono$ virus$ or "19-nCoV$" or "19nCoV$").ti,ab,kw,kf,ot.	165152

14	("SARS2$" or "SARS-2$" or SARScoronavirus$ or SARS-coronavirus$ or SARScoronovirus$ or SARS-coronovirus$).ti,ab,kw,kf,ot.	2763

15	("HCoV-19$" or "HCoV19$" or "HCoV-2019$" or "HCoV2019$").ti,ab,kw,kf,ot.	70

16	("Severe Acute Respiratory Syndrome Coronavirus 2" or "Severe Acute Respiratory Syndrome Corona Virus 2").ti,ab,kw,kf,ot.	42061

17	("2019 novel$" or Ncov$).ti,ab,kw,kf,ot.	5543

18	or/1-2,10-17	497376

19	exp animals/ not humans.sh.	5374612

20	18 not 19	488975

21	limit 20 to yr="2021 -Current"	403591

22	21 not (comment or editorial or letter).pt.	369389

23	exp epidemiologic studies/	3529783

24	Epidemiologic Methods/	31649

25	Observational Studies as Topic/	11368

26	Single-Case Studies as Topic/	166

27	case reports as topic/	226

28	case reports.pt.	2502175

29	(Observational Study or Validation Studies or Clinical Study).pt.	187841

30	Case control.ti,ab,ot,kw,kf.	176504

31	(cohort adj (study or studies)).ti,ab,ot,kw,kf.	420208

32	Cohort analy$.ti,ab,ot,kw,kf.	16647

33	("Follow up" adj (study or studies)).ti,ab,ot,kw,kf.	63379

34	(observational adj (study or studies)).ti,ab,ot,kw,kf.	210377

35	Longitudinal.ti,ab,ot,kw,kf.	393837

36	Retrospective.ti,ab,ot,kw,kf.	936314

37	Cross sectional.ti,ab,ot,kw,kf.	652444

38	(epidemiologic$ adj (study or studies)).ti,ab,ot,kw,kf.	104152

39	or/23-38	6881251

40	("real world" or RWE or RWD or "Real-life").ti,ab,ot,bk,kf,kw.	157059

41	(registry or register$ or survey).ti,ab.	1248686

42	(patient record$ or medical record$).ti,ab.	182617

43	(electronic health record$ or EHR).ti,ab,bk,kf,kw.	43761

44	(electronic medical record$ or EMR).ti,ab,bk,kf,kw.	39605

45	(dataset$ or data set$).ti,ab,bk,kf,kw.	438348

46	(administrat$ adj3 claim$).ti,ab,bk,kf,kw.	5389

47	(pragmatic trial$ or prct).ti,ab,ot,bk,kf,kw.	2684

48	or/40-47	1988609

49	39 or 48	8185174

50	BNT162 Vaccine/	4289

51	((pfizer$ or biontech) adj2 (vaccin$ or jab or jabs or shot or shots or immunis$ or immuniz$)).ti,ab,ot,bt,kf,kw.	2097

52	(tozinameran or Comirnaty or "Pfizer-BioNTech" or "pf07302048" or "pf-07302048" or " BNT-162b2" or "BNT162b2" or "rbp 020.2" or " rbp020.2").ti,ab,ot,bt,kf,kw,hw,nm,rn.	7511

53	(Pidacmeran or "BNT162C2" or "BNT-162C2" or Abdavomeran or "BNT-162B1" or "BNT162B1" or "BNT162A1" or "BNT-162A1").ti,ab,ot,bt,kf,kw,hw,nm,rn.	22

54	"2417899-77-3".af.	0

55	or/50-54	8337

56	2019-nCoV Vaccine mRNA-1273/	1064

57	(Elasomeran or moderna or Spikevax or davesomeran or imelasomeran or andusomeran or "mRNA 1273" or "mRNA1273" or "mRNA-1273.211" or "mRNA 1273.211" or Spikevax or "M-1273" or "M1273" or "CX-024414" or "CX024414" or "TAK-919" or "TAK919" or "mRNA1273" or "mRNA-1273" or "messenger RNA1273" or "messenger RNA 1273").ti,ab,ot,bt,kf,kw,hw,nm,rn.	4788

58	("2430046-03-8" or "2457298-05-2").af.	0

59	or/56-58	4788

60	(Jcovden or ibacovavec or "Ad26.COV2.S" or "JNJ-78436735" or "JNJ78436735" or "Ad26COVS1" or "VAC31518" or "VAC-31518").ti,ab,ot,bt,kf,kw,hw,nm,rn.	716

61	"2541607-46-7".af.	0

62	((Janssen or "J&J" or "Johnson & Johnson") adj2 (vaccin$ or jab or jabs or shot or shots or immunis$ or immuniz$)).ti,ab,ot,bt,kf,kw.	312

63	or/60-62	920

64	ChAdOx1 nCoV-19/	1350

65	(Vaxzevria or Covishield or "AZD1222" or "AZD-1222" or "AZD2816" or "AZD-2816" or "ChAdOx1" or "ChAdOx-1" or "ChAdOx1-s" or "ChAdOx1s" or "ChAdOx-1-s" or "ChAdOx-1s").ti,ab,ot,bt,kf,kw,hw,nm,rn.	3044

66	(("Oxford-AstraZeneca" or AstraZeneca or AZ) adj2 (vaccin$ or jab or jabs or shot or shots or immunis$ or immuniz$)).ti,ab,ot,bk,kf,kw.	832

67	"2420395-83-9".af.	0

68	or/64-67	3498

69	(Novavax or CovoVax or Nuvaxovid or "NVXCoV2373" or "NVX-CoV2373" or "nvx cov 2373" or "nvx cov2373" or "NVXCoV-2373" or " NVX-CoV2601" or " NVXCoV2601" or " NVX-CoV-2601" or " NVXCoV-2601" or "sars-cov-2 rs" or "sarscov-2 rs" or "sarscov2 rs" or "sars-cov-2rs" or "sarscov2rs" or "sarscov-2rs" or "tak 019" or "tak019").ti,ab,ot,bt,kf,kw,hw,nm,rn.	301

70	"2502099-58-1".af.	0

71	or/69-70	301

72	("VLA2001" or "VLA2101" or "VLA-2001" or "VLA-2101" or Valneva).ti,ab,ot,bt,kf,kw,hw,nm,rn.	32

73	"2695500-31-1".af.	0

74	or/72-73	32

75	("vidprevtyn beta" or "vidprevtynbeta" or "vidprevtynb" or "vidprevtyn-b" or "VAT0000" or "VAT00008" or "VAT-0000" or "VAT-00008").ti,ab,ot,bt,kf,kw,hw,nm,rn.	4

76	(("sp/gsk" or "subunit B.1.351" or "Sanofi GSK") adj2 (vaccin$ or jab or jabs or shot or shots or immunis$ or immuniz$)).ti,ab,ot,bk,kf,kw.	0

77	"2696235-99-9".af.	0

78	or/75-77	4

79	or/55,59,63,68,71,74,78	13081

80	exp Cardiovascular Diseases/	2891061

81	Cerebrovascular Disorders/	48696

82	(CVD or CBV or CHD or CBA or CBAs or TIA or TIAs or IHD or MI or CVA or CVAs or AF).ti,ab,ot,bt,kf,kw.	274516

83	(("Peripheral vascular" or Cerebrovascular or cardiovascular or cardio-vascular or Cerebro-vascular) adj2 (accident$ or failure$ or attack$ or clot or clots or embolus or emboli or disease$ or comorbidit$ or co-morbidit$)).ti,ab,ot,bt,kf,kw.	333734

84	((Congenital or coronary or ischaemic or ischemic or heart or congestive or cardiac or Atherosclerotic or cardiovascular or cardio-vascular) adj3 (failure$ or disease$ or attack$ or embolus or emboli or clot or clots)).ti,ab,ot,bt,kf,kw.	897250

85	(Atrial fibrillat$ or Thrombosis or thrombotic or thromboses or Stroke or strokes or "blood clot" or "blood clots" or Hypertension or Myocardial infarction$ or Transient Ischemic Attack$ or "pulmonary embolus" or "pulmonary emboli" or Congenital Heart Problem$).ti,ab,ot,bt,kf,kw.	1294321

86	((Circulatory or Cardiac or vascular) adj2 (injur$ or damag$ or patholog$ or failure$ or comorbidit$ or co-morbidit$ or complication$ or disorder$ or syndrome$ or disease$ or lesion$ or disturbance$ or syndrome$ or event$)).ti,ab,ot,bt,kf,kw.	273719

87	((cardiovascular or cardio-vascular or angiocardiovascular or angio-cardiovascular or angio-cardio-vascular or cerebrovascular or cerebro-vascular or "cerebral vascular" or heart or cardiac or myocardial or cerebral) adj3 (complication$ or disorder$ or syndrome$ or comorbidit$ or co-morbidit$ or disease$ or lesion$ or disturbance$ or event$ or syndrome$ or damag$ or injur$ or failure$ or patholog$)).ti,ab,ot,bt,kf,kw.	1009111

88	(cerebrovasculopath$ or cerebro-vasculopath$ or cerebroangiopath$ or cerebro-angiopath$ or cerebral vasculopath$ or cerebral angiopath$ or brain vasculopath$ or brain angiopath$ or Angiocardiopath$ or Angio-cardiopath$).ti,ab,ot,bt,kf,kw.	798

89	co-morbid$.ti,ab,ot,bt,kf,kw.	36751

90	exp Comorbidity/	135664

91	comorbid$.ti,ab,ot,bt,kf,kw.	284941

92	(multimorbid$ or multi-morbid$ or polymorbid$ or poly-morbid$).ti,ab,ot,bt,kf,kw.	14570

93	(pluralpathol$ or plural-pathol$ or pluralmorbid$ or plural-morbid$ or "dual diagnosis" or "dual diagnoses" or "plural diagnosis" or "plural diagnoses").ti,ab,ot,bt,kf,kw.	2470

94	Multiple Chronic Conditions/	846

95	((multifactorial or multi-factorial or multiple or concurrent or simultaneous) adj3 (condition$ or disease$ or syndrome$ or disorder$ or illness$)).ti,ab,ot,bt,kf,kw.	103077

96	((CVD or CVB or CVA or CV or Cerebrovascular or cardiovascular or cardio-vascular or Cerebro-vascular) adj5 (hospital$ or mortalit$ or death$ or admission$)).ti,ab,ot,bt,kf,kw.	93518

97	or/80-96	4138791

98	22 and 79 and 49 and 97	1279

*COVID facet based on terms from:*

 World Health Organization (26 May 2021) WHO COVID-19 Database Search Strategy. Systematic search of the COVID-19 literature performed Monday through Friday for the WHO Database. Search strategy as of 26 May 2021. Searches performed by Tomas Allen, Kavita Kothari, and Martha Knuth. Available from: <https://www.who.int/docs/default-source/coronaviruse/who-covid-19-database/who-covid-19_sources_searchstrategy_20210526.pdf?sfvrsn=65209cc2_5>

Canadian Agency for Drugs and Technologies in Health (2.9.21) CADTH COVID-19 Search Strings: COVID-19 — EMBASE (Internet). Available from: <https://covid.cadth.ca/literature-searching-tools/cadth-covid-19-search-strings/>

NICE (18 December 2020) [accessed 17.8.21] COVID-19 rapid guideline: managing the long-term effects of COVID-19 [NG188]. Search history record [PDF]. NICE: London. Available from: <https://www.nice.org.uk/guidance/ng188/evidence/search-strategies-pdf-8957634445>

*Observational study design filter adapted from:*

Scottish Intercollegiate Guidelines Network (SIGN). Search filters: observational studies. Medline. Edinburgh: SIGN, Last Modified 24/04/17 Available from: <https://www.sign.ac.uk/what-we-do/methodology/search-filters/>

Observational Studies - Medline. In: CADTH Search Filters Database. Ottawa: CADTH; 2023 [accessed 10.9.25]: <https://searchfilters.cadth.ca/link/37><https://searchfilters.cda-amc.ca/link/38>

#### PubMed strategy

PubMed: Ahead-of-Print (NLM): 2021-2025/09/11

Searched 11.9.25

<https://pubmed.ncbi.nlm.nih.gov/searches/7500133/?mode=full&sort=date>

61	#13 AND #45 AND #23 AND #60 AND #14	535

60	#46 OR #47 OR #48 OR #49 OR #50 OR #51 OR #52 OR #53 OR #54 OR #55 OR #56 OR #57 OR #58 OR #59	5,130,114

59	(multifactorial[Title/Abstract] OR multi-factorial[Title/Abstract] OR multiple[Title/Abstract] OR concurrent[Title/Abstract] OR simultaneous[Title/Abstract]) AND (condition[Title/Abstract] OR conditions[Title/Abstract] OR disease[Title/Abstract] OR diseases[Title/Abstract] OR syndrome[Title/Abstract] OR syndromes[Title/Abstract] OR disorder[Title/Abstract] OR disorders[Title/Abstract] OR illness[Title/Abstract] OR illnesses[Title/Abstract])	854,060

58	(CVD[Title/Abstract] OR CVB[Title/Abstract] OR CVA[Title/Abstract] OR CV[Title/Abstract] OR Cerebrovascular[Title/Abstract] OR cardiovascular[Title/Abstract] OR cardio-vascular[Title/Abstract] OR Cerebro-vascular[Title/Abstract]) AND (hospital[Title/Abstract] OR hospitals[Title/Abstract] OR hospitalisation[Title/Abstract] OR hospitalisations[Title/Abstract] OR hospitalization[Title/Abstract] OR hospitalizations[Title/Abstract] OR mortality[Title/Abstract] OR mortalities[Title/Abstract] OR death[Title/Abstract] OR deaths[Title/Abstract] OR admission[Title/Abstract] OR admissions[Title/Abstract])	227,008

57	"multiple chronic conditions"[Mesh]	846

56	"comorbidity"[Mesh:NoExp]	131,753

55	pluralpathology[Title/Abstract] OR pluralpathologies[Title/Abstract] OR plural-pathology[Title/Abstract] OR plural-patholologies[Title/Abstract] OR pluralmorbid[Title/Abstract] OR pluralmorbidity[Title/Abstract] OR pluralmorbidities[Title/Abstract] OR plural-morbid[Title/Abstract] OR plural-morbidity[Title/Abstract] OR plural-morbidities[Title/Abstract] OR "dual diagnosis"[Title/Abstract] OR "dual diagnoses"[Title/Abstract] OR "plural diagnosis"[Title/Abstract] OR "plural diagnoses"[Title/Abstract]	2,469

54	co-morbid[Title/Abstract] OR comorbid[Title/Abstract] OR co-morbidity[Title/Abstract] OR comorbidity[Title/Abstract] OR co-morbidities[Title/Abstract] OR comorbidities[Title/Abstract]	317,550

53	(cerebrovasculopathy[Title/Abstract] OR cerebrovasculopathies[Title/Abstract] OR cerebro-vasculopathy[Title/Abstract] OR cerebro-vasculopathies[Title/Abstract] OR cerebroangiopathy[Title/Abstract] OR cerebroangiopathies[Title/Abstract] OR cerebro-angiopathy[Title/Abstract] OR cerebro-angiopathies[Title/Abstract] OR "cerebral vasculopathy"[Title/Abstract] OR "cerebral vasculopathies"[Title/Abstract] OR "cerebral angiopathy"[Title/Abstract] OR "cerebral angiopathies"[Title/Abstract] OR "brain vasculopathy"[Title/Abstract] OR "brain vasculopathies"[Title/Abstract] OR "brain angiopathy"[Title/Abstract] OR "brain angiopathies"[Title/Abstract] OR Angiocardiopathy[Title/Abstract] OR Angiocardiopathies[Title/Abstract] OR "Angio-cardiopathy"[Title/Abstract] OR "Angio-cardiopathies"[Title/Abstract])	793

52	(cardiovascular[Title/Abstract] OR cardio-vascular[Title/Abstract] OR angiocardiovascular[Title/Abstract] OR angio-cardiovascular[Title/Abstract] OR angio-cardio-vascular[Title/Abstract] OR cerebrovascular[Title/Abstract] OR cerebro-vascular[Title/Abstract] OR "cerebral vascular"[Title/Abstract] OR heart[Title/Abstract] OR cardiac[Title/Abstract] OR myocardial[Title/Abstract] OR cerebral[Title/Abstract]) AND (complications[Title/Abstract] OR complications[Title/Abstract] OR disorder[Title/Abstract] OR disorders[Title/Abstract] OR syndrome[Title/Abstract] OR syndromes[Title/Abstract] OR comorbidity[Title/Abstract] OR comorbidities[Title/Abstract] OR co-morbidity[Title/Abstract] OR co-morbidities[Title/Abstract] OR disease[Title/Abstract] OR diseases[Title/Abstract] OR lesion[Title/Abstract] OR lesions[Title/Abstract] OR disturbance[Title/Abstract] OR disturbances[Title/Abstract] OR event[Title/Abstract] OR events[Title/Abstract] OR syndrome[Title/Abstract] OR syndromes[Title/Abstract] OR damage[Title/Abstract] OR damaging[Title/Abstract] OR damages[Title/Abstract] OR injury[Title/Abstract] OR injuries[Title/Abstract] OR failure[Title/Abstract] OR failures[Title/Abstract] OR pathology[Title/Abstract] OR pathologies[Title/Abstract])	1,576,252

51	(Circulatory[Title/Abstract] OR Cardiac[Title/Abstract] OR vascular[Title/Abstract]) AND (injury[Title/Abstract] OR injuries[Title/Abstract] OR damage[Title/Abstract] OR damages[Title/Abstract] OR pathology[Title/Abstract] OR pathologies[Title/Abstract] OR failure[Title/Abstract] OR failures[Title/Abstract] OR comorbidity[Title/Abstract] OR comorbidities[Title/Abstract] OR co-morbidity[Title/Abstract] OR co-morbidities[Title/Abstract] OR complication[Title/Abstract] OR complications[Title/Abstract] OR disorder[Title/Abstract] OR disorderS[Title/Abstract] OR syndrome[Title/Abstract] OR syndromes[Title/Abstract] OR disease[Title/Abstract] OR diseases[Title/Abstract] OR lesion[Title/Abstract] OR lesions[Title/Abstract] OR disturbance[Title/Abstract] OR disturbances[Title/Abstract] OR syndrome[Title/Abstract] OR syndromes[Title/Abstract] OR event[Title/Abstract] OR events[Title/Abstract])	900,144

50	"Atrial fibrillation"[Title/Abstract] OR Thrombosis[Title/Abstract] OR thrombotic[Title/Abstract] OR thromboses[Title/Abstract] OR Stroke[Title/Abstract] OR strokes[Title/Abstract] OR "blood clot"[Title/Abstract] OR "blood clots"[Title/Abstract] OR Hypertension[Title/Abstract] OR "Myocardial infarction"[Title/Abstract] OR "Transient Ischemic Attack"[Title/Abstract] OR "pulmonary embolus"[Title/Abstract] OR "pulmonary emboli"[Title/Abstract] OR "Congenital Heart Problem"[Title/Abstract]	1,291,256

49	(Congenital[Title/Abstract] OR coronary[Title/Abstract] OR ischaemic[Title/Abstract] OR ischemic[Title/Abstract] OR heart[Title/Abstract] OR congestive[Title/Abstract] OR cardiac[Title/Abstract] OR Atherosclerotic[Title/Abstract] OR cardiovascular[Title/Abstract] OR cardio-vascular[Title/Abstract]) AND (failure[Title/Abstract] OR failures[Title/Abstract] OR disease[Title/Abstract] OR diseases[Title/Abstract] OR attack[Title/Abstract] OR attacks[Title/Abstract] OR embolus[Title/Abstract] OR emboli[Title/Abstract] OR clot[Title/Abstract] OR clots[Title/Abstract])	1,192,501

48	("Peripheral vascular"[Title/Abstract] OR Cerebrovascular[Title/Abstract] OR cardiovascular[Title/Abstract] OR "cardio-vascular"[Title/Abstract] OR "Cerebro-vascular"[Title/Abstract]) AND (accident[Title/Abstract] OR accidents[Title/Abstract] OR failure[Title/Abstract] OR failures[Title/Abstract] OR attack[Title/Abstract] OR attacks[Title/Abstract] OR clot[Title/Abstract] OR clots[Title/Abstract] OR embolus[Title/Abstract] OR emboli[Title/Abstract] OR disease[Title/Abstract] OR diseases[Title/Abstract] OR comorbidity[Title/Abstract] OR comorbidities[Title/Abstract] OR co-morbidities[Title/Abstract] OR co-morbidity[Title/Abstract])	492,712

47	(CVD[Title/Abstract] OR CBV[Title/Abstract] OR CHD[Title/Abstract] OR CBA[Title/Abstract] OR CBAs[Title/Abstract] OR TIA[Title/Abstract] OR TIAs[Title/Abstract] OR IHD[Title/Abstract] OR MI[Title/Abstract] OR CVA[Title/Abstract] OR CVAs[Title/Abstract] OR AF[Title/Abstract])	261,097

46	("Cardiovascular Diseases"[Mesh]) OR "Cerebrovascular Disorders"[Mesh]	2,890,091

45	#24 OR #25 OR #26 OR #27 OR #28 OR #9 OR #30 OR #31 OR #32 OR #33 OR #34 OR #35 OR #36 OR #37 OR #38 OR #39 OR #40 OR #41 OR #42 OR #43 OR #44	5,002,392

44	"2696235-99-9" - Schema: all	0

43	("sp/gsk"[Title/Abstract] OR "subunit B.1.351"[Title/Abstract] OR "Sanofi GSK"[Title/Abstract]) AND (vaccine[Title/Abstract] OR vaccination[Title/Abstract] OR vaccinations[Title/Abstract] OR jab[Title/Abstract] OR jabs[Title/Abstract] OR shot[Title/Abstract] OR shots[Title/Abstract] OR immunise[Title/Abstract] OR immunisation[Title/Abstract] OR immunisations[Title/Abstract] OR immunize[Title/Abstract] OR immunization[Title/Abstract] OR immunizations[Title/Abstract])	3

42	("vidprevtyn beta"[Title/Abstract] OR "vidprevtynbeta"[Title/Abstract] OR "vidprevtynb"[Title/Abstract] OR "vidprevtyn-b"[Title/Abstract] OR "VAT0000"[Title/Abstract] OR "VAT00008"[Title/Abstract] OR "VAT-0000"[Title/Abstract] OR "VAT-00008"[Title/Abstract])	1

41	"2695500-31-1" - Schema: all	0

40	("VLA2001"[Title/Abstract] OR "VLA2101"[Title/Abstract] OR "VLA-2001"[Title/Abstract] OR "VLA-2101"[Title/Abstract] OR Valneva[Title/Abstract])	33

39	"2502099-58-1" - Schema: all	0

38	(Novavax[Title/Abstract] OR CovoVax[Title/Abstract] OR Nuvaxovid[Title/Abstract] OR "NVXCoV2373"[Title/Abstract] OR "NVX-CoV2373"[Title/Abstract] OR "nvx cov 2373"[Title/Abstract] OR "nvx cov2373"[Title/Abstract] OR "NVXCoV-2373"[Title/Abstract] OR " NVX-CoV2601"[Title/Abstract] OR " NVXCoV2601"[Title/Abstract] OR " NVX-CoV-2601"[Title/Abstract] OR " NVXCoV-2601"[Title/Abstract] OR "sars-cov-2 rs"[Title/Abstract] OR "sarscov-2 rs"[Title/Abstract] OR "sarscov2 rs"[Title/Abstract] OR "sars-cov-2rs"[Title/Abstract] OR "sarscov2rs"[Title/Abstract] OR "sarscov-2rs"[Title/Abstract] OR "tak 019"[Title/Abstract] OR "tak019"[Title/Abstract])	292

37	"2420395-83-9" - Schema: all	0

36	("Oxford-AstraZeneca"[Title/Abstract] OR AstraZeneca[Title/Abstract] OR AZ[Title/Abstract]) AND (vaccine[Title/Abstract] OR vaccination[Title/Abstract] OR vaccinations[Title/Abstract] OR jab[Title/Abstract] OR jabs[Title/Abstract] OR shot[Title/Abstract] OR shots[Title/Abstract] OR immunise[Title/Abstract] OR immunisation[Title/Abstract] OR immunisations[Title/Abstract] OR immunize[Title/Abstract] OR immunization[Title/Abstract] OR immunizations[Title/Abstract])	1,835

35	(Vaxzevria[Title/Abstract] OR Covishield[Title/Abstract] OR "AZD1222"[Title/Abstract] OR "AZD-1222"[Title/Abstract] OR "AZD2816"[Title/Abstract] OR "AZD-2816"[Title/Abstract] OR "ChAdOx1"[Title/Abstract] OR "ChAdOx-1"[Title/Abstract] OR "ChAdOx1-s"[Title/Abstract] OR "ChAdOx1s"[Title/Abstract] OR "ChAdOx-1-s"[Title/Abstract] OR "ChAdOx-1s"[Title/Abstract])	2,888

34	"2541607-46-7" - Schema: all	0

33	(Janssen[Title/Abstract] OR "J&J"[Title/Abstract] OR "Johnson & Johnson"[Title/Abstract]) AND (vaccine[Title/Abstract] OR vaccines[Title/Abstract] OR vaccination[Title/Abstract] OR vaccinations[Title/Abstract] OR jab[Title/Abstract] OR jabs[Title/Abstract] OR shot[Title/Abstract] OR shots[Title/Abstract] OR immunise[Title/Abstract] OR immunisation[Title/Abstract] OR immunisations[Title/Abstract] OR immunize[Title/Abstract] OR immunization[Title/Abstract] OR immunizations[Title/Abstract])	754

32	(Jcovden[Title/Abstract] OR ibacovavec[Title/Abstract] OR "Ad26.COV2.S"[Title/Abstract] OR "JNJ-78436735"[Title/Abstract] OR "JNJ78436735"[Title/Abstract] OR "Ad26COVS1"[Title/Abstract] OR "VAC31518"[Title/Abstract] OR "VAC-31518"[Title/Abstract])	685

31	"ChAdOx1 nCoV-19"[Mesh:NoExp]	1,350

30	"2430046-03-8" OR "2457298-05-2" - Schema: all	0

29	(Elasomeran[Title/Abstract] OR moderna[Title/Abstract] OR Spikevax[Title/Abstract] OR davesomeran[Title/Abstract] OR imelasomeran[Title/Abstract] OR andusomeran[Title/Abstract] OR "mRNA 1273"[Title/Abstract] OR "mRNA1273"[Title/Abstract] OR "mRNA-1273.211"[Title/Abstract] OR "mRNA 1273.211"[Title/Abstract] OR Spikevax[Title/Abstract] OR "M-1273"[Title/Abstract] OR "M1273"[Title/Abstract] OR "CX-024414"[Title/Abstract] OR "CX024414"[Title/Abstract] OR "TAK-919"[Title/Abstract] OR "TAK919"[Title/Abstract] OR "mRNA1273"[Title/Abstract] OR "mRNA-1273"[Title/Abstract] OR "messenger RNA1273"[Title/Abstract] OR "messenger RNA 1273"[Title/Abstract])	3,751

28	"2019-nCoV Vaccine mRNA-1273"[Mesh:NoExp]	1,060

27	"2417899-77-3" - Schema: all	0

26	(tozinameran OR Comirnaty OR "Pfizer-BioNTech" OR "pf07302048" OR "pf-07302048" OR " BNT-162b2" OR "BNT162b2" OR "rbp 020.2" OR " rbp020.2" OR Pidacmeran OR "BNT162C2" OR "BNT-162C2" OR Abdavomeran OR "BNT-162B1" OR "BNT162B1" OR "BNT162A1" OR "BNT-162A1")	7,985

25	(pfizer[Title/Abstract] OR biontech[Title/Abstract]) AND (vaccins[Title/Abstract] OR vaccines[Title/Abstract] OR jab[Title/Abstract] OR jabs[Title/Abstract] OR shot[Title/Abstract] OR shots[Title/Abstract] OR immuniss[Title/Abstract] OR immunisation[Title/Abstract] OR immunisations[Title/Abstract] OR immunize ORimmunization[Title/Abstract] OR immunizations[Title/Abstract])	2,805

24	"BNT162 Vaccine"[Mesh:NoExp]	4,285

23	#19 OR #22	7,773,253

22	#21 OR #20	1,542,539

21	"electronic medical record"[Title/Abstract] OR EMR[Title/Abstract] OR "ELECTRONIC MEDICAL RECORDS"[Title/Abstract] OR dataset[Title/Abstract] OR DATASETS[Title/Abstract] OR "data set"[Title/Abstract] OR "DATA SETS" "administrative claim"[Title/Abstract] OR "administrative claims"[Title/Abstract] OR "pragmatic trial"[Title/Abstract] OR prct[Title/Abstract] OR "pragmatic trial"[Title/Abstract]	6,239

20	"real world"[Title/Abstract] OR RWE[Title/Abstract] OR RWD[Title/Abstract] OR "Real-life"[Title/Abstract] OR registry[Title/Abstract] OR register[Title/Abstract] OR REGISTERS[Title/Abstract] OR survey[Title/Abstract] OR SURVEYS[Title/Abstract] OR "patient record"[Title/Abstract] OR "PATIENT RECORDS"[Title/Abstract] OR "medical record"[Title/Abstract] OR "MEDICAL RECORDS"[Title/Abstract] OR "electronic health record"[Title/Abstract] OR "ELECTRONIC HEALTH RECORDS"[Title/Abstract] OR EHR[Title/Abstract]	1,538,356

19	#15 OR #16 OR #17 OR #18	6,839,427

18	"Case control"[Title/Abstract] OR "cohort study"[Title/Abstract] OR "cohort studies"[Title/Abstract] OR "cohort analysis"[Title/Abstract] OR "cohort analyses"[Title/Abstract] OR "follow up study"[Title/Abstract] OR "follow up studies"[Title/Abstract] OR "observational study"[Title/Abstract] OR "observational studies"[Title/Abstract] OR Longitudinal[Title/Abstract] OR Retrospective[Title/Abstract] OR "Cross sectional"[Title/Abstract] OR "epidemiological studies"[Title/Abstract] OR " epidemiological study"[Title/Abstract]	2,493,638

17	"case reports"[Publication Type] OR "Observational Study"[Publication Type] OR "Validation Studies"[Publication Type]	2,683,347

16	"Observational Studies as Topic"[Mesh] OR "single-case studies as topic"[Mesh] OR "case reports as topic"[Mesh]	11,731

15	"Epidemiologic Studies"[Mesh:NoExp] OR "Epidemiologic Methods"[Mesh:NoExp] OR "Case-Control Studies"[Mesh] OR "Cohort Studies"[Mesh] OR "Cross-Sectional Studies"[Mesh:NoExp]	3,532,560

14	pubstatusaheadofprint OR publisher[sb] OR pubmednotmedline[sb]	6,553,674

13	#11 NOT #12	429,187

12	LETTER[Publication Type] OR EDITORIAL[Publication Type] OR COMMENT[Publication Type]	2,351,850

11	#7 NOT #10	482,502

10	#9 NOT (#8 AND #9)	3,969,737

9	rat[tiab] OR rats[tiab] OR mouse[tiab] OR mice[tiab] OR murine[tiab] OR rodent[tiab] OR rodents[tiab] OR hamster[tiab] OR hamsters[tiab] OR pig[tiab] OR pigs[tiab] OR porcine[tiab] OR rabbit[tiab] OR rabbits[tiab] OR animal[tiab] OR animals[tiab] OR dogs[tiab] OR dog[tiab] OR cats[tiab] OR cow[tiab] OR bovine[tiab] OR sheep[tiab] OR ovine[tiab] OR monkey[tiab] OR monkeys[tiab]	4,990,792

8	Human[tiab] OR humans[tiab]	3,511,491

7	#1 OR #4 OR #5 OR #6	494,310

6	"Covid 2019"[Title/Abstract] OR ncov[Title/Abstract] OR "sars cov"[Title/Abstract] OR sarscov[Title/Abstract] OR "sars coronavirus"[Title/Abstract] OR coronovirus[Title/Abstract] OR "corono virus"[Title/Abstract] OR "19-nCoV"[Title/Abstract] OR "19nCoV"[Title/Abstract] OR "SARS2"[Title/Abstract] OR "SARS-2"[Title/Abstract] OR SARScoronavirus[Title/Abstract] OR "SARS-coronavirus"[Title/Abstract] OR SARScoronovirus[Title/Abstract] OR "SARS-coronovirus"[Title/Abstract] OR "HCoV-19"[Title/Abstract] OR "HCoV19"[Title/Abstract] OR "HCoV-2019"[Title/Abstract] OR "HCoV2019"[Title/Abstract] OR "2019 novel"[Title/Abstract] OR "Severe Acute Respiratory Syndrome Coronavirus 2"[Title/Abstract] OR "Severe Acute Respiratory Syndrome Corona Virus 2"[Title/Abstract]	167,522

5	"Coronavirus"[Title/Abstract] OR "covid 19"[Title/Abstract] OR "2019-ncov"[Title/Abstract] OR "2019-ncov"[Title/Abstract] OR "2019ncov"[Title/Abstract] OR "corona-virus"[Title/Abstract] OR "cov19"[Title/Abstract] OR "cov-19"[Title/Abstract] OR "19nCoV"[Title/Abstract] OR "COVID-19"[Title/Abstract] OR "COVID2019"[Title/Abstract]	456,347

4	#2 AND #3	92,967

3	(("Epidemics"[Mesh:NoExp]) OR "Pandemics"[Mesh:NoExp]) OR "Disease Outbreaks"[Mesh:NoExp]	246,424

2	((("Coronaviridae"[Mesh:NoExp]) OR "Coronavirus"[Mesh]) OR "Betacoronavirus"[Mesh:NoExp]) OR "Coronavirus Infections"[Mesh:NoExp]	222,159

1	("COVID-19"[Mesh]) OR "Severe acute respiratory syndrome-related coronavirus"[Mesh]	309,688

*COVID facet based on terms from:*

 World Health Organization (26 May 2021) WHO COVID-19 Database Search Strategy. Systematic search of the COVID-19 literature performed Monday through Friday for the WHO Database. Search strategy as of 26 May 2021. Searches performed by Tomas Allen, Kavita Kothari, and Martha Knuth. Available from: <https://www.who.int/docs/default-source/coronaviruse/who-covid-19-database/who-covid-19_sources_searchstrategy_20210526.pdf?sfvrsn=65209cc2_5>

Canadian Agency for Drugs and Technologies in Health (2.9.21) CADTH COVID-19 Search Strings: COVID-19 — EMBASE (Internet). Available from: <https://covid.cadth.ca/literature-searching-tools/cadth-covid-19-search-strings/>

NICE (18 December 2020) [accessed 17.8.21] COVID-19 rapid guideline: managing the long-term effects of COVID-19 [NG188]. Search history record [PDF]. NICE: London. Available from: <https://www.nice.org.uk/guidance/ng188/evidence/search-strategies-pdf-8957634445>

*Observational study design filter adapted from:*

Scottish Intercollegiate Guidelines Network (SIGN). Search filters: observational studies. Medline. Edinburgh: SIGN, Last Modified 24/04/17 Available from: <https://www.sign.ac.uk/what-we-do/methodology/search-filters/>

Observational Studies - Medline. In: CADTH Search Filters Database. Ottawa: CADTH; 2023 [accessed 10.9.25]: <https://searchfilters.cadth.ca/link/37><https://searchfilters.cda-amc.ca/link/38>

### Meta-analysis feasibility assessment

All 20 studies reporting the impact of pre-infection vaccination on risk of cardiovascular, cerebrovascular, or venous thromboembolism outcomes after SARS-CoV-2 infection were included in the meta-analysis feasibility assessment. Study populations, data source, vaccination characteristics, risk measure, and study quality (risk of bias) were assessed for comparability. Where there were concerns about heterogeneity across studies, studies were excluded or sensitivity analyses were performed. A minimum of three studies was required for a meta-analysis to be performed.

The following studies were excluded from meta-analyses:

- El Seblani 2025^22^: only included individuals with COVID-associated ischaemic stroke (prior to assessment of the outcome of interest).
- Madrid 2024^16^ and Sritharan 2024^19^: considered at very high risk of bias.
- Wee 2025^26^: only study to compare bivalent and monovalent booster doses.
- Zisis 2022,^8^ Madrid 2024,^16^ Meister 2025,^24^ O’Carroll 2024,^17^ Yu 2024,^21^ Lin 2025,^25^ Montone 2025^25^: reported relative risk/risk ratios, incidence rate ratios, or odds ratios (within risk measures, fewer than three studies reporting comparable outcomes).

Among the ten studies reporting hazard ratios suitable for meta-analysis of the association between vaccination and post-COVID-19 outcomes, the most comparable exposure definition across studies was selected for the primary analyses. Where studies reported outcomes by vaccine dose, the most comparable dose category (2 or ≥2 doses) was included. Where studies reported outcomes at multiple post-infection time points, the time point most comparable to those used in other studies within the same meta-analysis was selected.

Sensitivity analyses were performed as follows:

- “Lowest dose”: For studies reporting outcomes by vaccine dose number, the hazard ratio corresponding to the lowest reported dose category was selected.
- “Highest dose”: For studies reporting outcomes by vaccine dose number, the hazard ratio corresponding to the highest reported dose category was selected.
- “Shortest follow-up”: For studies reporting outcomes at multiple post-infection time points, the hazard ratio for the shortest follow-up period was selected.
- “Longest follow-up”: For studies reporting outcomes at multiple post-infection time points, the hazard ratio for the longest follow-up period was selected.
- “Add CVD- and CeVD-only composite outcomes”: Two studies reporting either only composite cardiovascular outcomes (Al-Aly 2022^4^) or composite cerebrovascular outcomes (Chen 2024^12^) were included in the composite outcomes meta-analysis.
- “Remove potential population overlap”: Where potential overlap in study populations was identified, the study with the smaller population size was excluded (Taquet 2022^6^ in the stroke meta-analysis, and Kim 2024^14^ in the venous thromboembolism meta-analysis).
- “Replace with acute coronary disease outcome”: In the coronary syndrome meta-analysis, the myocardial infarction outcome reported by Al-Aly 2022^4^ was replaced with the acute coronary disease outcome.
- “Remove only myocardial infarction outcomes”: In the coronary syndrome meta-analysis, studies reporting the outcome only as myocardial infarction were excluded (Al-Aly 2022^4^ and Kim 2022^5^).
- “Remove outcomes pooled with cardiomyopathies”: In the heart failure meta-analysis, the outcome reported by Huh 2024^13^ (heart failure and cardiomyopathies) was excluded.
- “Replace with other dysrhythmias outcome”: In the arrhythmia meta-analysis, the atrial fibrillation outcome reported by Al-Aly 2022^4^ was replaced with the other dysrhythmias outcome.
- “Replace with pulmonary embolism outcome”: In the venous thromboembolism meta-analysis, the deep vein thrombosis or venous thromboembolism outcomes reported by Al-Aly 2022,^4^ Huh 2024,^13^ and Kim 2024^14^ were replaced with the pulmonary embolism outcomes.

### References

1.	WHO. WHO announces simple, easy-to-say labels for SARS-CoV-2 Variants of Interest and Concern [Internet]. 2021 [cited 2026 Mar 10]. Available from: https://www.emro.who.int/media/news/who-announces-simple-easy-to-say-labels-for-sars-cov-2-variants-of-interest-and-concern.html

2.	WHO. Classification of Omicron (B.1.1.529): SARS-CoV-2 Variant of Concern [Internet]. 2021 [cited 2026 Mar 10]. Available from: https://www.who.int/news/item/26-11-2021-classification-of-omicron-(b.1.1.529)-sars-cov-2-variant-of-concern

3.	Page MJ, McKenzie JE, Bossuyt PM, Boutron I, Hoffmann TC, Mulrow CD, et al. The PRISMA 2020 statement: an updated guideline for reporting systematic reviews. BMJ. 2021 Mar 29;372:n71. doi:10.1136/bmj.n71 PubMed PMID: 33782057.

4.	Al-Aly Z, Bowe B, Xie Y. Long COVID after breakthrough SARS-CoV-2 infection. Nat Med. 2022 Jul;28(7):7. doi:10.1038/s41591-022-01840-0

5.	Kim YE, Huh K, Park YJ, Peck KR, Jung J. Association Between Vaccination and Acute Myocardial Infarction and Ischemic Stroke After COVID-19 Infection. JAMA. 2022 Sep 6;328(9):887–9. doi:10.1001/jama.2022.12992

6.	Taquet M, Dercon Q, Harrison PJ. Six-month sequelae of post-vaccination SARS-CoV-2 infection: A retrospective cohort study of 10,024 breakthrough infections. Brain, Behavior, and Immunity. 2022 Jul 1;103:154–62. doi:10.1016/j.bbi.2022.04.013

7.	Xie J, Prats-Uribe A, Feng Q, Wang Y, Gill D, Paredes R, et al. Clinical and Genetic Risk Factors for Acute Incident Venous Thromboembolism in Ambulatory Patients With COVID-19. JAMA Intern Med. 2022 Oct 1;182(10):1063–70. doi:10.1001/jamainternmed.2022.3858

8.	Zisis SN, Durieux JC, Mouchati C, Perez JA, McComsey GA. The Protective Effect of Coronavirus Disease 2019 (COVID-19) Vaccination on Postacute Sequelae of COVID-19: A Multicenter Study From a Large National Health Research Network. Open Forum Infect Dis. 2022 Jul 1;9(7):ofac228. doi:10.1093/ofid/ofac228

9.	Jiang J, Chan L, Kauffman J, Narula J, Charney AW, Oh W, et al. Impact of Vaccination on Major Adverse Cardiovascular Events in Patients With COVID-19 Infection. JACC. 2023 Mar 7;81(9):928–30. doi:10.1016/j.jacc.2022.12.006

10.	Wan EYF, Mok AHY, Yan VKC, Chan CIY, Wang B, Lai FTT, et al. Association between BNT162b2 and CoronaVac vaccination and risk of CVD and mortality after COVID-19 infection: A population-based cohort study. Cell Reports Medicine. 2023 Oct 17;4(10):101195. doi:10.1016/j.xcrm.2023.101195

11.	Cezard GI, Denholm RE, Knight R, Wei Y, Teece L, Toms R, et al. Impact of vaccination on the association of COVID-19 with cardiovascular diseases: An OpenSAFELY cohort study. Nat Commun. 2024 Mar 11;15(1):2173. doi:10.1038/s41467-024-46497-0

12.	Chen SY, Hsieh TYJ, Hung YM, Oh JW, Chen SK, Wang SI, et al. Prior COVID-19 vaccination and reduced risk of cerebrovascular diseases among COVID-19 survivors. Journal of Medical Virology. 2024;96(5):e29648. doi:10.1002/jmv.29648

13.	Huh K, Kim YE, Bae GH, Moon JY, Kang JM, Lee J, et al. Vaccination and the risk of post-acute sequelae after COVID-19 in the Omicron-predominant period. Clinical Microbiology and Infection. 2024 May 1;30(5):666–73. doi:10.1016/j.cmi.2024.01.028 PubMed PMID: 38331252.

14.	Kim HJ, Jeong S, Song J, Park SJ, Park YJ, Oh YH, et al. Risk of pulmonary embolism and deep vein thrombosis following COVID-19: a nationwide cohort study. MedComm. 2024;5(7):e655. doi:10.1002/mco2.655

15.	Lim JT, Liang En W, Tay AT, Pang D, Chiew CJ, Ong B, et al. Long-term Cardiovascular, Cerebrovascular, and Other Thrombotic Complications in COVID-19 Survivors: A Retrospective Cohort Study. Clin Infect Dis. 2024 Jan 15;78(1):70–9. doi:10.1093/cid/ciad469

16.	Madrid J, Agarwal P, Müller-Peltzer K, Benning L, Selig M, Rolauffs B, et al. Cardioprotective effects of vaccination in hospitalized patients with COVID-19. Clin Exp Med. 2024 May 17;24(1):103. doi:10.1007/s10238-024-01367-3

17.	O’Carroll A, Richard SA, Byrne C, Rusiecki J, Wier B, Berjohn CM, et al. Estimating the Effect of Coronavirus Disease 2019 (COVID-19) Vaccination and Infection Variant on Post-COVID-19 Venous Thrombosis or Embolism Risk. Open Forum Infect Dis. 2024 Nov 1;11(11):ofae557. doi:10.1093/ofid/ofae557

18.	Song J, Choi S, Jeong S, Chang J young, Park SJ, Oh YH, et al. Protective effect of vaccination on the risk of cardiovascular disease after SARS-CoV-2 infection. Clin Res Cardiol. 2024 Feb 1;113(2):235–45. doi:10.1007/s00392-023-02271-8

19.	Sritharan HP, Bhatia KS, van Gaal W, Kritharides L, Chow CK, Bhindi R. Cardiovascular outcomes for people hospitalised with COVID-19 in Australia, and the effect of vaccination: an observational cohort study. Medical Journal of Australia. 2024;220(10):517–22. doi:10.5694/mja2.52307

20.	Tran HNQ, Risk M, Nair GB, Zhao L. Risk benefit analysis to evaluate risk of thromboembolic events after mRNA COVID-19 vaccination and COVID-19. npj Vaccines. 2024 Sep 13;9(1):166. doi:10.1038/s41541-024-00960-7

21.	Yu Q, Fan M, Lin CJ, Lui DTW, Tan KCB, Yiu KH, et al. Risk factors for long-term cardiovascular post-acute sequelae of COVID-19 infection: A nested case-control study in Hong Kong. npj Cardiovasc Health. 2024 Aug 2;1(1):10. doi:10.1038/s44325-024-00011-z

22.	El Seblani N, Gorenflo MP, Ortega-Gutierrez S, Reichwein RK, Xu R, Nagaraja N. Vaccination against COVID-19 and Outcomes in Patients with COVID-19 Infection and Stroke. npj Vaccines. 2025 Jul 1;10(1):133. doi:10.1038/s41541-025-01158-1

23.	Lin EPY, Hsu CY, Mishra S, Griffiths EA, Segal BH, Hwang C, et al. Associations of COVID-19 vaccination with risks for post-infectious cardiovascular complications: an international cohort study in cancer patients with SARS-CoV-2 infection. The Lancet Regional Health – Americas. 2025 Apr 1;44. doi:10.1016/j.lana.2025.101038

24.	Meister T, Maiväli Ü, Tenson K, Tisler A, Kalda R, Suija K, et al. Dynamic effects of COVID-19 vaccination on major acute cardiovascular events and mortality following SARS-CoV-2 infection in a target trial emulation study. Sci Rep. 2025 Jul 29;15(1):27530. doi:10.1038/s41598-025-13043-x

25.	Montone RA, Rinaldi R, Masciocchi C, Lilli L, Damiani A, La Vecchia G, et al. Vaccines and myocardial injury in patients hospitalized for COVID-19 infection: the CardioCOVID-Gemelli study. Eur Heart J Qual Care Clin Outcomes. 2025 Jan 1;11(1):59–67. doi:10.1093/ehjqcco/qcae016

26.	Wee LE, Lim JT, Goel M, Malek MIA, Chiew CJ, Ong B, et al. Bivalent Boosters and Risk of Postacute Sequelae Following Vaccine-Breakthrough SARS-CoV-2 Omicron Infection: A Cohort Study. Clin Infect Dis. 2025 Mar 15;80(3):520–8. doi:10.1093/cid/ciae598
